# Spectral normative modeling of brain structure

**DOI:** 10.1101/2025.01.16.25320639

**Authors:** L. Sina Mansour, Maria A. Di Biase, Chen Zhang, Fang Tian, Shaoshi Zhang, Hongwei Yan, Aihuiping Xue, Joanna Su Xian Chong, Niousha Dehestani, Eric Kwun-Kei Ng, Fang Ji, Xing Qian, Yichi Zhang, Wen Liang Loh, Joice Sin Yi Tham, Voon Hao Lew, Samantha Hui Fang Neo, Fiona Jia Wen Goh, Narayanaswamy Venketasubramanian, Eddie Chong, Nagaendran Kandiah, Ai Peng Tan, Michael J. Meaney, Marielle V. Fortier, Yap Seng Chong, Woon-Puay Koh, Vanessa Cropley, Jakob Seidlitz, Aaron Alexander-Bloch, Richard Bethlehem, Christopher Chen, Juan Helen Zhou, the Australian Imaging Biomarkers and Lifestyle Flagship Study of Ageing, the Alzheimer’s Disease Neuroimaging Initiative, the Lifespan Brain Chart Consortium, B.T. Thomas Yeo, Andrew Zalesky

## Abstract

Normative modeling in neuroscience aims to characterize interindividual variation in brain phenotypes and establish reference ranges, or brain charts, against which individuals can be compared. Normative models are typically limited to coarse spatial scales due to computational constraints, limiting their spatial specificity. Furthermore, dependence on fixed parcellation atlases limits their adaptability to alternative parcellation schemes. To overcome these key limitations, we propose *spectral normative modeling* (SNM), which leverages brain eigenmodes to efficiently generate normative ranges for arbitrarily defined regions of interest. Training SNM on over 78,000 healthy brain scans, we generate accurate lifespan thickness growth charts across different spatial scales, from millimeters to the whole brain. These charts reveal three principal thickness growth gradients, aligning neurotypical cortical change with established anatomical, genetic, and functional hierarchies. We further demonstrate SNM’s utility by elucidating high-resolution individual cortical atrophy patterns that characterize the heterogeneous expression of neurodegeneration in Alzheimer’s disease. SNM lays the groundwork for a new generation of spatially precise brain charts, offering substantial potential to drive advances in individualized precision medicine.

## 1 Introduction

Normative modeling aims to estimate population-level reference ranges for variation in a phenotype of interest, providing a methodological framework to quantify individual deviations from expected patterns^1–3^. Individuals can be benchmarked against established normative ranges while accounting for relevant covariates, such as age, sex, or other demographic, clinical, or biological variables, to determine whether, and to what extent, they fall outside a typical healthy range^4,5^. Studying such individualized deviations in relation to population trajectories has become an important goal in neuroscience, motivating the development of normative models for numerous whole-brain and regional phenotypes, such as cortical thickness and volume^6^. Prior research has demonstrated that these models can capture the heterogeneous patterns of individual deviation in brain structure^4,6^. Consequently, normative modeling has emerged as a powerful approach with significant potential to advance precision medicine^7–14^.

Normative brain charts for magnetic resonance imaging (MRI) phenotypes can, in principle, be established at the highly detailed spatial resolution at which the MRI scan is acquired. This would enable highly localized and spatially specific inference about an individual’s deviations in a cortical phenotype. However, in practice, considerations related to computational cost, high-resolution phenotype reliability, and model scalability have motivated most normative modeling studies to operate at coarser spatial resolutions. Conventional normative approaches are designed to estimate ranges for a single phenotypic summary statistic, such as mean cortical thickness averaged across the whole cortex^2,15,16^. To reach higher spatial specificity, normative brain charting studies typically repeat model-fitting for several regional summary statistics defined over a predetermined brain atlas^6,17^.

Alternatively, a mass-univariate application of conventional techniques over all voxels/vertices has been used to produce norms at high spatial resolution^8,13,18^. While enabling fine-grained inference, these approaches are sensitive to voxel-level noise and incur substantial computational cost, especially in population-wide studies using large-scale imaging biobanks. Moreover, with current normative modeling approaches, determining a reference range for a new spatial region of interest requires fitting a separate normative model anew, a process often hindered by limited access to the original training data that is typically unavailable to end-users. Establishing a methodological framework that efficiently alleviates such limitations will enable a more principled and efficient mapping of brain charts at high spatial resolution compared to the current mass-univariate approach.

Developing high-resolution normative models is challenging, particularly due to the high dimensionality of the feature space, i.e. hundreds of thousands of vertices on the cortical surface mesh. Importantly, these challenges are not unique to normative modeling, but are shared broadly across neuroimaging analyses that aim to balance spatial precision with computational tractability. For instance, spatial dependencies across vertices/voxels undermine independence assumptions and complicate the development of models that accurately explain high-resolution statistical interdependencies^19–21^. As such, finding an appropriate low-dimensional encoding of high-resolution cortical information may enable the development of computationally tractable techniques for high-resolution normative models.

Through recent advances in brain signal processing, eigenmodes constructed from the brain’s geometry and connectivity have yielded promising basis functions that can summarize phenotypic variations on the cortical surface^22–24^. As such, eigenmodes can provide a solution to high-resolution normative modeling of brain phenotypes. Spatial variation in a cortical phenotype can be captured using a lower dimensional graph spectral embedding^25,26^. We exploit this parsimonious eigenmode basis to establish normative models on the coefficients of cortical phenotypes expressed in this lower dimensional latent space. By formulating a method that relates the normative range of an arbitrary region of interest to eigenmode normative ranges, we develop a computationally efficient method that simultaneously estimates normative ranges over multiple spatial granularities.

We introduce spectral normative modeling (SNM), a framework for constructing normative reference brain charts that are independent of spatial resolution or parcellation. SNM represents high-dimensional brain phenotypes, such as cortical thickness, using a compact eigenmode representation, enabling reconstruction of normative ranges at arbitrary spatial scales. We validate SNM’s performance across multiple spatial scales and apply it to model cortical thickness norms in over 78,000 healthy individuals. Using this model, we identify three principal gradients of lifespan thickness growth that link to previously known brain hierarchies. Finally, we demonstrate the utility of this framework in characterizing individual cortical thickness deviations associated with cognitive impairment in Alzheimer’s disease (AD), highlighting its ability to generalize to unseen samples and elucidate individual heterogeneity in cortical atrophy.

## 2 Results

### 2.1 SNM: An Efficient Framework for Multi-Scale Normative Modeling

The fundamental characteristics of many engineered and natural systems can be modeled by their structural eigenmodes, which offer a simplified yet powerful means of capturing the system’s behavior. Recent advances in neuroscience have similarly demonstrated that brain eigenmodes provide a parsimonious basis for characterizing cortical information^22,25,27^. By leveraging such basis sets, we aim to enhance both the efficiency and spatial versatility of conventional normative models. In particular, we use brain connectivity eigenmodes, which naturally extend across both cortical hemispheres while preserving key neuroanatomical landmarks, such as homotopic symmetries and the separation of cortical lobes. This is especially advantageous given that pathological brain alterations often propagate along the brain’s structural network^28–34^, making connectivity eigenmodes well suited to capture normative deviations induced by mechanisms of axonal propagation. However, it is important to note that the methodological advances introduced in this work are not limited to connectivity eigenmodes and can be readily generalized to any basis set for information reconstruction.

To establish robust normative thickness trajectories across the human lifespan, we collated preprocessed structural MRI data from 78,405 healthy individuals (4–96 years) across 30 datasets and 189 sites (Fig. 1A). We leverage eigenmode representations to model age- and sex-adjusted cortical thickness norms while accounting for site effects. Conventional mass-univariate normative models typically estimate the normative range of a predefined phenotype, such as the average cortical thickness of a fixed region of interest. Here, we term these conventional models “*direct* normative models,” with the spatial region of interest referred to as a “*spatial query*.” (Fig. 1B; see Section 4.8.1 Spatial Normative Query). The spatial query thus encompasses standard brain-wide or regional averages, while also supporting flexible, weighted queries to characterize fine-grained variations, such as thickness norms in the vicinity of individual vertices.

**Fig. 1.**
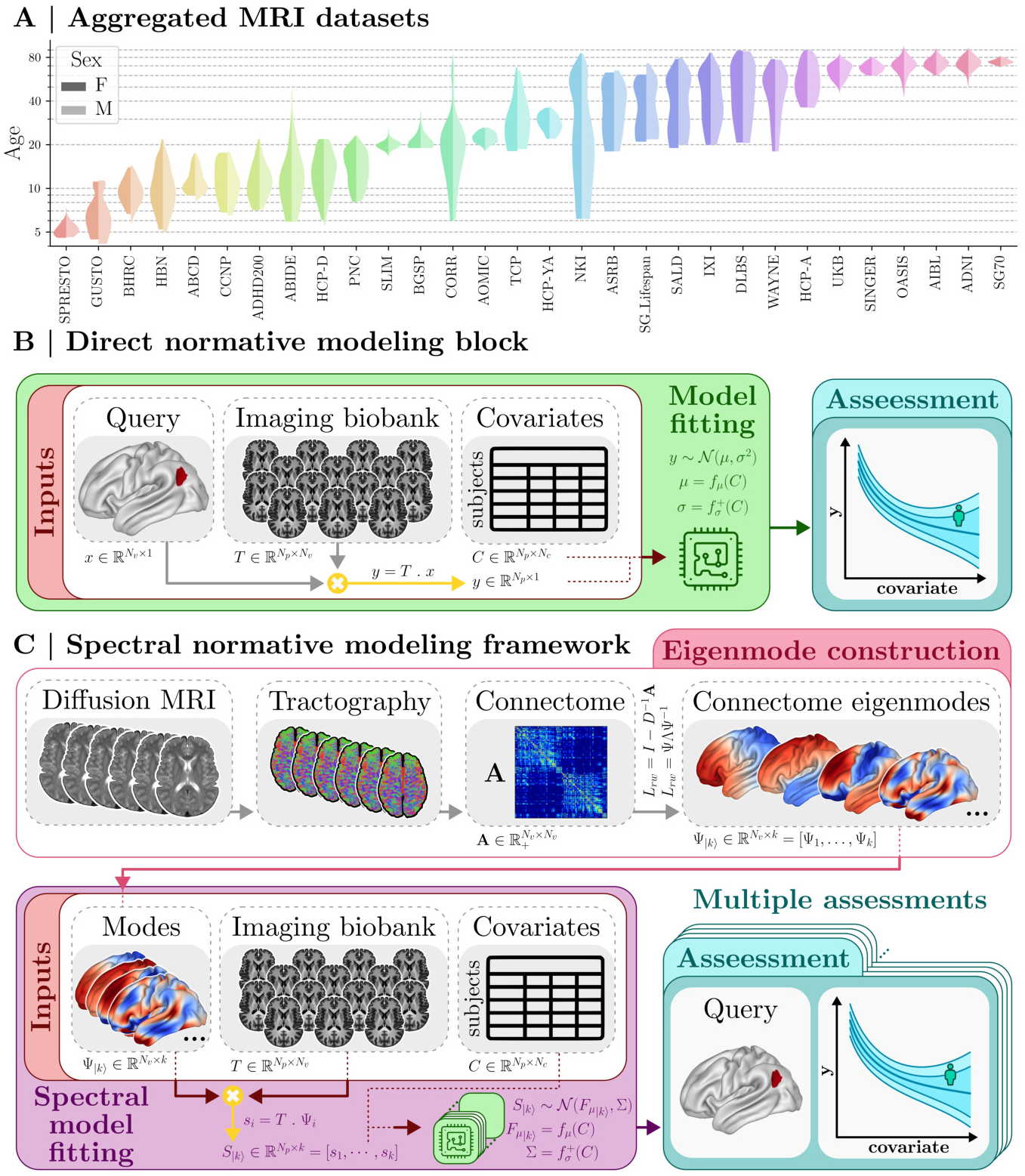
Spectral Normative Modeling (SNM) to Chart Multi-Scale Lifespan Normative Growth Trajectories. **(A)** We aggregated quality-controlled, preprocessed MRI data from 78,405 healthy participants across 30 datasets and 189 scanning sites, spanning ages 4–96 years. Violin plots show the age distributions for both sexes in each dataset. Detailed demographic information for all datasets, sites, and scanners is provided in Supplementary Table S.1. **(B)** Architecture of a conventional univariate (direct) normative modeling block, which models the distribution of a phenotype of interest *y*, obtained by applying a fixed spatial query to imaging data, as a function of covariates *C*. **(C)** Architecture of the SNM framework, which extends direct normative modeling by removing the need for a predefined spatial query. Using diffusion MRI tractography, we construct a group-level, vertex-resolution structural connectome *A* capturing the major anatomical pathways linking the cortical sheet. Laplacian eigendecomposition of *A* yields connectome eigenmodes that serve as a low-dimensional information reconstruction basis. By modeling the multivariate distribution of spectral coefficients *S*_|*k*)_ as a function of covariates *C*, a pretrained SNM (with *k* modes) can estimate normative ranges for any user-defined spatial query, enabling seamless generation of charts at arbitrary spatial resolutions, including vertex-level maps and multiple cortical parcellations.

SNM extends direct normative modeling by embedding cortical measurements into a low-dimensional eigenmode basis. Using connectivity eigenmodes computed via singular value decomposition of a random-walk Laplacian from a high-resolution group-level structural connectivity matrix (Fig. 1C), SNM proceeds in three stages: first, cortical thickness maps are projected onto the eigenmode basis; next, each eigenmode coefficient is modeled with a direct normative model to estimate its normative range while accounting for cross-basis covariance; finally, spectral estimates are combined to generate normative ranges for any spatial query without additional training. In essence, SNM synthesizes pretrained norms of eigenmode coefficients to approximate normative ranges for arbitrary spatial delineations.

### SNM Accurately Charts Population Trajectories Across Spatial Scales

Unlike mass-univariate normative approaches, which are trained on the exact phenotype associated with each spatial query, SNM does not rely on query-specific training. Instead, any query is represented through the multivariate distribution of its spectral coefficients. Each coefficient corresponds to a brain eigenmode, with early modes encoding smooth, global structure and later modes progressively capturing finer, more localized spatial detail (Fig. 2A). As a result, SNM accuracy is governed by two factors: the spatial complexity of the queried phenotype and the dimensionality of the spectral manifold, i.e., the number of eigenmodes retained. To examine their impact, we systematically evaluated performance across multiple spatial queries and spectral dimensionalities by constructing three families of queries (brain-wide, regional, and high-resolution; see Section 4.9 Model Evaluation) and training SNMs with varying numbers of eigenmodes (*k* = {10, 10^2^, 10^3^, 10^4^}).

**Fig. 2.**
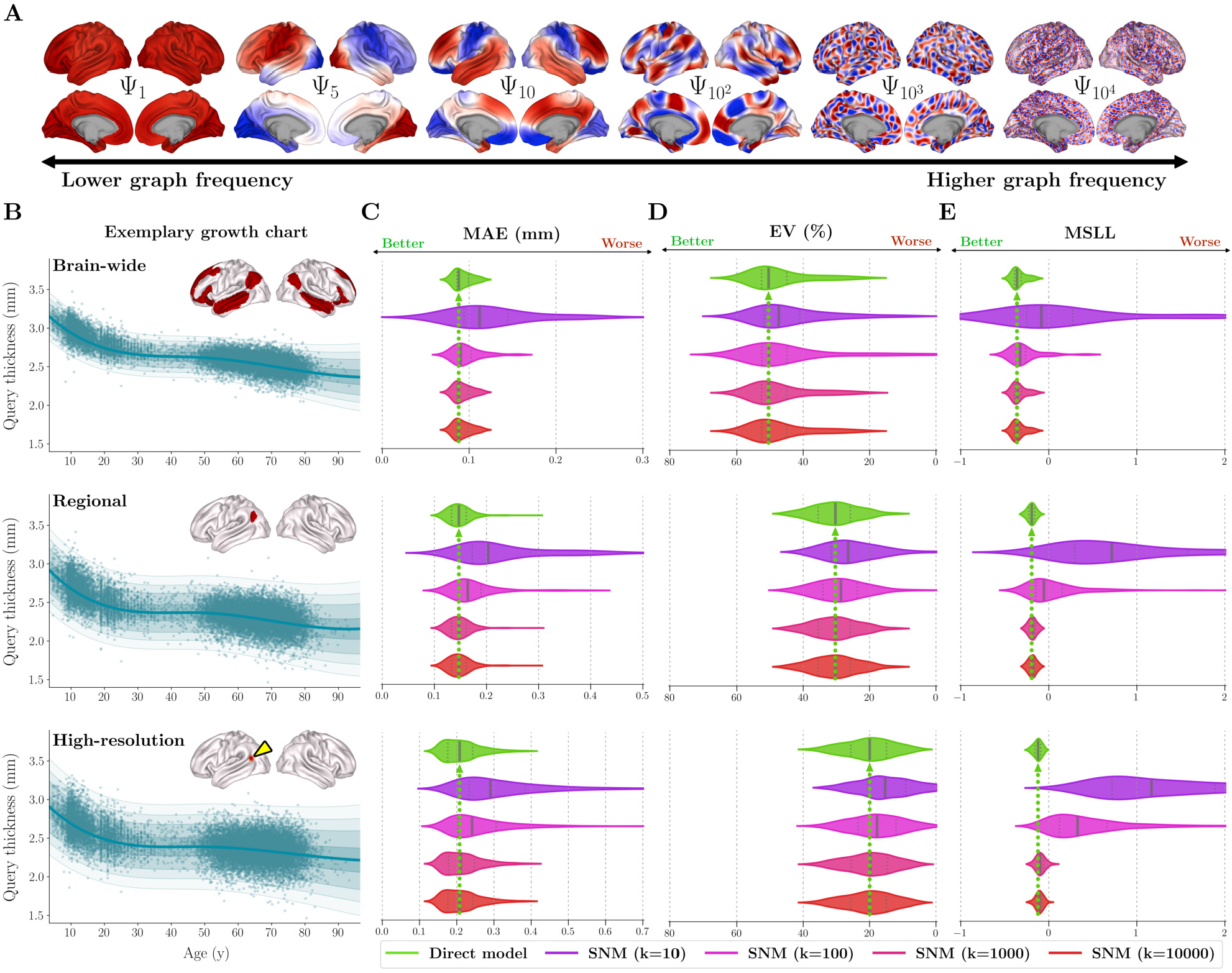
SNM Normative Performance. This figure benchmarks SNM performance across different numbers of modes (*k* = 10, 10^2^, 10^3^, 10^4^) relative to the direct approach. **(A)** Increasing the number of modes progressively increases spatial specificity: low-order modes capture smooth, global structure, whereas higher-order modes encode finer, more localized patterns. **(B)** Performance is evaluated across three spatial scales of brain-wide, regional, and high-resolution queries, with exemplary normative charts and cortical visualizations illustrating one representative query per scale. Performance metrics include mean absolute error (MAE, **C**) and explained variance (EV, **D**), which quantify accuracy in modeling central tendencies, and mean standardized log-loss (MSLL, **E**), which evaluates the full predictive distribution. For all violin plots, leftward shifts indicate better performance; green violins denote the direct model, and SNM results are shown in shades from purple to red by the number of modes. Violins reflect variability across different queries within each spatial scale. Solid and dashed lines indicate median and first/third quartiles, respectively, with a green arrow marking the direct model’s median for reference. Across all evaluations, SNMs with ≥1000 modes achieve performance comparable to the direct model.

Having established that eigenmodes accurately reconstruct cortical thickness and spatial query information (Extended Data Fig. E.1), we benchmarked SNM against direct normative models trained on the corresponding query-specific phenotypes, with the goal of assessing how closely the spectral approximation recapitulates the direct-model benchmark. Both models were trained on the same cohort of 62,724 healthy participants and evaluated on an independent held-out sample of 15,681 participants, using mean absolute error (MAE) and explained variance (EV) to assess central tendency and mean standardized log-loss (MSLL) to evaluate distributional accuracy^2^. As shown in Fig. 2, SNM accuracy increases monotonically with spectral dimensionality before plateauing at direct-model performance, with the required number of modes determined by the spatial complexity of the query: brain-wide phenotypes are well captured with as few as 100 modes, whereas regional and high-resolution queries require higher-dimensional representations reflecting finer spatial detail encoded in higher-frequency eigenmodes. Importantly, only 1000 modes, less than 2% of the original spatial dimensionality, are sufficient to match direct-model performance across all spatial scales, with additional modes yielding only marginal gains. Notably, training even the most computationally demanding configuration (*k* = 10,000) required under 9 hours on a high-performance computing node, highlighting the practical scalability of SNM even at high spectral dimensionalities and sample sizes. Supplementary analyses further characterize regional performance and the effects of query symmetry and granularity (see Supplementary Section S.1.7 Cortical Projections of Performance Metrics and Section S.1.8 Sensitivity Analyses for Query Characteristics).

### 2.3 SNM Enables High-Resolution Normative Brain Charting

A key advance of SNM is its ability to concurrently generate normative ranges for multiple spatial queries using a single pretrained model (Fig. 3A). By representing cortical phenotypes in a low-dimensional graph–spectral manifold, SNM enables efficient, vectorized inference for many regions of interest, eliminating the need to train separate models per vertex. Using the fitted model with *k* = 1000 modes (SNM-1000), we produce vertex-wise normative charts with an approximately 98% reduction in computation time relative to mass-univariate computations. We leverage this efficiency to derive high-resolution maps of the population mean and variance of cortical thickness, as well as the percent yearly change estimated from the derivative of the normative median.

**Fig. 3.**
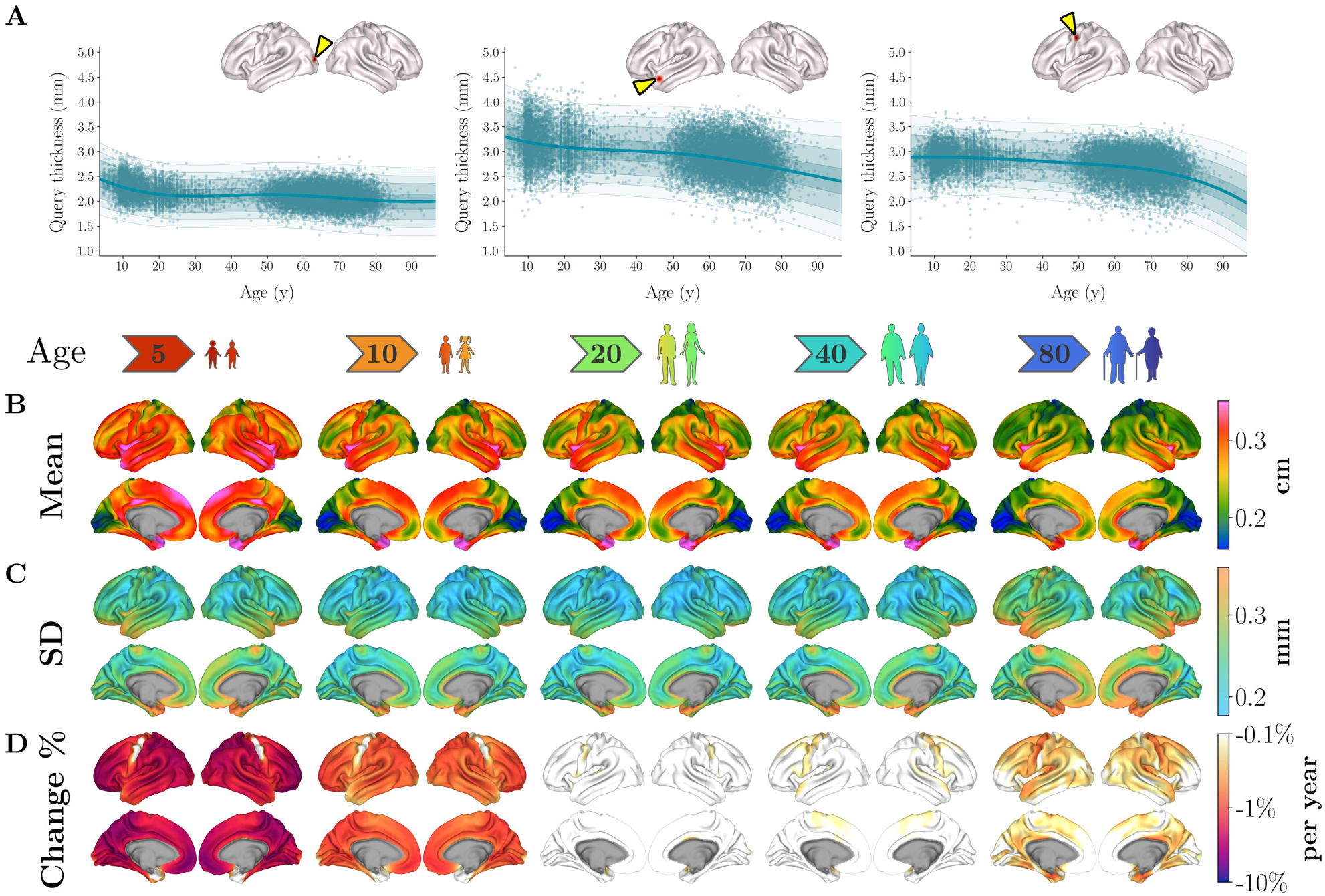
SNM-Derived High-Resolution Charts of Cortical Thickness across the Lifespan. A single pretrained spectral model enables the extraction of lifespan thickness growth charts for many spatial queries simultaneously. **(A)** Normative trajectories are shown for three example vertices located in occipital, temporal, and precentral cortices. Shaded regions denote the [25–75], [5–95], [1–99], and [0.1–99.9] centile bands, and scatter points represent harmonized individual observations with site effects removed. Extending this procedure to all cortical vertices yields high-resolution maps of cortical thickness development and aging, showing normative patterns of **(B)** mean thickness, **(C)** intersubject variation (distributional spread), and **(D)** percent yearly change at five representative ages (5, 10, 20, 40, and 80 years). The yearly rate of change was computed from the derivative of the median trajectories. All charts were generated using an SNM with 1000 modes.

These charts reveal intricate cortical patterning of thickness across the lifespan (Fig. 3B–D). Mean maps show that regions such as the insula, temporal pole, and middle temporal gyrus remain among the thickest throughout life, whereas occipital cortex consistently exhibits lower thickness. Systematic asymmetries along the central sulcus are also evident, with the precentral gyrus remaining thicker than the postcentral gyrus across all ages, consistent with prior observations ^35^.

Variance maps reveal patterns of interindividual variability across the lifespan; variability remains relatively stable through adolescence and adulthood but increases in later life, indicative of accumulating divergence in aging trajectories. Higher regional variance is consistently observed near the medial wall, including the parahippocampal and cingulate gyri, likely reflecting a combination of true anatomical heterogeneity in limbic regions ^36^ and reduced measurement reliability in these areas.

Percent-change maps delineate two periods of accelerated cortical thinning, during development and in late life, separated by relative stability in young adulthood. The spatial patterns of thinning differ across these phases: regions such as the precentral gyrus and temporal pole show minimal thinning during early development but undergo steep declines in late life. These contrasts highlight distinct neurobiological processes underlying cortical remodeling across development and aging^37^.

### 2.4 Gradients of Lifespan Thickness Growth

To characterize organizational principles in normative thickness development and aging, we applied principal component analysis to vertex-wise normative median thickness trajectories, identifying three orthogonal thickness growth gradients (TGG1–3) that together explained over 90% of variance in vertex-wise growth charts. Fig. 4A shows the spatial organization of each gradient. TGG1 follows the canonical cortical thickness hierarchy, distinguishing comparatively thicker temporal, fronto-medial, and insular regions from comparatively thinner occipital and postcentral areas. TGG2 follows a ventral–dorsal axis, extending from orbitofrontal and inferior temporal cortices at one pole to precentral and dorsolateral prefrontal regions at the other. Finally, TGG3 contrasts occipital cortex, temporal poles, and precentral cortex with medial and orbitofrontal cortices (Fig. 4A).

**Fig. 4.**
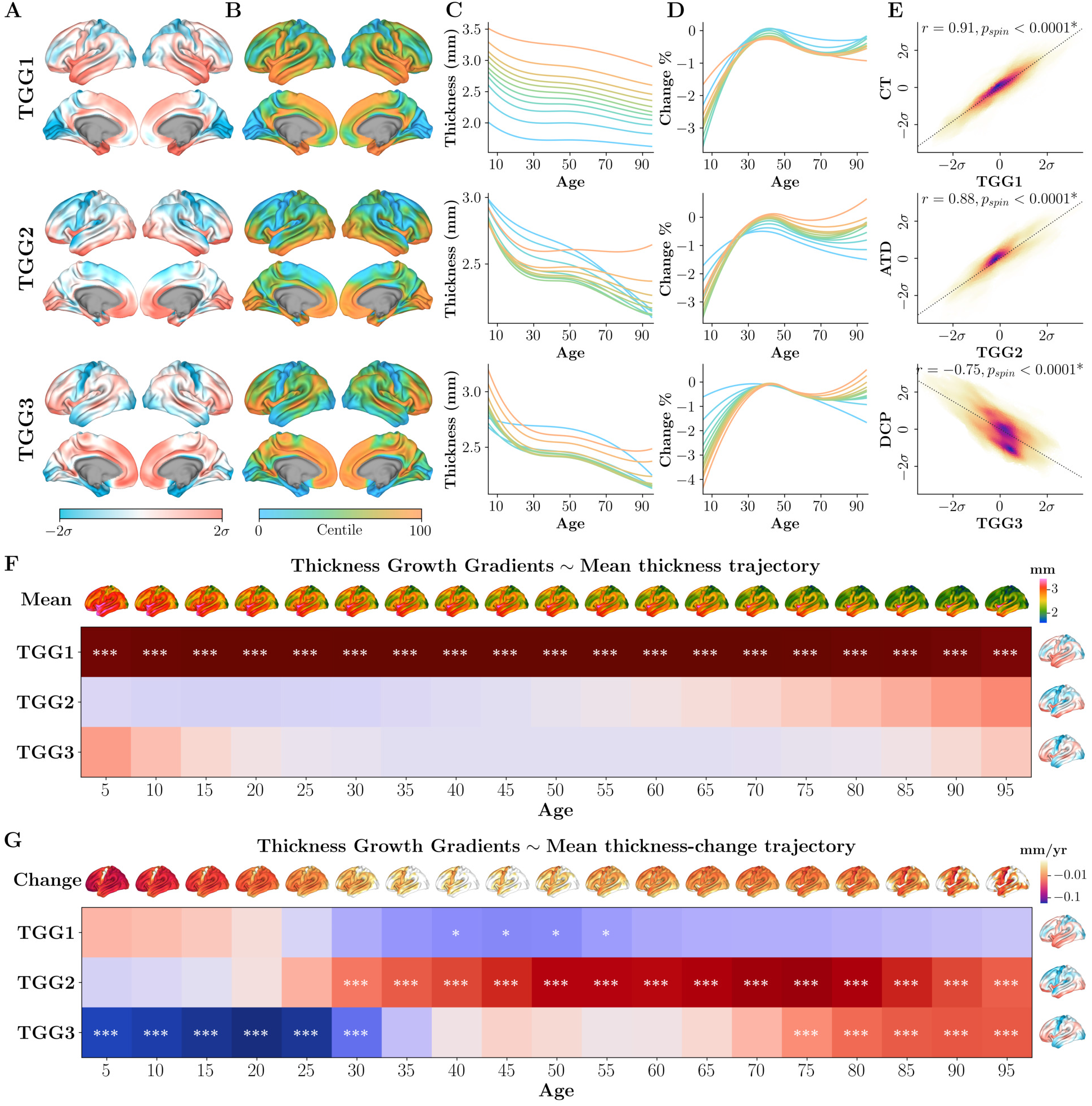
Lifespan Thickness Growth Gradients. SNM-1000 was used to estimate median cortical thickness at one-year increments from ages 5 to 95. **(A)** Principal component decomposition of these estimates yielded three thickness growth gradients. Cortical projections show the spatial distribution of each gradient (*σ*-scaled). **(B)** These maps were converted to centile ranks to delineate strata along gradients. Vertices were stratified into groups along each gradient to define spatial queries, which were used to generate gradient-aligned charts of median thickness **(C)** and percent annual change **(D)**, thereby illustrating the characteristic growth scheme captured by each gradient (see also Extended Data Figs. E.2, E.3, E.4). **(E)** To aid interpretation, gradients are respectively compared against cortical maps of mean thickness (CT), aging thickness decline (ATD), and developmental cortical pruning (DCP); *p*_*spin*_ *<* 0.0001, FDR-adjusted across all 3 tests. **(F)** Correlation of each gradient against the normative mean thickness map at 5-year age increments from 5 to 95, and **(G)** against the corresponding normative thickness-change map (\**p*_*FDR*_ *<* 0.05, \*\*\**p*_*FDR*_ *<* 0.001, FDR-adjusted within each panel). Abbreviations: TGG, Thickness Growth Gradient; CT, Cortical Thickness; ATD, Aging-related Thickness Decline; DCP, Developmental Cortical Pruning.

Cortical vertices were stratified based on their centile rank along each gradient (Fig. 4B, also see Extended Data Figs. E.2, E.3, E.4) to generate representative mean and percent-change trajectories, providing continuous profiles of thickness growth variation along gradients (Fig. 4C,D). This revealed three distinct, interpretable growth schemes. TGG1 captures a stable lifelong thickness hierarchy (Extended Data Fig. E.5), distinguishing regions that remain consistently thick from those that are characteristically thin, effectively reflecting the global pattern of population-wide mean cortical thickness. TGG2 primarily reflects late-life dynamics, with one end encompassing regions that undergo pronounced thinning after mid-adulthood and the other including areas that remain relatively stable in older ages (Extended Data Fig. E.6). TGG3, in contrast, captures early-life developmental effects, separating cortical regions by the rate of thinning during childhood and adolescence (Extended Data Fig. E.7). To validate these interpretations, we compared each gradient against cortical maps reflecting the corresponding growth scheme (Fig. 4E). TGG1 strongly corresponded with an established average cortical thickness map^38^ (CT; *r* = 0.91, *p*_*spin*_ *<* 0.0001), supporting its role as a global normative thickness axis. TGG2 aligned with a map of aging-related thickness decline (ATD; defined as the average percent change between ages 40 and 95; *r* = 0.88, *p*_*spin*_ *<* 0.0001), while TGG3 corresponded with developmental cortical pruning (DCP; average percent change between ages 5 and 20; *r* = −0.75, *p*_*spin*_ *<* 0.0001).

To further evaluate these interpretations, we correlated each gradient with normative mean-thickness and thickness-change maps computed at 5-year age intervals from 5 to 95 years (Fig. 4F,G). TGG1 remained significantly associated with mean thickness throughout the lifespan (all *p*_*FDR*_ *<* 0.001; Fig. 4F). This correspondence was replicated in three non-overlapping cohorts spanning development, young adulthood, and aging (all *r >* 0.88, *p*_*spin*(*FDR*)_ *<* 0.0001; Extended Data Fig. E.8), confirming that TGG1 represents a stable lifelong mean-thickness pattern. As such, the subsequent orthogonal gradients (TGG2–3) inherently represent the principal modes of variation around this stable mean, capturing patterns of cortical thickness change over the lifespan. TGG2 was significantly associated with the rate of thickness change from approximately 30 years of age onward, consistent with an aging-related decline signature. In contrast, TGG3 was significantly associated with the rate of change early in life and, notably, again in advanced age, but with the opposite sign (Fig. 4G). This sign reversal indicates that cortical regions undergoing the greatest thinning during development are among the most resistant to thinning in later life. We further observed a negative association between ATD and DCP (*r* = −0.43, *p*_*spin*_ = 0.017; Extended Data Fig. E.9), reemphasizing the inverted spatial characteristics of cortical reorganization during development and aging. This pattern is consistent with retrogenesis^37,39,40^, whereby developmental and aging-related cortical change follows a shared but inverted spatial axis; notably, this reversal is found to be specifically expressed along TGG3, whereas TGG2 mainly tracks aging-related changes.

We next compared TGG1–3 with established cortical hierarchies spanning cellular, anatomical, functional, genetic, and developmental phenotypes. All three gradients were significantly differentiated across histologically defined laminar cytoarchitectonic classes^41^ (omnibus ANOVA against spatial-autocorrelation-preserving surrogates^42^; TGG1: *F* = 32.4, *p*_*FDR*_ = 0.002; TGG2: *F* = 18.0, *p*_*FDR*_ = 0.018; TGG3: *F* = 25.9, *p*_*FDR*_ = 0.008; Fig. 5A). TGG1 separated idiotypic (primary sensory/motor) from paralimbic cortex (both *p*_*FDR*_ = 0.031), recapitulating the canonical sensorimotor-to-association (SA) hierarchy. TGG3 showed a heteromodal-specific elevation (*p*_*FDR*_ = 0.007), whereas TGG2 exhibited a graded pattern across laminar classes, although no individual class survived post-hoc correction.

**Fig. 5.**
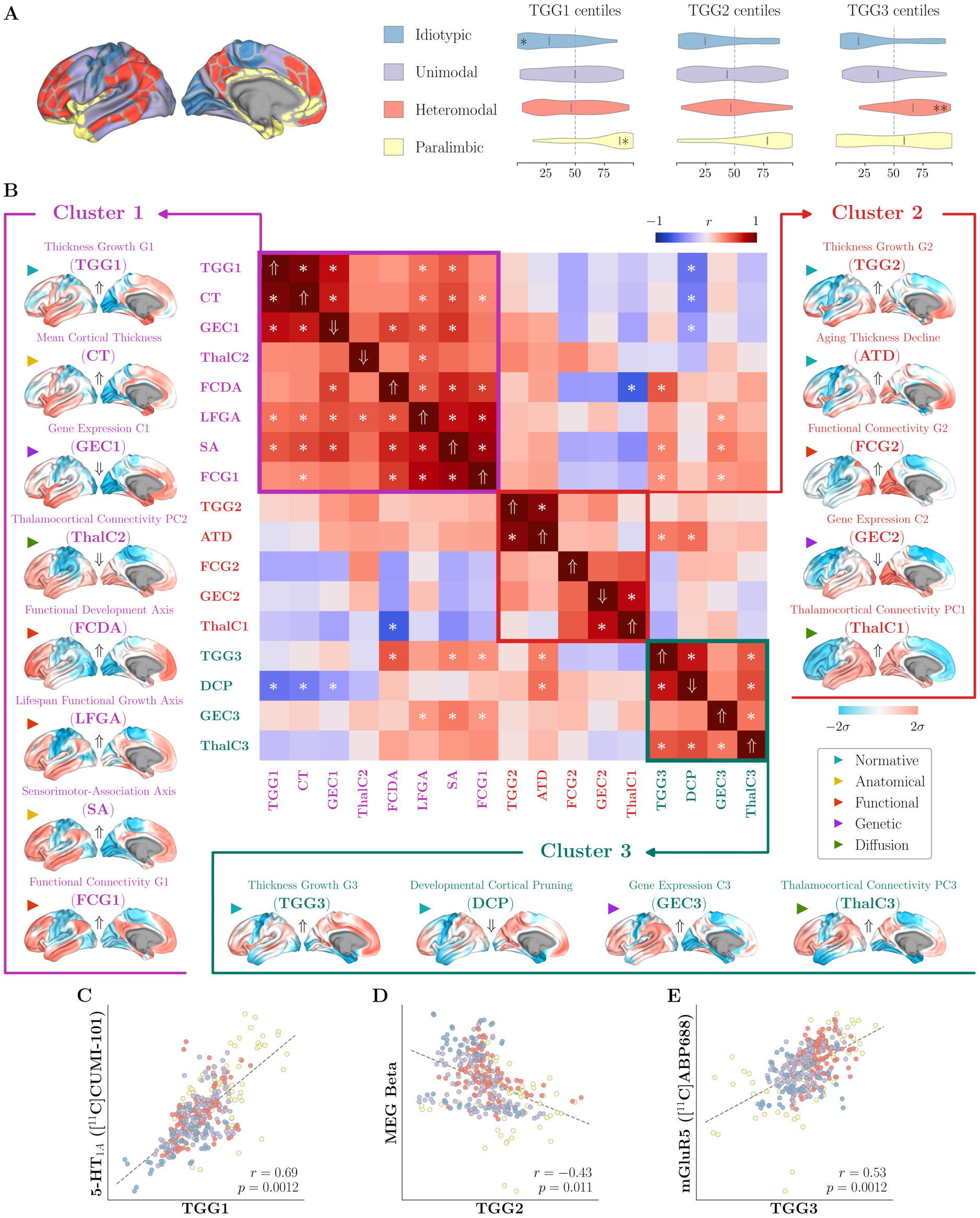
Thickness Growth Gradients Recapitulate Established Cortical Hierarchies. **(A)** Distributions of TGG1–3 centile values across histologically defined laminar cytoarchitectonic classes^41^. Omnibus differentiation was tested per gradient via one-way ANOVA against spatial-autocorrelation-preserving surrogates^42^. All three gradients exhibited significant class-level differentiation after correcting for the three omnibus tests (*p*_*FDR*_ *<* 0.05). Per-class follow-up tests identified classes driving differentiation effects (FDR-adjusted across 12 tests, \**p*_*FDR*_ *<* 0.05, \*\**p*_*FDR*_ *<* 0.01). **(B)** Data-driven clustering of TGG1–3 alongside a set of established anatomical, functional, genetic, and diffusion cortical hierarchies identified three clusters of spatially corresponding maps (C1–C3); heatmap shows pairwise correlations across cortical maps (\**p*_*FDR*_ *<* 0.05, FDR-adjusted across all 136 tests); arrows indicate instances where maps were sign-flipped to maintain positive within-cluster associations (⇑ unflipped, ⇓ flipped). **(C–E)** Exemplary associations between each gradient and independent neuromaps drawn from a comprehensive comparison in Supplementary Fig. S.17: TGG1 vs. 5-HT1A receptor density^51^ **(C)**, TGG2 vs. MEG beta-band power^53^ **(D)**, and TGG3 vs. mGluR5 receptor density^54^ **(E)**; points colored by cytoarchitectonic classes. Abbreviations: TGG, Thickness Growth Gradient; CT, Cortical Thickness; GEC, Gene Expression Component; ThalC, Thalamocortical Connectivity component; FCDA, Functional Development Axis; LFGA, Lifespan Functional Growth Axis; SA, Sensorimotor-Association Axis; FCG, Functional Connectivity Gradient; ATD, Aging-related Thickness Decline; DCP, Developmental Cortical Pruning.

An exploratory clustering analysis over a broad set of anatomical, functional, genetic, and diffusion-derived cortical hierarchies (Fig. 5B) grouped TGG1 with CT^38^, the principal transcriptional component (GEC1)^43,44^, the SA axis^45^, the principal functional connectivity gradient (FCG1)^46^, the second thalamocortical connectivity component (ThalC2)^47^, the functional development axis (FCDA)^48^, and the lifespan functional growth axis (LFGA)^49^. TGG2 clustered with ATD, FCG2^46^, GEC2^43^, and ThalC1^47^, whereas TGG3 clustered with DCP, ThalC3, and GEC3 (Fig. 5B). While some maps aligned predominantly with a single cluster (e.g., GEC1 with C1), others showed partial alignment across clusters (e.g., GEC3 with C1 and C3), highlighting an interplay across cortical hierarchies.

Many associations remained significant under stringent spatial-correspondence testing (two-sided permutation tests, FDR-adjusted across 136 pairwise comparisons, 36 significant pairs with *p*_*FDR*_ *<* 0.05), indicating that the derived normative thickness growth gradients are closely intertwined with many established human cortical hierarchies. An expanded supplementary comparison against 81 cortical maps^50^ reiterates this observation, uncovering 60 significant pairwise associations (*p*_*FDR*_ *<* 0.05 across 243 tests, Supplementary Fig. S.17). Fig. 5C–E shows representative examples from this analysis, where TGG1 aligned with cortical 5-HT1A receptor density^51^ (*r* = 0.69), TGG2 with MEG beta-band power^52,53^ (*r* = −0.43), and TGG3 with mGluR5 receptor density^54^ (*r* = 0.53; Fig. 5C–E).

### 2.5 SNM Recovers Established AD Signatures and Quantifies Individual Atrophy Burden

SNM’s pretrained brain charts can be adapted to new imaging sites using only a small healthy sample, enabling translational potential for normative clinical assessment. To illustrate this, we calibrated SNM-1000 on an independent elderly cohort comprising healthy controls (HC, *N* = 132), individuals with mild cognitive impairment (MCI, *N* = 202), and patients with Alzheimer’s Disease (AD, *N* = 208; see Supplementary Section S.1.4 MACC Clinical Imaging Data). Fine-tuning harmonization parameters via transfer learning to this dataset allowed identification of vertex-wise deviations in cortical thickness relative to the normative reference (Fig. 6A).

**Fig. 6.**
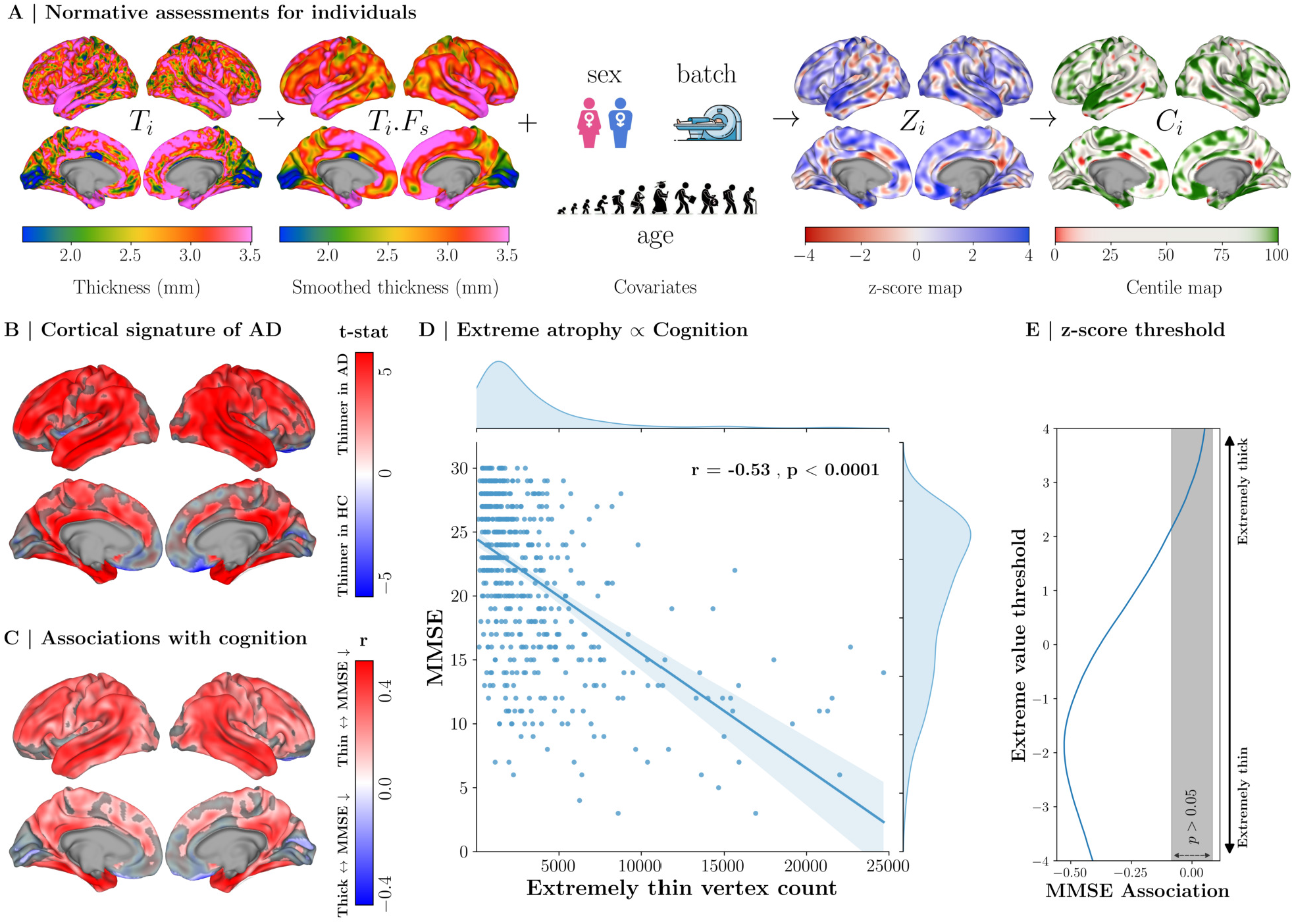
Cortical Signature of Atrophy in Alzheimer’s Disease and Its Cognitive Correlates. Individualized cortical atrophy assessments in AD can be derived from pretrained SNM models, revealing characteristic atrophy patterns and cognitive associations. **(A)** Schematic illustrating SNM-based assessment of individual deviations. Individual cortical thickness maps are smoothed, and vertex-resolution normative deviation maps (z-scores or centiles) are then generated based on the individual’s covariates; cortical projections show an exemplary participant. **(B)** Group-level normative differences between HC and AD. **(C)** Vertex-wise associations between normative deviation z-scores and cognitive performance (MMSE). **(D)** The ETVC metric, an individual normative marker quantifying extreme atrophy, is associated with cognitive performance in the clinical cohort. **(E)** Comparison of z-score thresholds for ETVC reveals that vertices with extreme atrophy show the strongest association with cognitive impairment. Cortical projections highlight significant regions (dimmed for non-significant vertices) at *α* = 5% after FDR correction. Abbreviations: HC, Healthy Cohort; AD, Alzheimer’s Disease; MMSE, Mini-Mental State Examination; ETVC, Extremely Thin Vertex Count; FDR, False Discovery Rate.

Building on prior regional normative AD studies^14,55,56^, SNM’s vertex-resolution z-score maps recover the characteristic cortical signature of AD atrophy^39,57–59^ (Fig. 6B), with pronounced normative thinning across temporal, parietal, and frontal regions, alongside relative preservation of visual and medial frontal areas. These deviations were also associated with cognitive performance, as assessed by Mini-Mental State Examination (MMSE) scores, with greater cognitive impairment linked to stronger cortical thinning, particularly in temporal regions (Fig. 6C). Notably, the spatial pattern of cognitively relevant thinning aligned selectively with the second thickness growth gradient (TGG2; *r* = −0.37, *p*_*FDR*_ = 0.01), but not with TGG1 or TGG3. Consistent with prior work^39^, this correspondence indicates that the topography of cognitively relevant atrophy in AD mirrors patterns of accelerated cortical aging. By recapitulating established group-level patterns of AD atrophy in an independent cohort, these analyses verify the validity of SNM’s vertex-resolution normative assessments in clinical populations, providing a foundation for quantifying cortical abnormalities at the individual level.

Inspired by prior approaches^14,60^, we derive patient-specific abnormality profiles using extreme value statistics. For each individual, the Extremely Thin Vertex Count (ETVC) quantifies the number of vertices with cortical thickness below a normative threshold (e.g., *z <* −1.96), measuring the extent of severe atrophy. ETVC was robustly associated with cognitive impairment across the full sample (*r* = −0.53, *p <* 0.0001), within AD (*r* = −0.34, *p <* 0.0001) and MCI (*r* = −0.20, *p* = 0.004) subgroups (Fig. 6D; Supplementary Section S.1.10 Within-Group Cognitive Associations). Notably, ETVC showed the strongest association with cognitive impairment at more extreme normative thresholds (−3 *< z <* −2; Fig. 6E), suggesting that severe focal cortical abnormalities carry greater cognitive relevance than more modest deviations distributed across the cortex. Supplementary analyses show that ETVC is more strongly associated with cognitive scores than normative mean cortical thickness (Supplementary Section S.1.11 Brain-Wide Cognitive Associations), underscoring the utility of high-resolution brain charts for deriving clinically relevant predictive biomarkers.

### 2.6 SNM Highlights Heterogeneity in Individual Cortical Atrophy

While group-level analyses characterizing shared patterns of cortical deviation provide a population-level description of AD-related atrophy, group averages cannot capture the extent to which individual patients conform to or depart from these shared patterns, potentially masking personalized atrophy profiles that may hold crucial insights into disease mechanisms and inform individually-tailored clinical interventions^1,18^. The same limitation may extend to averaging within putative subtypes if substantial heterogeneity persists among patients^61^, a possibility we examine below. SNM’s vertex-resolution assessments capture individual deviations, enabling detailed exploration of heterogeneity in cortical atrophy among AD patients. As illustrated in Fig. 7A, even individuals with similar demographics and diagnoses can exhibit markedly distinct thinning patterns, with minimal spatial overlap (*r* = −0.02, *p*_*spin*_ = 0.49). Importantly, these deviation maps can be projected onto each individual’s native brain space, in either surface or volumetric format, yielding clinically interpretable atrophy maps that can potentially support personalized evaluation and monitoring.

**Fig. 7.**
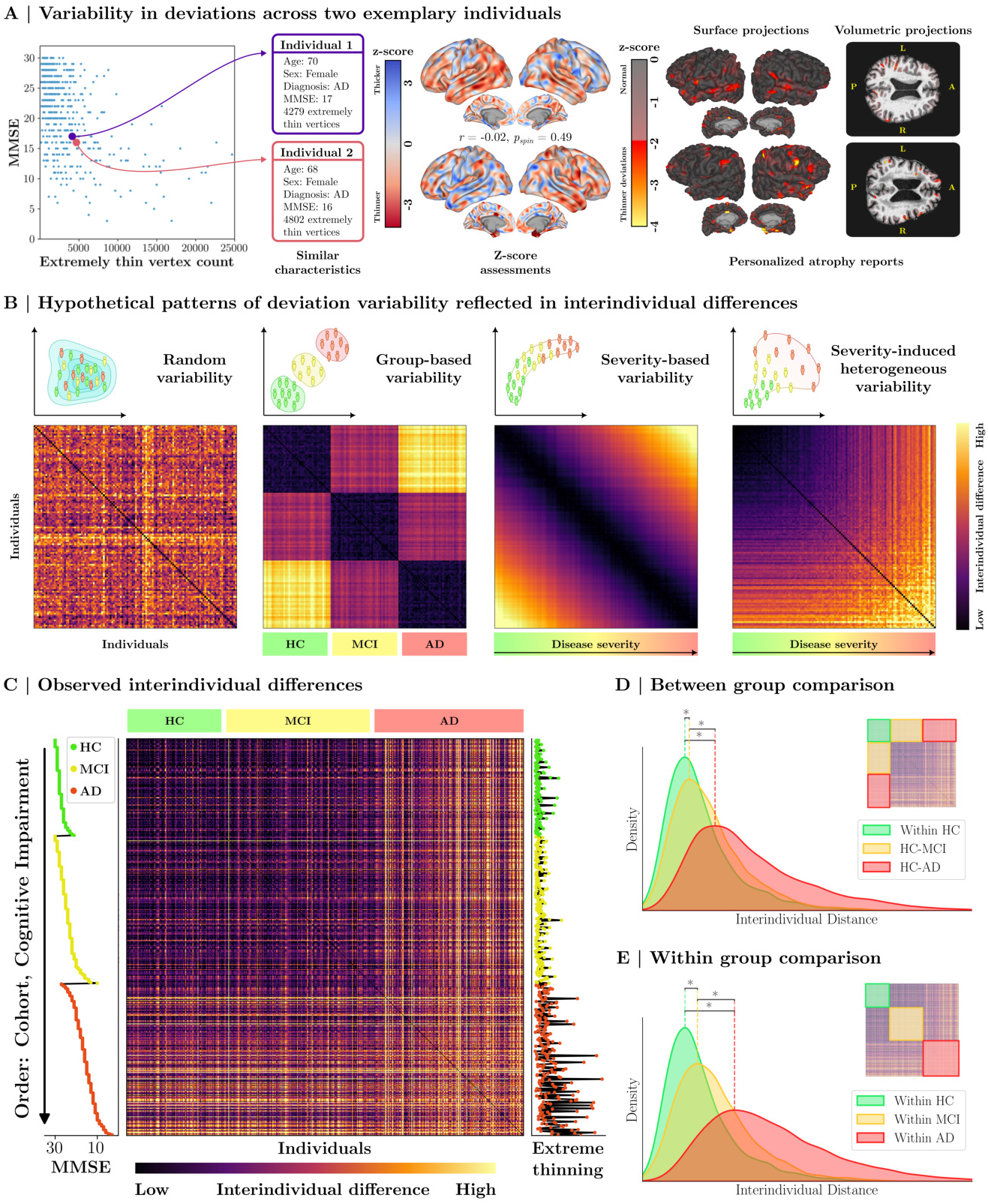
Interindividual variability in AD atrophy. Individualized deviation maps were used to characterize heterogeneity in AD-related cortical atrophy. **(A)** Two exemplary individuals with similar age, sex, cognitive scores, and diagnosis who exhibit markedly different cortical atrophy patterns. **(B)** Interindividual differences are quantified via Euclidean distances between maps; schematic examples illustrate four hypothetical deviation mechanisms: random deviations unrelated to pathology, diagnosis-aligned deviations, uniformly progressive deviations along disease stages, and severity-induced heterogeneous divergence from norms. **(C)** The empirical interindividual distance matrix, sorted by diagnosis and, within each diagnosis, by decreasing cognitive performance (MMSE, left), reveals heightened heterogeneity in deviations of AD-diagnosed individuals; per-individual ETVC is shown on the right for reference. **(D–E)** Distributions of empirical interindividual distances across cohorts. **(D)** Between-group comparisons relative to HC show progressively increasing deviations in MCI and AD cohorts. **(E)** Within-group heterogeneity significantly increases from HC to MCI and AD. (∗: Welch’s t-tests, FDR-corrected, *p <* 0.0001)

Differences in atrophy patterns may reflect distinct mechanisms through which pathology associates with normative deviations. To illustrate this, we simulated four hypothetical scenarios (Fig. 7B), each presenting differences across 60 individual normative deviation maps. Pairwise differences were quantified by Euclidean distance to measure interindividual variability. In the first scenario, deviations occur in a random, non-systematic manner. In the second scenario, deviations are tightly linked to diagnostic categories. In the third scenario, deviations are linked to disease severity, with individuals of similar severity exhibiting similar atrophy patterns. In the final scenario, increasing severity produces progressively greater heterogeneity in deviations.

We next compute the empirical interindividual difference matrix (Fig. 7C) and sort it by diagnostic group (HC, MCI, and AD) and, within each group, by cognitive performance (as a marker of symptom severity). The resulting matrix reveals progressively greater differences in cortical deviation patterns from HC to MCI and AD, with particularly pronounced interindividual variability among patients with AD. Relative to HC, both MCI and AD exhibited significantly greater differences in deviation patterns, with larger effects in AD (Welch’s two-sample t-tests: HC–HC vs. HC–MCI, *t* = 19.1, *p <* 0.0001, *d* = 0.23; HC–HC vs. HC–AD, *t* = 95.9, *p <* 0.0001, *d* = 0.91; HC–MCI vs. HC–AD, *t* = 97.46, *p <* 0.0001, *d* = 0.83; Fig. 7D). Within-group comparisons likewise showed increasing heterogeneity from HC to MCI to AD (HC vs. MCI, *t* = 31.4, *p <* 0.0001, *d* = 0.39; HC vs. AD, *t* = 121.8, *p <* 0.0001, *d* = 1.23; MCI vs. AD, *t* = 107.6, *p <* 0.0001, *d* = 1.04; Fig. 7E). These findings suggest that AD is associated with increasingly divergent cortical atrophy patterns across individuals, highlighting the heterogeneous nature of individual atrophy in AD.

### 2.7 Individual Atrophy Patterns Form a Continuous Heterogeneity Landscape

To further elucidate the heterogeneity in deviation patterns, we embed the high-dimensional normative assessments into a lower-dimensional latent space that preserves interindividual differences (Fig. 8A). Within this landscape, healthy individuals form a dense central cluster, consistent with relatively homogeneous deviation maps, whereas individuals with MCI show greater dispersion and those with AD exhibit the greatest departure from the healthy cluster (Fig. 8B). Notably, individuals with AD diverge in multiple directions, indicating heterogeneity not only in the magnitude but also in the spatial pattern of normative deviations. Averaging deviation maps within local neighborhoods of the landscape (cortical projections in Fig. 8A, bottom) further reveals distinct patterns ranging from negligible (left) to localized (middle) and widespread (right) atrophy (see Supplementary Section S.1.13 Heterogeneity Landscape Subgroups). Exemplary cortical projections in Fig. 8A (right) further illustrate that individuals with AD vary in both the severity and spatial organization of cortical atrophy. Supplementary analyses confirm that this heterogeneity landscape is robust across dimensionality reduction methods and parameter settings (see Supplementary Section S.1.14 Robustness of the Heterogeneity Landscape).

**Fig. 8.**
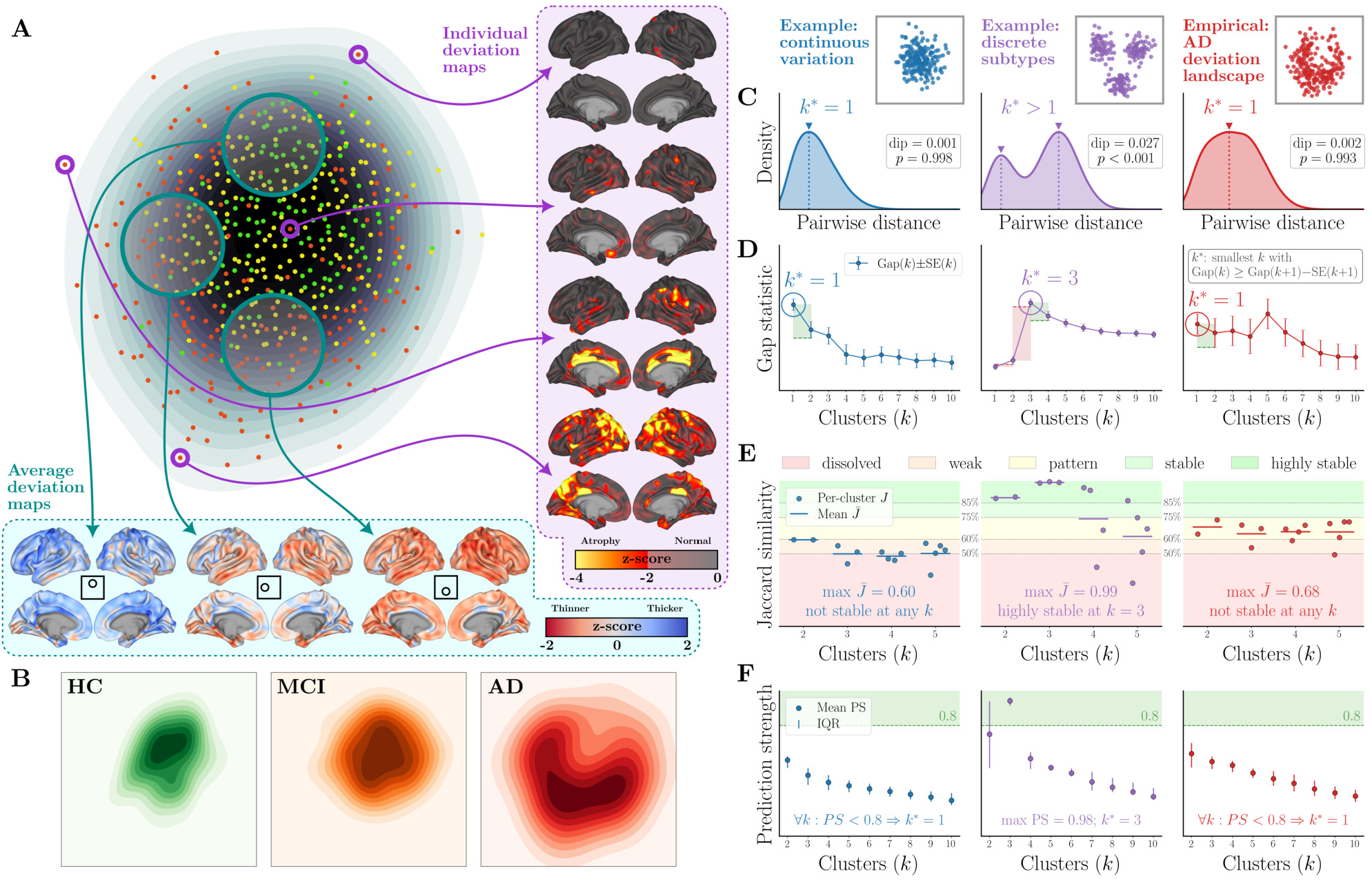
Heterogeneity Landscape of Atrophy in AD. **(A)** Individual deviation maps are embedded into a 2D landscape preserving interindividual differences, with HC (green), MCI (yellow), and AD (red) cohorts occupying partially overlapping regions. Average deviation maps within selected subregions of the landscape (bottom) and exemplary individual AD maps (right) are shown. **(B)** Density plots highlight regions predominantly occupied by each cohort. **(C–F)** Four complementary tests assess whether the empirical AD deviation maps (red, right) are organized into discrete subtypes. Each test is shown alongside two reference cases of matched sample size and covariance: one comprising three well-separated subtypes (purple, middle) and one comprising continuous variation without discrete subtypes (blue, left). **(C)** Unimodality of the distribution of interindividual distances, assessed via Hartigan’s dip statistic^62^; discrete subtypes produce a multimodal distribution, whereas continuous variation does not. **(D)** Cluster tendency across candidate partitions (*k* = 1–10), assessed via the gap statistic^63^; *k*^∗^ denotes the selected number of clusters, where *k*^∗^ = 1 indicates that no partition is favored over a single cluster. **(E)** Stability of candidate partitions (*k* = 2–5) under bootstrap resampling, quantified by the Jaccard similarity of recovered clusters^64^; shaded bands denote conventional interpretation thresholds, with stable partitions requiring all constituent clusters to exceed *J* = 0.75. Horizontal lines indicate the mean across clusters. **(F)** Generalizability of candidate partitions across independent subsamples, quantified by prediction strength^65^; partitions are considered generalizable above the recommended threshold of 0.8. Across all four tests, the empirical deviation maps showed no evidence of discrete subtype structure. Additional detail and replication across alternative dimensional representations are reported in Supplementary Section S.1.15 Clusterability of AD Deviation Patterns.

We next quantitatively assessed whether the apparent continuity of these individual differences reflects an absence of discrete cluster structure in the underlying deviation maps. Because clustering algorithms return a partition even when none is warranted, we first tested whether AD deviation maps exhibit evidence of discrete cluster structure. Results were reported alongside simulated examples of discrete and continuous variation, matched for sample size and covariance (Fig. 8C–F). Hartigan’s dip statistic^62,66^ applied to pairwise differences revealed strongly unimodal distributions in the full sample (Dip = 0.0005, *p* = 0.995) and within AD (Dip = 0.0014, *p* = 0.996), with no evidence of distinct modes corresponding to diagnostic groups or AD subtypes (Fig. 8C). Assessments of cluster tendency^63^, stability under resampling^64^, and generalizability across independent subsamples^65^ converged on the same conclusion: the gap statistic favored a single-cluster solution, while no candidate multi-cluster partition met conventional criteria for stability or generalizability (Fig. 8D–F). Consistent with these results, a formal test of hierarchical cluster splits (SigClust2^67^) identified no significant partition of the AD cohort. Sensitivity analyses confirmed that these findings held across alternative representations of the deviation maps (Supplementary Section S.1.15 Clusterability of AD Deviation Patterns). Together, these analyses suggest that cortical atrophy in AD varies predominantly along a continuum rather than across discrete subtypes. This highlights how group- or subtype-level summaries can obscure individual variation in cortical atrophy and motivates approaches such as vertex-resolution normative assessment that explicitly characterize personalized deviation patterns.

## 3 Discussion

In this manuscript, we introduce Spectral Normative Modeling (SNM), which builds on prior normative approaches by leveraging eigenmode representations as a compact basis for normative inference. Operating in the spectral domain enables efficient generation of normative brain charts across multiple spatial scales, including vertex resolution. This, in turn, allows growth trajectories to be efficiently examined at high resolution, revealing principal organizational hierarchies that capture distinct lifelong, maturational, and aging processes. This further supports individualized, high-resolution assessments of cortical anomalies, providing a potential pathway toward translating normative modeling into precision clinical applications. To facilitate broader adoption, we release the pretrained models alongside a curated software package to facilitate application to new datasets and alternative clinical cohorts.

### 3.1 Lifespan Gradients of Cortical Growth

A key advance enabled by SNM is its ability to generate normative estimates for novel, *a posteriori*-defined regional delineations without retraining the model. Combined with the computational efficiency afforded by spectral representations, which substantially reduce complexity when concurrently generating tens to hundreds of thousands of normative ranges across queries, SNM facilitates normative mapping at higher resolutions. This capability enabled the characterization of principal gradients of cortical growth across the lifespan. Moreover, by charting normative ranges along gradient strata, SNM provides an interpretable framework for probing neuroanatomical growth schemes along cortical hierarchies.

Our results provide a new normative perspective on the brain’s macroscopic anatomical organization across the lifespan. We show that cortical thickness is structured along three distinguishable growth gradients. The first gradient (TGG1) captures a stable cortical thickness hierarchy whose relative ordering is maintained across lifespan. This hierarchy is anchored in cortical microstructure, being significantly differentiated across laminar histology classes^41^ and aligning with intracortical myelination^38^. This situates TGG1 along an established cortical microstructural organizational axis^68^ and suggests that population-average cortical thickness reflects a conserved program of laminar differentiation that is not tied to any particular age^45^.

Consistent with this interpretation, TGG1 further aligns with the sensorimotor-association axis^45^, the principal functional connectivity gradient^46^, the principal transcriptional hierarchy^43,44^, and the recently described lifespan functional growth axis^49^. At the molecular level, thicker cortical regions along TGG1 show higher densities of multiple neuromodulatory receptor and transporter systems^50,54^, alongside lower myelination^38^ and lower oxidative and glucose metabolism^69^. Together, these correspondences suggest that TGG1 reflects a shared cytoarchitectonic and transcriptional scaffold shaping the principal axes of molecular, structural, and functional cortical organization. Thickness variation around this stable hierarchy resolves into two higher-order gradients (TGG2 and TGG3): one primarily reflecting late-life cortical thinning, the other capturing developmental cortical pruning that reverses in late life.

TGG2 aligns with the second functional connectivity gradient^46^, in that both differentiate primary visual from primary motor areas, revealing that visual cortices are relatively resilient to late-life atrophy while precentral motor areas show greater thinning. This organization is mirrored by the cortical distribution of beta-band oscillatory power, implicated in both motor control and cognition^70^. Consistently, TGG2 aligns with the second transcriptional component, enriched in metabolic processes^43^, the primary cognitive-affective axis^71–73^, and, in our own data, the cortical map linking AD atrophy to cognitive impairments (Section 2.5 SNM Recovers Established AD Signatures and Quantifies Individual Atrophy Burden). Together, these findings implicate metabolic demand as a key determinant linking cortical organization to cognitive vulnerability during aging^40,74^.

TGG3 co-localizes with the transcriptional component most associated with synaptic plasticity and learning^43^, reinforcing its interpretation as a developmental pruning axis^75^. Independent molecular evidence further supports this assignment: TGG3 aligns with metabotropic glutamate receptor density^54,76,77^, GABA_A_ receptor density^78,79^, presynaptic vesicle protein density, and a diverse set of neuromodulatory receptor systems implicated in cortical plasticity^51,54^. Consistent with this molecular profile, TGG3 preferentially involves the association cortex^45^, distinguishing heteromodal laminar areas and aligning with the association-dominant third thalamocortical connectivity component reconstructed from perinatal tractography^47^ as well as the functional development axis^48^. Together, these convergent molecular and anatomical correspondences identify TGG3 as a cortical axis implicated in developmental synaptic organization, underpinned by excitatory and inhibitory neurotransmission.

Notably, the observed anticorrelation between developmental and aging thinning processes (DCP, ATD) demonstrates that regions most stable during development are among those most susceptible to atrophy later in life, providing a nuanced extension of the retrogenesis hypothesis^37,39,40,80^ to normative brain charts. Retrogenesis, originally proposed to describe the reversal of developmental order in aging^80^, has since been supported by evidence spanning cytoarchitectonic^81^, cellular^40^, genetic^82^, and longitudinal imaging studies^83^. Our findings provide a spatially resolved normative perspective on this phenomenon by showing that the developmental–aging reversal is organized along TGG3, separating regions that are developmentally stable but vulnerable to late-life thinning from those that undergo rapid developmental pruning yet remain relatively stable in aging.

### 3.2 Accessibility Benefits

The latent spectral formulation of SNM enables a single, pretrained model to generate normative ranges for a wide variety of queries, including high-resolution queries on different cortical surface templates, regional queries on different cortical surface parcellations^38,84–92^, or custom probabilistic regional delineations such as strata along gradients, all without requiring access to the original imaging data used for training. This is particularly valuable because the large-scale datasets used to train population-wide normative models are costly and time-consuming to acquire^93–96^, restricting access for researchers interested in novel queries. SNM thus overcomes these key barriers to adoption. To support wider use, we provide the pretrained SNM-1000 model alongside a software package for application to new datasets and phenotypes. For clinical scans acquired on previously unseen scanners, local calibration using a small set of healthy reference scans, commonly available in most clinics (for example, scans acquired to rule out pathology), is sufficient for reliable inference without compromising data privacy.

### 3.3 Clinical Applications in AD

Neurodegenerative mechanisms, such as amyloid-*β* plaque deposition and tau-related neurofibrillary tangles, are known to be associated with cortical atrophy in AD^97–100^. As signs of cortical atrophy can appear up to a decade before clinical symptoms^101,102^, high-resolution normative assessments can facilitate early detection and characterization of neurodegeneration. In this study, the pretrained SNM model was used to examine the spatial distribution of cortical atrophy in a clinical AD cohort, demonstrating how insights derived from a large healthy reference sample can be extended to smaller clinical datasets^103,104^, and providing imaging biomarkers with potential applications in AD research^105^.

Our analysis recapitulated hallmark group-level AD cortical atrophy patterns, with strong effects centered on the temporal lobe (temporal pole, superior, middle, and inferior temporal gyri)^57–59,100,106–108^, and extending to distributed neocortical regions including the supramarginal and angular gyri^57–59,107^, superior parietal lobule, frontal gyri, posterior cingulate cortex^59,108–110^, and the precuneus^57–59,100,110^.

We also observed less commonly reported anomalies, such as higher cortical thickness in selected visual and orbitofrontal regions^111^, potentially reflecting compensatory neurobiological mechanisms^112^. Importantly, AD-related atrophy preferentially aligned with the thickness growth gradient associated with aging-related cortical reorganization (TGG2), consistent with the notion of premature aging: regions most susceptible to thinning during healthy aging exhibit the most pronounced atrophy in AD^39,113,114^.

By assessing the occurrence of extreme deviations^60^ across thresholds, ETVC analyses demonstrate that the *focal severity* of cortical thinning, rather than its *spatial extent*, is more closely associated with cognitive impairment in AD. Regions exhibiting extreme localized deviations from normative thickness contribute disproportionately to cognitive decline, whereas more widespread but moderate thinning is less informative. Consequently, comparable overall atrophy burdens can have distinct cognitive consequences depending on whether degeneration manifests as localized, severe loss or diffuse, modest reductions. These findings underscore how high-resolution normative frameworks such as SNM capture cognitively relevant neurodegenerative patterns that are obscured by global atrophy metrics, enabling the detection of early biomarkers before degeneration becomes widespread.

Our evaluations further emphasize the pronounced heterogeneity in individual cortical atrophy patterns in AD, revealing diverse, pathology-driven deviation profiles that are often obscured by group-level analyses^115^. Such heterogeneity may reflect the influence of comorbidities and interacting disease processes^107,116–119^, and has traditionally been addressed using clustering approaches that partition patients into putative atrophy subtypes^59,108,120–124^. However, building on recent regional studies of AD heterogeneity^56,125,126^, our findings indicate that AD-related cortical deviations show no statistically supported cluster structure. Complementary assessments of distributional unimodality^62,66^, cluster tendency^63^, stability^64^, predictive validity^65^, and statistical significance^67^ consistently provided no support for discrete subtypes, suggesting instead that these deviation patterns are better characterized as a continuous, multidimensional spectrum. This conclusion pertains specifically to vertex-wise cortical thickness deviations in the present cohort and does not preclude discrete structure in other phenotypes, including molecular biomarkers or regional volumetric measures, for which subtypes have been proposed^123,124^. In this setting, imposing subtype boundaries on continuous variation risks obscuring clinically meaningful individual differences and can lead to unstable or imprecise characterizations^61^.

Our landscape did not exhibit clear boundaries that would justify subtype separation. Instead, individuals occupied different localities along a continuum: some AD-diagnosed participants showed minimal or negligible atrophy, echoing reports of “minimal-atrophy” subtypes^59,99,108,123^, whereas some HC/MCI individuals exhibited marked atrophy without severe cognitive impairment, potentially reflecting resilience mechanisms^127,128^. This continuity also carries a methodological implication: clustering can produce apparent subtypes even when the underlying variation is continuous. Common internal validity indices select among candidate partitions without testing whether partitioning is warranted in the first place (see Fig. 8C–F). We therefore suggest that subtype claims be accompanied by convergent evidence that the inferred structure is stable, generalizable, and statistically supported, as elaborated in Supplementary Section S.1.15 Clusterability of AD Deviation Patterns. Moreover, when substantial heterogeneity persists within inferred subtypes, subtype averages can obscure the same individual-level variation that such stratification is intended to resolve. These observations favor characterizing cortical neurodegenerative heterogeneity along continuous dimensions^129^, particularly in the absence of convergent evidence for discrete structure.

While these findings highlight the potential clinical utility of SNM for early detection and characterization of atrophy patterns, they remain preliminary, and dedicated application-focused studies will be needed to establish its diagnostic and prognostic value. SNM has potential to support future investigations of differential neurodegenerative patterns across dementia syndromes, including Alzheimer’s disease, vascular dementia, Lewy body dementia, and frontotemporal dementia^130^. More broadly, this framework is applicable to other neurological conditions exhibiting localized abnormalities, such as stroke or traumatic brain injury, where deviations could be examined in relation to functional outcomes and recovery trajectories. Finally, SNM’s sensitivity to subtle, localized atrophy may facilitate the detection of early biomarkers, such as microinfarcts, enabling identification of at-risk individuals and opening opportunities for preventive intervention before widespread pathology emerges^131^.

### 3.4 Contextualizing SNM in Relation to Deep Learning Approaches

Given the recent emergence of deep learning (DL)–based normative modeling frameworks, often based on variational autoencoders^132–136^, it is important to situate SNM within this broader methodological landscape. At a conceptual level, both DL-based models and SNM encode imaging data into a latent space and perform multivariate normative estimation. However, the underlying architectural trade-offs differ substantially. Existing DL-based normative models typically employ deep, nonlinear encoders operating on fixed, low-dimensional regional phenotypes ^132–136^. In contrast, SNM operates directly in vertex space as a wide, single-layer linear encoder defined by fixed eigenmodes rather than trainable weights. This architectural choice enables closed-form normative inference for arbitrary spatial queries, but comes at the cost of reduced capacity to model complex nonlinear relationships. These complementary strengths motivate future development of deep spectral models that integrate eigenmode-based representations with nonlinear learning to achieve both spatial flexibility and increased modeling capacity.

### 3.5 Future Directions and Limitations

The present study introduces SNM as a flexible framework with scope for continued expansion. Beyond our provided thickness charts, future efforts could extend SNM to additional phenotypes and establish a comprehensive library of spectral normative brain charts. Voxel-wise eigenmodes^137^ could extend SNM to whole-brain volumetric phenotypes. Similarly, spectral norms from other modalities, such as volumetric morphometry^13^, white matter microstructure^60,138,139^, functional organization^140,141^, or PET-derived metabolic indices^142^, could provide complementary insights. Together, these extensions would contribute to a deeper, multifaceted understanding of neurotypical brain growth.

Several limitations warrant consideration. First, SNM currently assumes Gaussianity, which may limit applicability to phenotypes with skewness or heavy tails; although beyond the scope of this manuscript, likelihood-warping extensions^16,139^ could address this limitation. Second, SNM presumes spatial queries lie within a low-pass spectral regime captured by a finite set of eigenmodes. Applications should verify this assumption via reconstruction tests of spatial queries (similar to evaluations in Extended Data Fig. E.1). Finally, although we used random-walk Laplacian eigenmodes, SNM is agnostic to the choice of basis. The software supports alternative bases (e.g., derived from geometry^143^, diffusion^25,27^, and functional^46,144^) and shift operators (combinatorial, normalized, or fractional Laplacians^145,146^), allowing adaptation across phenotypes, modalities, or spatial priors.

## 4 Methods

In the ensuing sections, we first describe the brain imaging datasets utilized in this work. We provide a formal description of the high-resolution cortical thickness phenotypes used as input to the normative modeling framework. Next, we introduce the mathematical foundations of SNM, covering core concepts from graph signal processing and normative modeling. Finally, we describe experiments undertaken to evaluate SNM’s performance and suitability.

### 4.1 Brain Imaging Data

This study leverages a large multi-cohort biobank comprising *N*_*p*_=78,405 healthy participants (after quality control and exclusion criteria) aggregated across 30 publicly available neuroimaging datasets^52,147–180^. Several datasets include multiple acquisition sites and/or scanners, yielding 189 distinct site/scanner combinations, which were modeled as a categorical site covariate in all normative estimations. Participants spanned ages 4–96 with 50.6% female representation. In addition to the healthy datasets used to train the normative model, we evaluated the model’s translational capabilities to real-world data from memory clinics (Memory, Ageing & Cognition Centre at the National University of Singapore: MACC)^181,182^. This data comprised 542 samples (ages 50–91, 61.4% female), including 132 cognitively healthy individuals (24.3%), 202 individuals with mild cognitive impairment (MCI; 37.3%), and 208 individuals diagnosed with Alzheimer’s disease (38.4%) based on DSM-IV criteria^182^. While specific datasets differ in acquisition characteristics, all T1-weighted (T1w) structural MRI data underwent cortical thickness extraction using FreeSurfer^183^, with additional details regarding acquisition and preprocessing procedures for each dataset documented in the Supplementary Information (see Section S.1.1 Healthy Lifespan Imaging Data).

### 4.3 Train Test Split

Healthy individuals were stratified into train and test splits to enable validation of the fitted models on unseen data. Namely, 80% (*N*_train_=62,724 individuals) of the healthy data were used to train the model, and the remaining 20% (*N*_test_=15,681) were used for out-of-sample validation (*N*_*p*_ = *N*_train_ + *N*_test_). A single split fold was used, with a randomized approach that controlled for covariate distributions of age, sex, and dataset by stratification; this ensured that the covariates had similar distributions across splits^184^. Supplementary Fig. S.2 provides a visual summary of covariate distributions before and after splitting.

### 4.3 Cortical Thickness

For all cohorts, vertex-wise cortical thickness estimates were derived from preprocessed data. To ensure consistent surface geometry across subjects, an adaptive area–preserving barycentric transformation^185^ was used to resample thickness measurements from native FreeSurfer surfaces^186^ onto the fs-LR 32k template. This template surface uses 32,492 vertices to model each hemisphere, resulting in an average inter-vertex distance of approximately 2 mm; it includes a total of 59,412 vertices after the exclusion of the medial wall. Vertices are aligned between the left and right hemispheres, enabling inter-hemispheric comparison)^52^.

In this space, cortical thickness is represented as a high-resolution vector 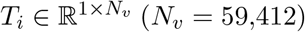, providing a thickness estimate for each vertex of the *i*th individual’s cortical surface. The collection of these high-resolution thickness features across all participants forms the complete high-resolution thickness data sample 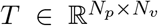 which is used to estimate and evaluate normative ranges of thickness variation across the human lifespan. These data are divided into a training sample 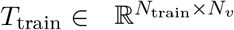 used to construct the normative model and an independent test sample 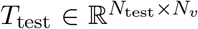 used to evaluate the normative model’s goodness of fit.

### 4.4 Normative Covariates

In normative modeling, the aim is to provide a statistical model that describes the distribution of a variable of interest *y* with respect to a set of covariates *C* (fixed effects), while potentially accounting for batch effect *Z* (random effects)^187^. For simplicity, we consider the covariates of age and sex, although other covariates can be straightforwardly incorporated. Moreover, batch effects were included in the model to facilitate harmonization among the three imaging cohorts (development, young adult, and aging).

### 4.5 Clinical Fine-Tuning

The normative model, pre-trained on *N*_train_ healthy individuals, was subsequently fine-tuned for the clinical sample of *N*_*MACC*_ = 542 individuals using transfer learning. During fine-tuning, the model parameters related to age and sex were kept frozen to maintain consistency with the initial healthy cohort model. However, site parameters for dataset harmonization were fine-tuned on a subsample of 66 healthy individuals (50% of the HC data) from the MACC clinical cohort. The fine-tuned model was then applied to the rest of the clinical sample to generate individualized, high-resolution normative deviation maps. These maps were utilized in normative evaluations of the clinical cohort.

### 4.6 High-Resolution Connectomes

Our proposed approach uses brain connectivity eigenmodes computed from a reference brain connectivity structure^23^. Connectivity eigenmodes were selected for their ability to span both cortical hemispheres, enabling a cohesive representation of interhemispheric relationships. In contrast, geometric modes analyze hemispheres separately, overlooking interhemispheric interactions. Additionally, as many brain disorders with structural deviations from normative patterns are hypothesized to express spatial signatures aligned with anatomical connectivity, connectome eigenmodes are optimally suited to capture such abnormalities^28–33^. High-resolution structural connectivity data were utilized for this purpose^188^.

To this end, we sourced connectomes computed from a tractography pipeline detailed elsewhere^189^. In brief, diffusion-weighted imaging data were used to estimate structural connectivity. Probabilistic tractography was conducted using MRtrix3 to reconstruct whole-brain tractograms (5 million streamlines, with anatomically constrained tractography). Tractography endpoints were used to construct connectomes encoding streamline count at the resolution of the fs-LR 32k template (same space as the thickness information, with *N*_*v*_ = 59,412 vertices representing network nodes). Individual connectomes were combined to form an estimate of group-level connectivity. Connectome spatial smoothing was performed to account for endpoint inaccuracies and improve connectome reliability^190,191^. This yielded a high-resolution weighted adjacency matrix 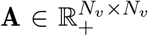 in which element *A*_*i,j*_ denotes the group average strength of structural connectivity between nodes *i* and *j*. The eigenmodes resulting from this high-resolution connectome mapping pipeline were previously tested to assess their accuracy in encoding brain signals^23^.

### 4.7 Brain Signal Reconstruction

Graph signal processing enables the study of data encoded on a graph/network structure^192^. As such, considering that the human brain is fundamentally a network structure, graph signal processing can be utilized to design new ways to analyze and study the brain^193^. Here, we utilize dimensionality reduction and signal reconstruction techniques that encode cortical information in a lower-dimensional latent space^194,195^. To this end, we use high-resolution connectomes as a weighted graph *G* : (*V*, A) where *V* = {1, 2, · · ·, *N*_*v*_} is the set of high-resolution cortical nodes (vertices) and 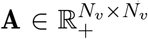 is a weighted (non-negative) adjacency matrix. We utilize GSP to model any brain signal 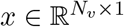 that is defined on the set of vertices *V*. In this context, we model vertex-wise cortical thickness estimates as a brain signal.

The random-walk Laplacian matrix *L*_*rw*_ = *I* − *D*^−1^A is used as the graph shift operator (where 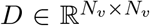 denotes the diagonal strength matrix). This shift operator is diagonalized via singular value decomposition *L*_*rw*_ = ΨΛΨ^−1^ to compute an orthogonal basis for information reconstruction. Here, 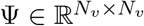 denotes the full set of eigenvectors (also called eigenmodes) corresponding to the left singular vectors of *L*_*rw*_, and 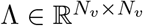 denotes the diagonal matrix of eigenvalues (singular values) associated with the eigenmodes such that Λ_*i,i*_ = *λ*_*i*_ is the *i*th smallest eigenvalue and 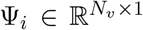 (*i*th column of Ψ) refers to its associated eigenmode. The graph Fourier transform (GFT) encodes *x* into the graph spectral domain 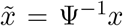, where 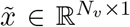 denotes the encoded signal, and 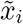 is the *i*th element of 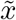 quantifying the signal loading on the *i*th eigenmode 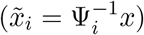. Conceptually, this encoding transforms the brain signal *x* defined in the spatial domain to a latent embedding 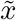 in graph spectral domain. The encoded signal in the graph spectral domain can be mapped back to the spatial domain via the inverse GFT: 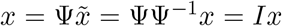. Notably, this transformation, utilizing the full set of eigenmodes, yields an exact reconstruction of any brain signal. Our goal is to use only a fraction of Ψ to derive a low-dimensional representation of brain signals. We selected the random-walk Laplacian shift operator as it yields eigenmodes encoding topological frequency such that eigenmodes associated with smaller eigenvalues capture signals that vary more smoothly along network edges^196^.

Deriving the full set of eigenmodes (Ψ_1_ to 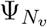) is computationally demanding; however, graph Fourier filtering enables efficient approximation of brain signals using a filter that only requires a limited set of eigenmodes. Specifically, we can define a diagonal filtering matrix 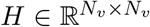, where *H*_*i,i*_ = *h*(*λ*_*i*_) denotes the filter’s frequency response to the eigenmode Ψ_*i*_ associated with the frequency *λ*_*i*_. An ideal low-pass filter *H*_|*k*)_ with frequency response *h*_*k*_ that discards the information of eigenmodes with graph frequencies higher than *λ*_*k*_ is defined using the step function:

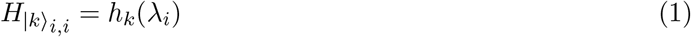

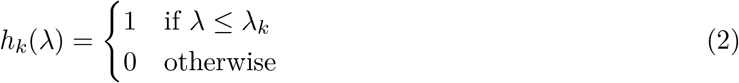

Using this ideal low-pass filter, brain signal *x* is approximated by 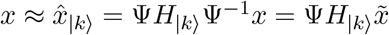, where 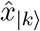 is the low-pass filtered approximation of *x* reconstructed from the first *k* eigenmodes. It should be noted that all elements beyond the *k*th row and column of *H*_|*k*)_ are zeros. As a result, this approximation does not require knowledge about higher frequency eigenmodes Ψ_*i*_ : *i > k*. Specifically, let 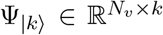 denote the submatrix of Ψ formed by its first *k* eigenmodes, Ψ_|*k*)_ = [Ψ_1_, …, Ψ_*k*_], i.e., the first *k* columns of Ψ. We use 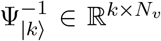 to denote the corresponding submatrix of Ψ^−1^ containing the first *k* rows, such that 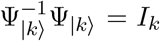. The low-pass approximation of *x* is then given by:

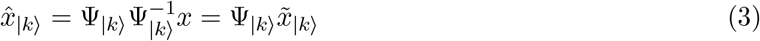

Where 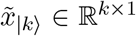 is the latent spectral representation of *x* via the first *k* modes. This low-pass filtering effectively reduces an *N*_*v*_-dimensional brain signal *x* to *k* dimensions. We anticipate that the majority of spatial variation in a brain signal such as cortical thickness is captured by a low-pass approximation of adequate bandwidth (i.e., an appropriate *k*). The choice of *k* controls the trade-off between computational complexity and spatial specificity; lower *k* reduces computation but spatially simplifies the signal; an optimal *k* balances between computational tractability and spatial specificity.

### 4.8 Normative Modeling Framework

In the context of our paper, conventional normative models encompass all univariate approaches in which the normative ranges of a fixed spatial query (e.g., the thickness of a region of interest) are modeled as a function of demographic covariates. SNM is a general framework that is compatible with alternative normative models, such as GAMLSS^2,197^, BLR^17,198^, and HBR^15,187^, etc.). For clarity and without loss of generality, we focus on an exemplary instantiation using Hierarchical Bayesian Regression (HBR)^15^, although the accompanying software package already supports Bayesian Linear Regression (BLR)^17,198^ as an alternative model. We begin by formally defining a spatial normative query to specify the region of interest, followed by an overview of HBR (referred to as the direct method, see Fig. 1A). We then describe how SNM extends the direct approach by using eigenmodes, facilitating the estimation of arbitrary, a posteriori-defined spatial normative queries.

#### 4.8.1 Spatial Normative Query

The concept of a spatial normative *query* is introduced in this work as a mechanism for specifying the spatial extent over which a normative range is to be evaluated. In practical terms, we may wish to determine the normative range of cortical thickness over a defined region of interest, which can be as fine-grained as a single vertex on the cortical mesh or as broad as a parcel, functional network, or even the entire cortex. Formally, a spatial normative query is represented as a weight vector 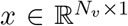 that linearly maps a cortical phenotype (e.g., a subject’s thickness map) to a single summary value via *y* = *t*^⊤^*x*, where 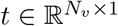 denotes the subject-specific thickness measurement.

In its simplest form, a spatial query corresponds to an average over a region of interest. For example, the query defining the average thickness over the entire cortex can be written as 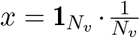, where 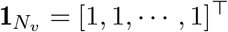 denotes a unit vector of length *N*_*v*_. Likewise, if the region of interest consists of *N*_*r*_ vertices, we assign uniform weights to those vertices and zero elsewhere, defining the query as

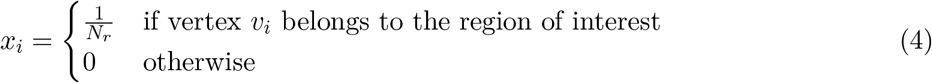

Here, *x*_*i*_ denotes the *i*th element of 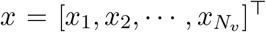. This formulation provides a flexible interface through which any region, parcel, or network can be queried by adjusting the weights in *x*.

While spatial normative queries are often expressed as simple averages, this formulation is not restricted to this case. In general, any weight vector *x* that encodes a meaningful spatial hypothesis can serve as a query, allowing users to interrogate more complex composite spatial patterns of interest. For example, the normative thickness of a functional network can be queried by assigning weights proportional to the probability that each vertex belongs to that network (with normalization such that 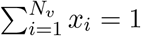).

Importantly, this flexibility also enables comparative queries, facilitating the assessment of relative normative variations between regions. For instance, cortical asymmetry (lateralization) can be evaluated by assigning positive weights to vertices in the left hemisphere and negative weights to those in the right hemisphere:

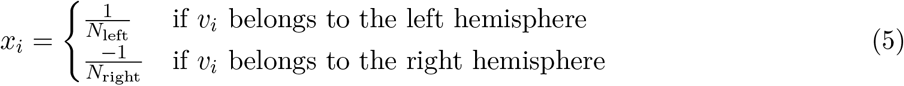

where *N*_left_ and *N*_right_ denote the number of vertices in each hemisphere.

Importantly, the accompanying software package streamlines the application of spatial queries by providing various built-in utilities, e.g., to project FreeSurfer outputs from native surfaces onto the standard template and to construct queries from arbitrary brain parcellations. This allows users to apply the pre-trained model directly, without having to reimplement preprocessing or query-construction procedures.

#### 4.8.2 Direct Method

The direct method refers to the conventional normative modeling approach in which a fixed a-priori-defined spatial query is used (e.g. mean thickness). In this manuscript, we utilize HBR as the normative approach used for the direct method^15,187^, allowing modeling of site variability as random effects via partial pooling and reuse of site hyperpriors to support normative inference at unseen sites. Using a fixed spatial query *x*, the direct method models the expression of the phenotype (e.g. cortical thickness) on the query in the training set, 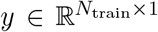; which is computed as the inner product between the spatial query *x* and the high-resolution thickness data *T, y* = *T*_train_. *x*. HBR assumes that these observations follow a normal distribution 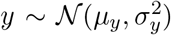 and aims to model its moments (mean and variance) as a function of covariates (age and sex) while controlling for batch effects:

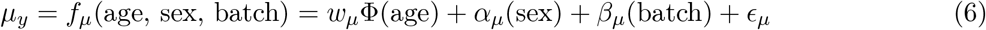

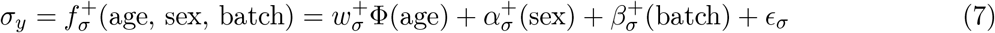

In particular, *w*_*µ*_ and 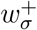 are weights that model the fixed effect of age by a basis expansion φ. A B-spline basis expansion (cubic spline with three evenly spaced knots; df=5) was used to capture the non-linear effects of age. The fixed effect of sex is modeled by intercepts for mean (*α*_*µ*_) and deviation 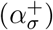. Batch effects (for dataset harmonization) are modeled as intercepts for the mean 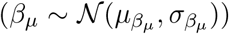 and deviation 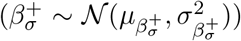 that are randomly drawn from another prior distribution; this results in hierarchically modeling batch as a random effect. The prior assumptions of the Bayesian model can be summarized as follows:

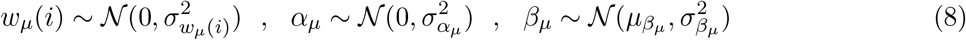

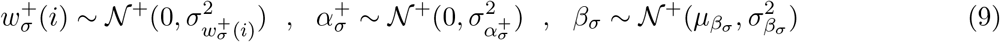

Where *N*^+^ denotes a positive half-normal distribution. The random effect parameters are hierarchically sampled from a set of batch hyperparameters:

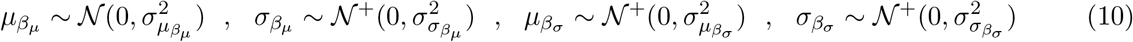

Notably, the model can quantify the normative deviation of an individual subject *y*_*i*_ using a z-score:

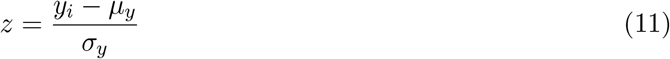

This z-score can be used to compute normative centiles using the cumulative distribution function (CDF) of the normal distribution.

#### 4.8.3 Spectral Normative Model (SNM)

Next, we explain the theory behind the proposed SNM framework. In the training phase, *k* direct normative models are independently fitted to the projection of phenotypes on the first *k* brain eigenmodes with the lowest graph frequencies. This forms the low-dimensional latent approximation containing information about the low-frequency characteristics of the cortical phenotype. For example, in the case of cortical thickness, for each *i* ≤ *k*, the spectral coefficient 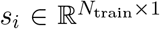 associated with Ψ_*i*_ is first computed:

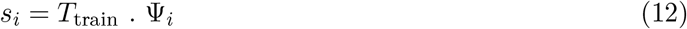

Similar to the direct model, normative estimators of the mean 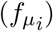 and deviation 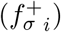 are trained to model normative ranges of all low-frequency spectral coefficients:

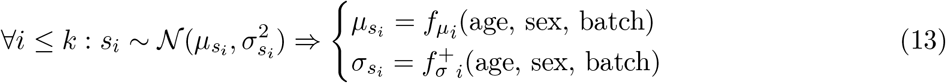

The normative model for each spectral coefficient is the same as the direct case (see equations 6, 7, 8, 9, 10). Next, the normative ranges of any arbitrary spatial query is approximated using this set of *k* pre-trained normative models. For any spatial query *x*, its observed cortical phenotype *y* = *T*_train_. *x* is approximated by a low-pass graph filter (see equation 3):

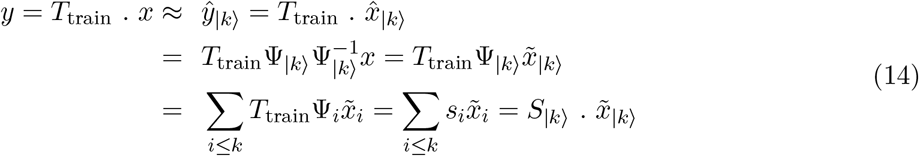

where 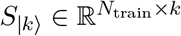 denotes the matrix containing the first *k* spectral coefficients. The variable of interest *y* can hence be approximated by a linear combination of the spectral coefficients *S*_|*k*)_. Notably, if the spatial query is frequency-bounded this approximation will be exact (i.e. if the query is a linear combination of low-frequency eigenmodes such that 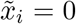 for *i > k*). Equation 14 indicates that for any spatial query *x*, the associated phenotype *y* can be modeled as a multivariate normal distribution such that its normative ranges are estimated by the linear combination 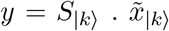. This is formalized by the following equation:

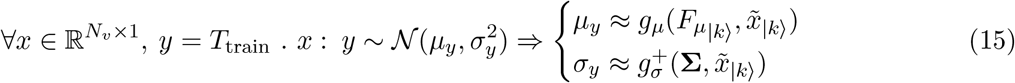

Where *g*_*µ*_ and 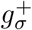 are functions that respectively approximate the mean and standard deviation of the multivariate normal distribution based on the trained low-pass spectral moments 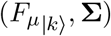. Explicitly, *g*_*µ*_ defines the mean of the normal distribution *y* as a linear combination of the means of each spectral coefficient and is based on the graph spectral encoding 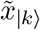:

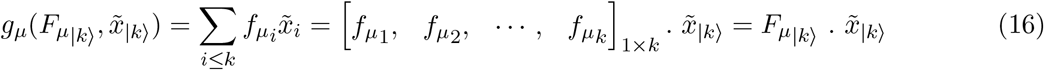

where 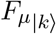 is used to summarize the set of estimated means for the first *k* spectral coefficients in *S*_|*k*)_ into a 1×*k* vector. Note that equation 16 arises because the expected value of a linear combination of a set of random variables is equal to the linear combination of their respective expected values:

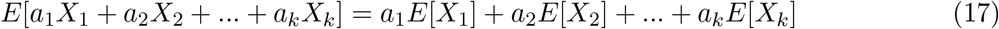

Moreover, 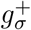 defines the standard deviation of the normal distribution *y* as a function of the covariance matrix describing cross-basis dependencies of spectral coefficients 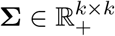:

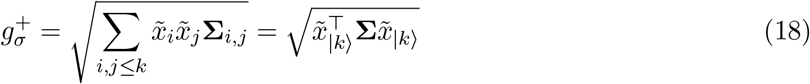

Equation 18 describes how the standard deviation of the variable of interest *y* is related to the covariance matrix Σ describing dependencies across spectral coefficients *s*_*i*_. This equation is based on the following notion that relates the variance of a linear combination of a set of normally distributed variables to their covariance structure:

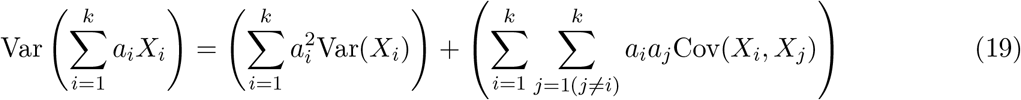

Elements of Σ can also change as a function of covariates and batch effects. We note that for a pair of random variables (*X* and *Y*), covariance (Cov(*X, Y*)) and correlation (Corr(*X, Y*)) are related by Cov(*X, Y*) = *σ*_*X*_*σ*_*Y*_ Corr(*X, Y*), where *σ* denotes the standard deviation of each variable. We use this notion to separate the effect of within-mode variance from the effect of cross-mode correlation on Σ. In particular, the following equation decomposes the cross-basis covariance matrix Σ into a correlation matrix P and an independent set of eigenmode standard deviations:

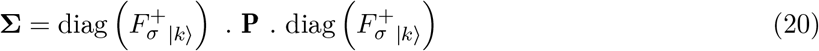

where 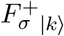 summarizes the set of estimated standard deviations of the first *k* spectral coefficients (*y*_1_, · · ·, *y*_*k*_) to a 1 × *k* vector, and the *diag()* operator is used to map a 1 × *k* vector to a *k* × *k* diagonal matrix:

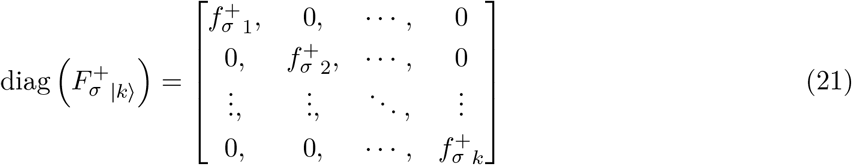

Thus far, we show that any spatial query can be represented as a multivariate normal distribution across *k* spectral components, the normative ranges of which can be approximated from the mean 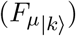, standard deviation 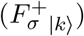, and cross-correlation (P) of spectral coefficients. Mean and standard deviation estimates are computed similarly to a direct HBR model. However, approximations may be needed to estimate the cross-correlation structure. This is because the complexity of the correlation structure increases quadratically with the number of eigenmodes included in the low-pass approximation (*k*). We hence impose a sparsity constraint on the correlation structure using a threshold on the observed cross-correlation of spectral phenotypes within the training sample. Our evaluations indicate that the cross-correlation matrix is naturally sparse (see Supplementary Fig. S.3). As a result, the majority of elements P_*i,j*_ : Corr(*s*_*i*_, *s*_*j*_) ≤ *ρ* are small and will hence be replaced with zero through this sparsification step. The remaining pairs of suprathreshold correlations form a sparse matrix representation as summarized by the following derivation of P:

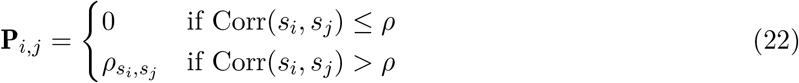

The suprathreshold elements 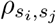 are affected by covariates (age and sex) and batch effects. As such, for every suprathreshold element, the spectral coefficient pair was modeled as a bivariate normal distribution:

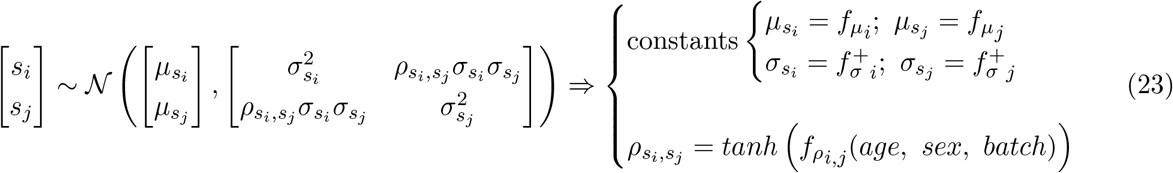

where spectral coefficients’ estimates of the mean 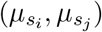 and deviation 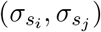 were inputted as constants (yielded from the solution of Equation 13). The cross-correlation estimate 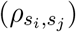 was adjusted by a hyperbolic tangent function to limit the range of possible correlation values to [−1, 1]. Effects of covariates and batch were modeled as follows:

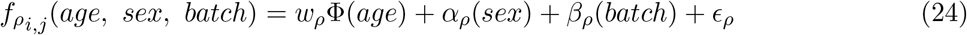

with the following prior assumptions:

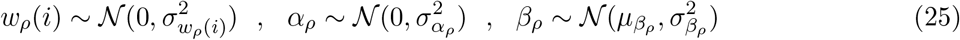

and the following hierarchical batch priors:

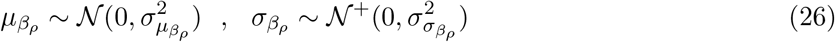

In summary, Equations 12–26 formalize how SNM derives normative ranges for a target phenotype *y*. The method first projects the cortical data for a given spatial query 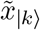 into the low-dimensional spectral domain, then models the resulting spectral coefficients *S*_|*k*)_ as a multivariate normal distribution. Normative means and deviations for the target phenotype are then estimated directly from this distribution, enabling rapid inference for arbitrary spatial queries without requiring direct training on novel spatial queries.

### 4.9 Model Evaluation

We use three different families of spatial normative queries (*x*) to evaluate SNM’s performance. These families include (I) *brain-wide*, (II) *regional*, and (III) *high-resolution* signals, each of which groups queries according to their spatial scale. Brain-wide queries are regional masks that describe coarse and large-scale characteristics along the cortex. Namely, this includes a total of 25 queries that represent the average of total cortical thickness, as well as the average thickness over predefined brain networks that divide the brain into 7/17 functionally distinct segments (Yeo functional networks)^87^. Regional queries included 200 signals, each describing the average cortical thickness within a region of interest as defined by a brain parcellation. For regional queries, we used a homotopic parcellation of the cortex comprising 200 cortical regions^199^ (Yan200 atlas). In the supplementary analysis, we evaluate the sensitivity of our findings to atlas granularity (see Supplementary Figs. S.10, S.11). Finally, a total of 4,000 high-resolution queries were constructed (2,000 per hemisphere), each describing the average cortical thickness in the vicinity of a selected cortical vertex. Vertices were randomly sampled to ensure uniform coverage across all cortical regions (see Supplementary Section S.1.6 Benchmarking High-Resolution Spatial Queries). This subsample is used solely for evaluation purposes; SNM itself scales efficiently to full vertex resolution (i.e., hundreds of thousands of vertices). The limiting factor in our benchmark design is the direct model comparison, which requires fitting a new model for each query, making large-scale comparison computationally prohibitive.

Every high-resolution query was formed by an 8mm FWHM smoothing kernel based on the geodesic distance to the respective vertex^191^. The smoothing kernel serves two key purposes: (1) to mitigate the impact of local intersubject registration misalignments and (2) to ensure vertex-resolution queries are appropriately reconstructed by the low-pass graph filter, thereby reducing ringing effects in reconstructions. In a supplementary analysis, we evaluate the sensitivity of our evaluations to the smoothing kernel choice (see Supplementary Figs. S.10, S.12). The signals evaluated here quantify the average thickness of the cortex with different spatial granularities (ranging from tens of centimeters to a few millimeters). While such average queries are the only type of normative queries that have been previously studied for brain charting, in supplementary analyses we evaluate the applicability of our proposed model to other possible cases, e.g. when a comparative asymmetric norm is being studied (see Supplementary Figs. S.13, S.14, S.15, and S.16).

For all signal families (whether presented in the main text or supplementary information), we evaluated the performance of SNM (using different values of *k*) based on (i) reconstruction accuracy, normative model’s (ii) goodness of fit of the central tendency, and (iii) total goodness of fit of the estimated distribution. These evaluation criteria are detailed in the ensuing sections.

#### 4.9.1 Reconstruction Accuracy

For the spectral approach to provide accurate estimates of normative ranges, both the cortical phenotype under study (i.e. cortical thickness) and the spatial normative query must be adequately reconstructed by the low-dimensional latent space defined by the brain eigenmodes. We hence use the following two metrics to evaluate the validity of the brain eigenmodes for reconstructing brain signals:

##### Energy Proportion

For any arbitrary brain signal 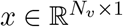 with graph spectral encoding 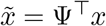, the total energy of the signal can be quantified by 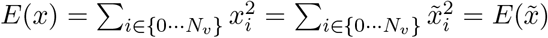. In other words, the signal energy in vertex domain is equal to the signal energy in the spectral domain (Parseval’s theorem^200^). As such, the proportional contribution of every single eigenmode Ψ_*i*_ to the total signal energy can be quantified by 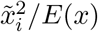. We provide a cumulative line plot of observed ranges for this proportional energy for different brain signals. This indicates the number of eigenmodes that capture major energy proportions for different brain signals.

##### Standardized Mean Squared Error (SMSE)

This measure normalizes the mean squared error (MSE) by signal variance to provide an error metric that does not depend on signal variance^201^. Using this metric, a trivial reconstruction that replaces values of all vertices *x*_*i*_ with the signal mean will have an SMSE of one, and a perfect reconstruction will result in an SMSE of zero. By considering 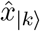 to be a reconstruction of the original brain signal *x*, we compute SMSE for different *k*s to quantify reconstruction error at different low-pass frequency bandwidths. A high SMSE (close to one) indicates poor reconstruction performance and a low SMSE (close to zero) marks accurate reconstruction.

#### 4.9.2 Model’s Central Tendency

Independent of how accurately the eigenmode basis reconstructs various brain signals, we also need to validate that SNM yields accurate normative fits for spatial queries. This is quantified by measures of goodness of fit of the central tendency^2^. These measures can particularly indicate the accuracy of the normative model in explaining trends in the data. To this end, we utilize the mean absolute error (MAE) and explained variance (EV) metrics. An out-of-sample MAE score was evaluated using 20% of the data in the test set. Specifically, for any spatial query *x*, the thickness of that query in the test set is measured 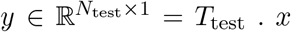 and MAE quantifies the extent to which these thickness values deviate from model predictions 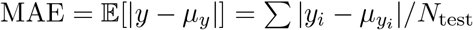. In addition to MAE, we also quantify the out-of-sample explained variance (EV), which measures the proportion of phenotype variance in the test sample that is captured by the model predictions, EV = 1 −Var(*y* − *µ*_*y*_)/Var(*y*). MAE and EV were computed for each of the three families of spatial queries, comparing the performance of a direct HBR model against four alternative SNMs, which utilized the first 10, 100, 1,000, and 10,000 eigenmodes, respectively. Since all models provided mean thickness estimates in millimeters, MAE offers a practical measure of the average error in millimeters when inferring the thickness of a given spatial query.

#### 4.9.3 Model’s Total Goodness of Fit

Measures of central tendency, such as mean or median, assess only the accuracy of modeling normative trends but do not capture the validity of the estimated distribution’s deviations. To evaluate the model’s overall goodness of fit—including both mean and variance estimates—we utilize the mean standardized log loss (MSLL)^201^. Mean log loss quantifies the mean negative log probability of the observed data (in the test sample) under the hypothesis of the fitted normative model:

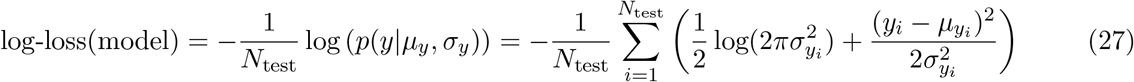

MSLL is computed by comparing the log loss of competing models against the log loss of a trivial reference model that is based on the mean and standard deviation of the training sample, MSLL = log-loss(model) − log-loss(reference). By definition, the trivial model achieves an MSLL of zero, and models providing better than trivial normative ranges will yield negative MSLL values. Here, we estimate log loss using a censored log-likelihood measure to reduce the sensitivity to outliers^2,202^. As such, for observations falling outside of the range of 1^st^ to 99^th^ percentile, the value of log loss was replaced by − log(0.02). This censored MSLL score favors models with accurate distribution estimates within this percentile range and is thus robust against extreme outliers.

### 4.10 Mapping Cortical Growth Gradients

The cortical growth gradients reported in Section 2.4 Gradients of Lifespan Thickness Growth were derived using a principal component analysis (PCA) framework applied to normative lifespan predictions from SNM. Specifically, the pretrained SNM with *k* = 1000 eigenmodes was used to estimate median cortical thickness at all *N*_*v*_ = 59,412 vertices across 1-year age intervals spanning 5 to 95 years, while partialling out site and sex effects. This procedure yielded a matrix 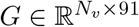 containing vertex-wise normative thickness trajectories across the lifespan. To ensure comparability across vertices while preserving relative biological variability, *G* was globally standardized by subtracting its mean and dividing by its standard deviation, 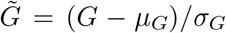. This normalization retains relative magnitude differences across cortical regions, allowing vertices with greater lifespan variability to contribute proportionally to the extracted components.

Centered PCA was then applied to 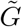 to identify orthogonal axes of maximal variance. The first three principal components, termed thickness growth gradients (TGGs), were retained and mapped back onto the cortical surface to represent dominant spatial modes of lifespan cortical growth. The first component (TGG1) captured a global thickness factor closely aligned with mean cortical morphology and accounted for 97.4% of the total variance. To examine more subtle spatial patterns beyond this global effect, we analyzed the second and third components (TGG2 and TGG3), which together explained 91.5% of the residual variance not captured by TGG1 (66.5% by TGG2 and 25.0% by TGG3). Conceptually, this procedure summarizes shared and divergent patterns of cortical thickness change across the lifespan into a small number of spatially interpretable gradients.

#### 4.10.1 Charting Along Gradients

SNM’s spatial query framework also supports the characterization of normative cortical trajectories along continuous spatial gradients. For each growth gradient (TGG1–3), we stratified the cortex at regular intervals along the gradient using a probabilistic approach. First, the vertex-wise values of each gradient map (Fig. 4A) were transformed into centile maps ranging from 0 to 100 across all vertices (Fig. 4B). We defined 11 target centiles, *C* = [0, 10, 20, …, 100], to serve as reference points along each gradient. For each centile *c*_*j*_, we applied a Gaussian weighting function to the gradient map *i* to define smooth gradient strata:

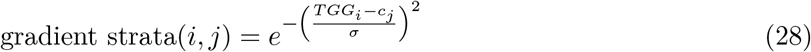

where *σ* denotes the kernel width and was set to 10 centiles FWHM. This probabilistic weighting allows vertices near the target centile to contribute strongly to its normative trajectory. Each stratum was normalized to sum to one, producing a spatial query that represents the weighted average cortical thickness of regions near the target centile along the gradient (see Extended Data Figs. E.2, E.3, E.4 for illustrative examples). Finally, the pretrained SNM was used to estimate normative cortical thickness trajectories of these spatial queries (Fig. 4C,D, and Extended Data Figs. E.5, E.6, E.7).

### 4.11 Testing Spatial Correspondence of Cortical Maps

Spatial correspondence between cortical maps was quantified using Pearson’s correlation coefficient. Statistical significance was assessed using two-sided non-parametric permutation tests (10,000 permutations) based on surrogate maps that preserve the spatial structure of the empirical data, controlling false discovery rate (FDR) using the Benjamini–Hochberg procedure within each family of tests. Two permutation frameworks were used according to the highest spatial resolution at which all maps in a given comparison were available. When all maps in a comparison were available at vertex resolution (*N*_*v*_ = 59,412), spatial correspondence was assessed using spin permutations^203^. This approach was used for comparisons between the thickness growth gradients and cortical thickness or thickness-change maps (Fig. 4E–G; Extended Data Figs. E.8, E.9). For comparisons involving maps available only at parcel resolution, all maps were instead transformed to a common 360-parcel parcellation by averaging vertex-wise values within each parcel^38^. As spin permutations are less suited to low-resolution parcellated maps, spatial-autocorrelation-preserving surrogate maps were generated using BrainSMASH^42^. This framework was used for comparisons with established cortical hierarchies and published neuromaps (Fig. 5; Supplementary Fig. S.17).

Among these maps, age-resolved cortical thickness and thickness-change maps were derived from the pretrained spectral normative model (SNM-1000). Vertex-wise predicted median cortical thickness and its annual rate of change, computed as the first derivative of the predicted median with respect to age, were sampled at 5-year increments from 5 to 95 years and compared with each gradient to characterize age-specific and lifelong correlates (Fig. 4F,G; FDR-corrected across the 3 × 19 = 57 tests within each panel). Normalizing the annual change by the predicted median thickness at each age and averaging across specified age windows yielded two additional SNM-derived maps: developmental cortical pruning (DCP; mean annual percent change, ages 5–20) and aging-related thickness decline (ATD; ages 40–95), which were used in subsequent correspondence analyses (Fig. 4E; Fig. 5; Extended Data Fig. E.9).

Alignment with laminar cytoarchitecture (Fig. 5A) was assessed by assigning each parcel to one of four laminar cytoarchitectonic classes, defined at parcel resolution^41,43^, and centile-ranking gradient values across parcels. Differences between class mean scores were tested for each gradient using an omnibus ANOVA, with significance assessed against the BrainSMASH surrogate null, FDR corrected across the three tests. The contribution of individual classes was then quantified as the z-score of each class mean’s signed deviation from the gradient median, normalized against the same surrogate distribution, with significance FDR-corrected across the twelve tests.

Seeking to situate the derived thickness growth gradients within previously established cortical hierarchies, we performed an exploratory spatial correspondence analysis across a set of 17 cortical maps (Fig. 5B), comprising:

- The three thickness growth gradients (TGG1–3);
- Mean cortical thickness^38^;
- Aging-related thickness decline (ATD) and developmental cortical pruning (DCP);
- Three cortical gene-expression components (GEC1–3)^43^;
- The first two functional connectivity gradients (FCG1–2)^46^;
- The sensorimotor–association axis (SA)^45^;
- The functional development axis (FCDA)^48^;
- The lifespan functional growth axis (LFGA)^49^; and
- Three perinatal thalamocortical connectivity components (ThalC1–3)^47^.

Published maps were obtained from their original sources or via neuromaps^50^. All 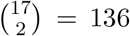 pairwise correspondences were tested, with FDR correction across the full set. The resulting correlation matrix was clustered based on correlation magnitudes to summarize its correspondence structure, yielding three clusters. For visualization, maps were sign-flipped where necessary such that within-cluster correspondences were positive (Fig. 5B). In a complementary extended analysis, TGG1–3 were compared with a broader library of 81 published cortical maps, yielding 81 × 3 = 243 tests with FDR correction; this analysis is detailed in Supplementary Section S.1.9 Extended Spatial Correspondence Analysis of Cortical Maps.

### 4.12 Clinical Evaluations

We examined the clinical insights provided by SNM when applied to the clinical MACC dataset. A subset of 66 healthy individuals (50% of the HC data) from the MACC cohort was used to fine-tune the pretrained SNM for the clinical sample. During fine-tuning, parameters linking cortical phenotypes to age and sex 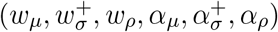 were held constant, while only site-specific parameters for batch effect harmonization 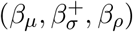 were updated based on the MACC data. The fine-tuned model was then applied to generate normative deviation maps for smoothed, vertex-resolution cortical thickness phenotypes.

#### 4.12.1 HC vs. AD Comparison

A two-sample t-test was conducted to compare high-resolution deviation maps between the healthy and AD groups within the MACC dataset. To identify vertices with statistically significant differences, a nonparametric false discovery rate (FDR) correction^204^ was applied across all vertices (FDR-*α* = 5%, 1000 permutations), controlling for multiple comparisons. Vertices surpassing the corrected threshold were deemed to exhibit significantly different deviation patterns between groups.

#### 4.12.2 Linking Atrophy to Cognitive Impairment

We next evaluated the potential of normative deviation maps to serve as biomarkers of cognitive impairment. To begin, we performed a univariate vertex-wise assessment, measuring the Pearson correlation coefficient between normative z-scores and cognitive performance scores (MMSE). The significance of associations was evaluated with a nonparametric FDR correction via permutation testing. Specifically, we shuffled MMSE scores across 1000 permutations, calculating correlation coefficients for each permutation to produce a null distribution under the hypothesis of no association between normative deviation and cognitive performance. Using this null distribution, we derived nonparametric p-values for each vertex, which were subsequently FDR-corrected at *α* = 5%.

In addition, we derived an extreme value statistic^60^ for atrophy by counting vertices with a z-score below a threshold of −1.96 for each individual. This measure, termed Extremely Thin Vertex Count (ETVC), was then tested for its predictive power on MMSE scores via Pearson correlation. To assess the sensitivity of ETVC to the threshold choice, we repeated this analysis for a range of z-score thresholds. We performed these assessments within each cohort (HC, MCI, and AD) to evaluate the specificity of ETVC as a biomarker for AD-related cognitive impairment, noting that no meaningful association is expected in HC, weaker associations may be present in MCI, and stronger associations are anticipated in AD (see Supplementary Section S.1.10 Within-Group Cognitive Associations).

#### 4.12.3 Examining Interindividual Heterogeneity

Next, we aimed to demonstrate the capabilities of the SNM in revealing the heterogeneity landscape of brain atrophy in AD. We quantified interindividual differences in atrophy by computing the Euclidean distance between vectors of vertex-level z-score deviation maps, yielding a *N*_*MACC*_ × *N*_*MACC*_ distance matrix that captures interindividual differences. This matrix was visualized as a heatmap to provide an overview of the variation across individuals. Additionally, we applied multidimensional scaling (MDS)^205^ to reduce the distance matrix to a 2-dimensional embedding, providing a spatial representation of interindividual differences. Average deviation maps for subgroups within local vicinities in this 2-dimensional space were then visualized as cortical projections (see Fig. 8 and Supplementary Section S.1.13 Heterogeneity Landscape Subgroups).

To quantitatively verify the illustrated heterogeneity, we compared distance distributions across groups using two-sample t-tests and computed Cohen’s *d* effect sizes. These comparisons test the hypothesis that individuals with MCI and AD exhibit progressively greater deviations from healthy norms, as well as increased interindividual variability within each diagnostic group, consistent with the observed heterogeneity landscape. We next assessed whether deviation patterns exhibit discrete cluster structure or instead reflect continuous variation, using five complementary criteria: multimodality of the pairwise-distance distribution (Hartigan’s dip statistic^62,66^), cluster tendency relative to a reference distribution without cluster structure (gap statistic^63^), stability of individual clusters under resampling (bootstrap cluster stability^64^), generalizability of inferred clusters across independent subsamples (prediction strength^65^), and statistical significance of hierarchical cluster splits against a multivariate Gaussian null (SigClust2^67^). Evaluations were tested across dimensional representations to verify that findings were not artifacts of dimensionality reduction. Full methodological specifications and results are provided in Supplementary Section S.1.15 Clusterability of AD Deviation Patterns. Together, these analyses complement the visualizations by quantitatively characterizing both the magnitude and structural organization of heterogeneity within and between diagnostic groups.

## Data Availability

This study integrates multiple large-scale neuroimaging datasets (see Supplementary Information, Section S.1.1, Healthy Lifespan Imaging Data), each subject to its own data access and usage agreements. While some datasets are publicly available, others require approval from data access committees or the execution of data use or material transfer agreements. Researchers interested in accessing these datasets should consult the respective data providers for specific access procedures.

https://www.ukbiobank.ac.uk/

https://www.humanconnectome.org/study/hcp-lifespan-development

https://www.humanconnectome.org/study/hcp-young-adult

https://www.humanconnectome.org/study/hcp-lifespan-aging

https://medicine.nus.edu.sg/macc-2/projects/harmonization-study

http://www.macc.sg/

https://abcdstudy.org/

https://fcon_1000.projects.nitrc.org/indi/abide/

https://fcon_1000.projects.nitrc.org/indi/adhd200/

https://adni.loni.usc.edu/data-samples/adni-data/

## Code and Data Availability

All scripts used to conduct the analyses and generate the figures in this study are openly available at https://github.com/sina-mansour/normative_brain_charts. The repository additionally hosts supplementary resources, including the pretrained SNM-1000 model, derived annual normative charts for several established brain atlases, as well as cortical maps of thickness growth gradients.

An online version of the full analysis pipeline is available at https://sina-mansour.github.io/normative_brain_charts/, allowing every notebook and its corresponding outputs to be browsed without local execution or access to dedicated computing resources. The site also provides a reproducibility walkthrough (https://sina-mansour.github.io/normative_brain_charts/reproducibility_walkthrough/) detailing environment setup, approximate runtimes and hardware requirements for each stage of the pipeline, together with a map linking each manuscript figure to the notebook cells that produce it.

To facilitate broader adoption and reuse, the methodological components of the SNM framework are provided in the `spectranorm` Python software package^206^ (https://sina-mansour.github.io/spectranorm/). The accompanying website provides comprehensive documentation and tutorials describing the range of normative modeling functionalities supported by the software.

This study integrates multiple large-scale neuroimaging datasets (see Section S.1.1 Healthy Lifespan Imaging Data), each subject to its own data access and usage agreements. While some datasets are publicly available, others require approval through data access committees or the execution of material transfer or data-use agreements. Researchers interested in accessing these datasets should consult the corresponding data providers for specific access procedures.

## Acknowledgments

The analysis for this study was supported by the Spartan High-Performance Computing infrastructure^207,208^ and dedicated computing and storage solutions provided by Research Computing Services at the University of Melbourne. We gratefully acknowledge the invaluable contribution of several open-source software packages that significantly facilitated our data analysis and interpretation. The analytical pipeline made use of several open-source Python packages, including Numpy^209^, Scipy^210^, Matplotlib^211^, Pandas^212^, scikit-learn^213^, PyMC^214^, xarray^215^, and Nibabel^216^. The connectome spatial smoothing package^190^ was utilized to compute smoothing kernels. Brain visualizations were generated by the Cerebro brain viewer^217^. This study was supported by the NHMRC Ideas Grant (GNT2011592) titled *Normative Reference Ranges for Brain Phenotypes*, awarded to A. Zalesky, C. Bousman, C. Pantelis, and M. Di Biase; the NUS Yong Loo Lin School of Medicine (NUHSRO/2020/124/TMR/LOA); the Singapore National Medical Research Council (NMRC) OF-YIRG (OFYIRG25jan-0049), LCG (OFLCG19May-0035), NMRC CTG-IIT (CTGIIT23jan-0001), NMRC OF-IRG (OFIRG24jan-0030), NMRC STaR (STaR20nov-0003); Singapore Ministry of Health (MOH) Centre Grant (CG21APR1009); the Temasek Foundation (TF2223-IMH-01); and the United States National Institutes of Health (R01MH133334 to B.T.T.Y. and R01MH133843 to A.A.B.). Any opinions, findings, conclusions, or recommendations expressed in this material are those of the authors and do not reflect the views of the funders.

We gratefully acknowledge the use of several openly shared MRI biobank initiatives that made this work possible, including OpenNeuro (https://openneuro.org/), the Reproducible Brain Charts (https://reprobrainchart.github.io/), the Healthy Brain Network (https://healthybrainnetwork.org/), UK Biobank (https://www.ukbiobank.ac.uk/), the Adolescent Brain Cognitive Development (ABCD) Study (https://abcdstudy.org/), the Human Connectome Project (http://www.humanconnectomeproject.org/), data made available by the Open Science Framework (https://osf.io/), the Laboratory of NeuroImaging (https://loni.usc.edu/), the International Neuroimaging Datasharing Initiative (INDI) (https://fcon_1000.projects.nitrc.org/), and the NIMH Data Archive (https://nda.nih.gov/).

HCP Young Adult data used in this study were provided by the Human Connectome Project, WU–Minn Consortium (Principal Investigators: David Van Essen and Kamil Ugurbil; 1U54MH091657), funded by the 16 NIH Institutes and Centers that support the NIH Blueprint for Neuroscience Research, and by the McDonnell Center for Systems Neuroscience at Washington University. Data from the HCP Aging cohort used in this study were supported by the National Institute on Aging of the National Institutes of Health under Award Number U01AG052564 and by funds provided by the McDonnell Center for Systems Neuroscience at Washington University in St. Louis. The HCP-Aging 2.0 Release data used in this report were obtained from DOI: 10.15154/1520707. Data from the HCP Development cohort used in this study were supported by the National Institute of Mental Health of the National Institutes of Health under Award Number U01MH109589 and by funds provided by the McDonnell Center for Systems Neuroscience at Washington University in St. Louis. The HCP-Development 2.0 Release data used in this report were obtained from DOI: 10.15154/1520708.

Data from the Adolescent Brain Cognitive Development^®^ (ABCD) Study used in this work were obtained from the NIH Brain Development Cohorts (NBDC) Data Sharing Platform. The ABCD Study is a multisite, longitudinal study designed to recruit more than 10,000 children aged 9-10 years and follow them over 10 years into early adulthood. The ABCD Study is supported by the National Institutes of Health and additional federal partners under award numbers U01DA041048, U01DA050989, U01DA051016, U01DA041022, U01DA051018, U01DA051037, U01DA050987, U01DA041174, U01DA041106, U01DA041117, U01DA041028, U01DA041134, U01DA050988, U01DA051039, U01DA041156, U01DA041025, U01DA041120, U01DA051038, U01DA041148, U01DA041093, U01DA041089, U24DA041123, and U24DA041147. ABCD Consortium investigators designed and implemented the ABCD study and/or provided data but did not necessarily participate in the analysis or writing of this report. This manuscript reflects the views of the authors and may not reflect the opinions or views of the NIH or ABCD Consortium investigators. The ABCD data repository grows and changes over time. The ABCD data used in this report came from the ABCD Release 6.0 (NIMH Data Archive DOI: https://doi.org/10.82525/jy7n-g441).

UK Biobank data used in this study were accessed under Application Number `60698`. Data from the Brazilian High-Risk Cohort (BHRC), the Philadelphia Neurodevelopmental Cohort (PNC), the developing Chinese Connectome Project (dCCNP), the Healthy Brain Network (HBN), and the Nathan Kline Institute (NKI) were accessed through the Reproducible Brain Charts (RBC) platform (https://reprobrainchart.github.io/). Data were provided in part by the Brain Genomics Superstruct Project (BGSP) of Harvard University and the Massachusetts General Hospital, (Principal Investigators: Randy Buckner, Joshua Roffman, and Jordan Smoller), with support from the Center for Brain Science Neuroinformatics Research Group, the Athinoula A. Martinos Centerfor Biomedical Imaging, and the Center for Human Genetic Research. 20 individual investigators at Harvard and MGH generously contributed data to the overall project.

Data used in preparation of this article were in part obtained from the Alzheimer’s Disease Neuroimaging Initiative (ADNI) database (https://adni.loni.usc.edu/). The investigators within the ADNI contributed to the design and implementation of ADNI and/or provided data but did not participate in analysis or writing of this report. A full list of ADNI investigators is available at https://adni.loni.usc.edu/wp-content/uploads/how_to_apply/ADNI_Acknowledgement_List.pdf. Data collection and sharing for ADNI is funded by the National Institute on Aging (National Institutes of Health Grant U19AG024904). The grantee organization is the Northern California Institute for Research and Education. In the past, ADNI has also received funding from the National Institute of Biomedical Imaging and Bioengineering, the Canadian Institutes of Health Research, and private sector contributions through the Foundation for the National Institutes of Health (FNIH) including generous contributions from the following: AbbVie, Alzheimer’s Association; Alzheimer’s Drug Discovery Foundation; Araclon Biotech; BioClinica, Inc.; Biogen; Bristol-Myers Squibb Company; CereSpir, Inc.; Cogstate; Eisai Inc.; Elan Pharmaceuticals, Inc.; Eli Lilly and Company; EuroImmun; F. Hoffmann-La Roche Ltd and its affiliated company Genentech, Inc.; Fujirebio; GE Healthcare; IXICO Ltd.; Janssen Alzheimer Immunotherapy Research & Development, LLC.; Johnson & Johnson Pharmaceutical Research & Development LLC.; Lumosity; Lundbeck; Merck & Co., Inc.; Meso Scale Diagnostics, LLC.; NeuroRx Research; Neurotrack Technologies; Novartis Pharmaceuticals Corporation; Pfizer Inc.; Piramal Imaging; Servier; Takeda Pharmaceutical Company; and Transition Therapeutics.

Data for this study were provided in part by the Australian Schizophrenia Research Bank (ASRB), accessed with approval from the ASRB Access Committee. The ASRB was supported by funding from the National Health and Medical Research Council (Enabling Grant 386500), the Pratt Foundation, Ramsay Health Care, the Viertel Charitable Foundation, and the Schizophrenia Research Institute. Data were provided in part by OASIS-3: Longitudinal Multimodal Neuroimaging Dataset: Principal Investigators: T. Benzinger, D. Marcus, J. Morris; NIH P30 AG066444, P50 AG00561, P30 NS09857781, P01 AG026276, P01 AG003991, R01 AG043434, UL1 TR000448, R01 EB009352, P30 AG066444, AW00006993.

Data used in part in this study were obtained from the Australian Imaging Biomarkers and Lifestyle flagship study of ageing (AIBL), a consortium comprising Austin Health, the Commonwealth Scientific and Industrial Research Organisation (CSIRO), Edith Cowan University, the Florey Institute for Neuroscience and Mental Health (University of Melbourne), and the National Ageing Research Institute. Numerous commercial interactions have supported data collection and analyses. In-kind support has been provided by the consortium institutions, as well as Alzheimer’s Research Australia, Sir Charles Gairdner Hospital, Hollywood Private Hospital, and St Vincent’s Hospital. The study has also received financial support at various stages from the Alzheimer’s Drug Discovery Foundation, the Victorian Government’s Operational Infrastructure Support program, Alzheimer’s Research Australia, and the National Health and Medical Research Council (NHMRC). The AIBL researchers contributed data but did not participate in analysis or writing of this report. AIBL researchers are listed at https://aibl.org.au/. Data for this study were provided in part by the Growing Up in Singapore Towards Healthy Outcomes (GUSTO) study. The GUSTO dataset was supported by funding from the NRF’s Human Health and Potential Domain, under the Human Potential Programme. Data used in preparation of this article were in part obtained from the SG70 study. The SG70 study was supported by Singapore National Medical Research Council [CSA-SI (MOH-000434)] and Yong Loo Lin School of Medicine, National University of Singapore.

## Extended Data Figures

**Extended Data Fig. E.1.**
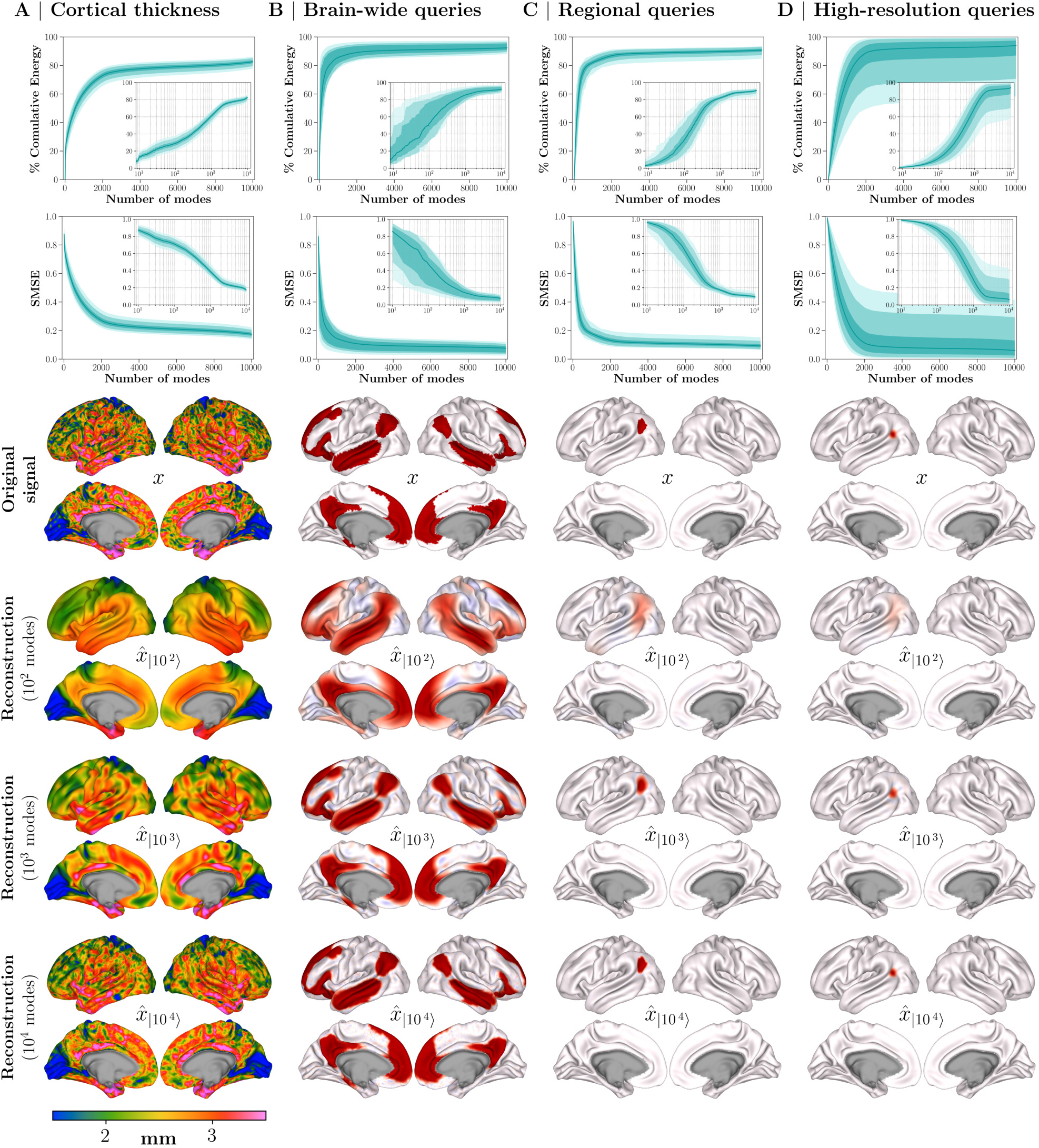
Eigenmode-Based Reconstruction of Cortical Thickness and Spatial Query Maps. Eigen-mode reconstruction accuracy for **(A)** individual cortical thickness maps and three families of spatial queries, including **(B)** brain-wide, **(C)** regional, and **(D)** high-resolution queries. Shaded line plots show the median reconstruction performance across observations, with shaded regions indicating the [25, 75], [5, 95], and [1, 99] percentiles. Cortical thickness results are based on 5,000 randomly selected individual healthy scans, while spatial query results include 25 brain-wide, 200 regional, and 4,000 high-resolution queries (see Section 4.9 Model Evaluation). The first and second rows report cumulative signal energy and standardized mean squared error (SMSE), respectively, as a function of the number of low-frequency eigenmodes used for reconstruction (logarithmic x-axis shown in the insets). The third row displays an example map from each category, and subsequent rows show reconstructions using 100, 1,000, and 10,000 eigenmodes. Cortical thickness projections in panel A correspond to a single representative individual. Additional supplementary analyses examine regional variability in reconstruction accuracy, as well as the effects of spatial granularity and cortical symmetry (see Supplementary Figs. S.5, S.10, and S.13).

**Extended Data Fig. E.2.**
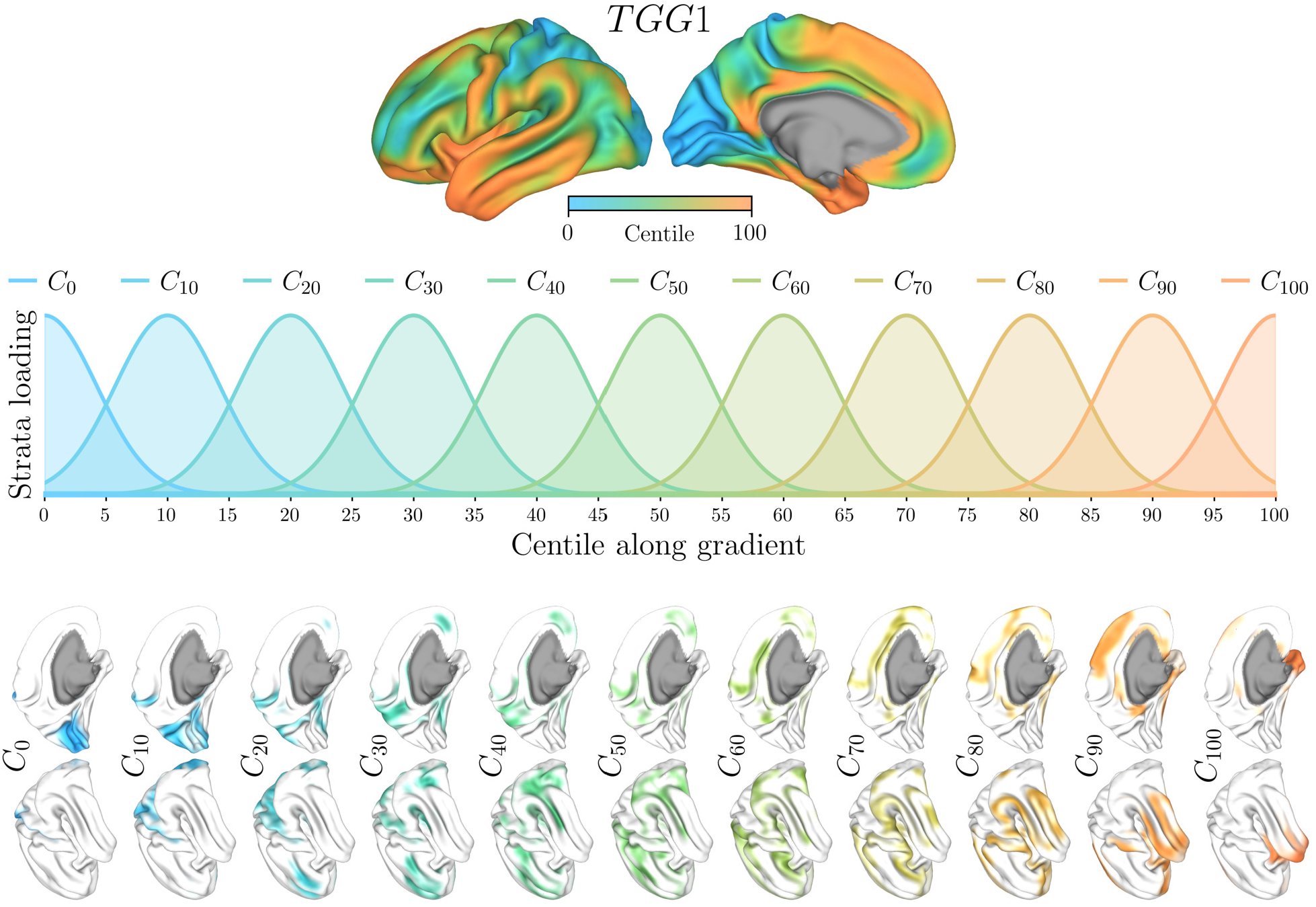
Construction of Probabilistic Gradient Strata along TGG1. The cortical surface is stratified along the first growth gradient (TGG1) into 11 strata, each defined by a target centile, *C* = [0, 10, 20, …, 100], using probabilistic spatial query. **(A)** Vertex-wise TGG1 values transformed to centile space (0–100). **(B)** Eleven target centiles used as reference points along the gradient, with Gaussian-weighted functions defining smooth, overlapping strata centered at each centile. **(C)** Cortical projections illustrating the spatial delineation of each stratum, which are used to extract weighted mean cortical thickness estimates along the gradient.

**Extended Data Fig. E.3.**
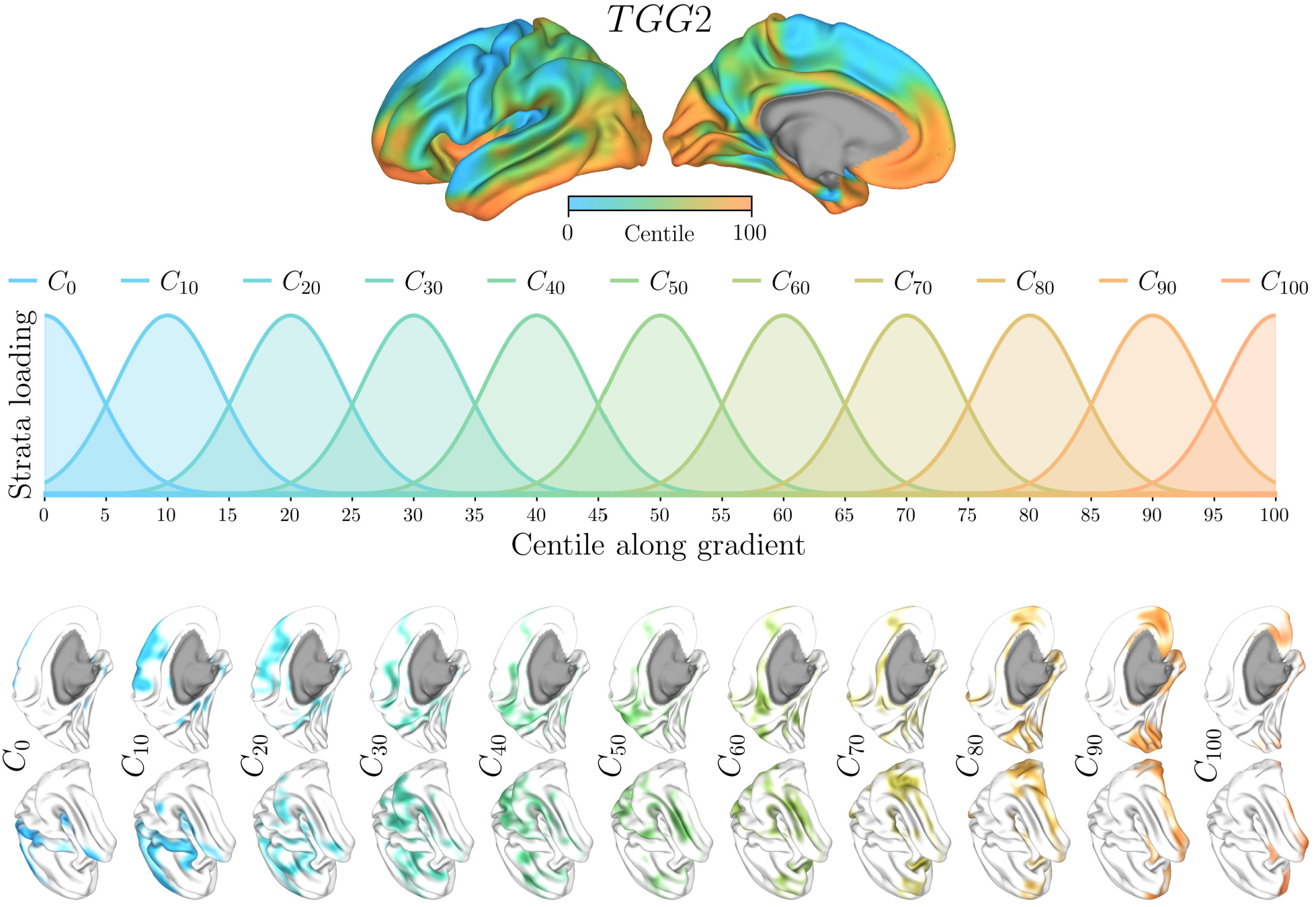
Construction of Probabilistic Gradient Strata along TGG2. The cortical surface is stratified along the second growth gradient (TGG2) into 11 strata, each defined by a target centile, *C* = [0, 10, 20, …, 100], using probabilistic spatial query. **(A)** Vertex-wise TGG2 values transformed to centile space (0–100). **(B)** Eleven target centiles used as reference points along the gradient, with Gaussian-weighted functions defining smooth, overlapping strata centered at each centile. **(C)** Cortical projections illustrating the spatial delineation of each stratum, which are used to extract weighted mean cortical thickness estimates along the gradient.

**Extended Data Fig. E.4.**
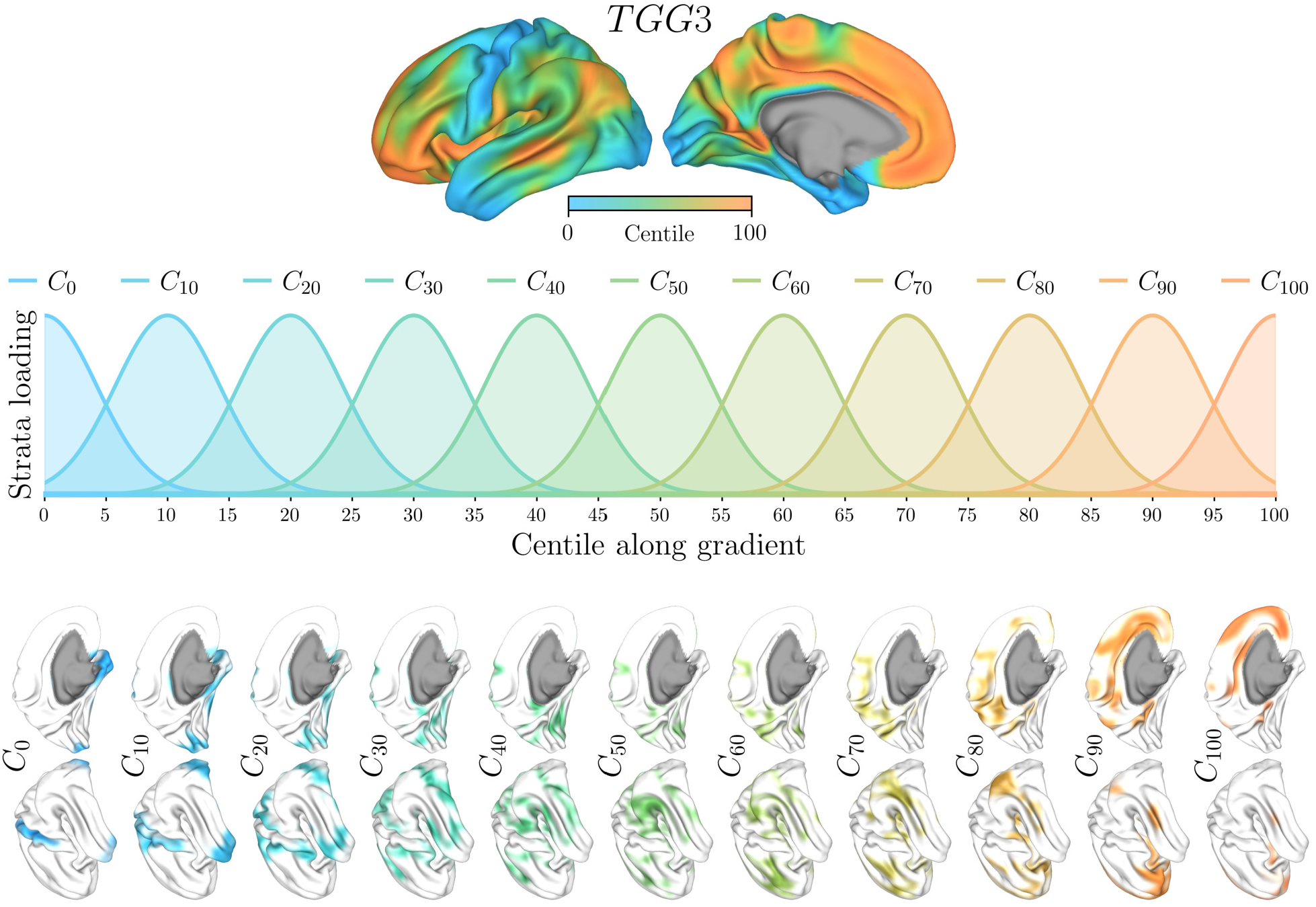
Construction of Probabilistic Gradient Strata along TGG3. The cortical surface is stratified along the third growth gradient (TGG3) into 11 strata, each defined by a target centile, *C* = [0, 10, 20, …, 100], using probabilistic spatial query. **(A)** Vertex-wise TGG3 values transformed to centile space (0–100). **(B)** Eleven target centiles used as reference points along the gradient, with Gaussian-weighted functions defining smooth, overlapping strata centered at each centile. **(C)** Cortical projections illustrating the spatial delineation of each stratum, which are used to extract weighted mean cortical thickness estimates along the gradient.

**Extended Data Fig. E.5.**
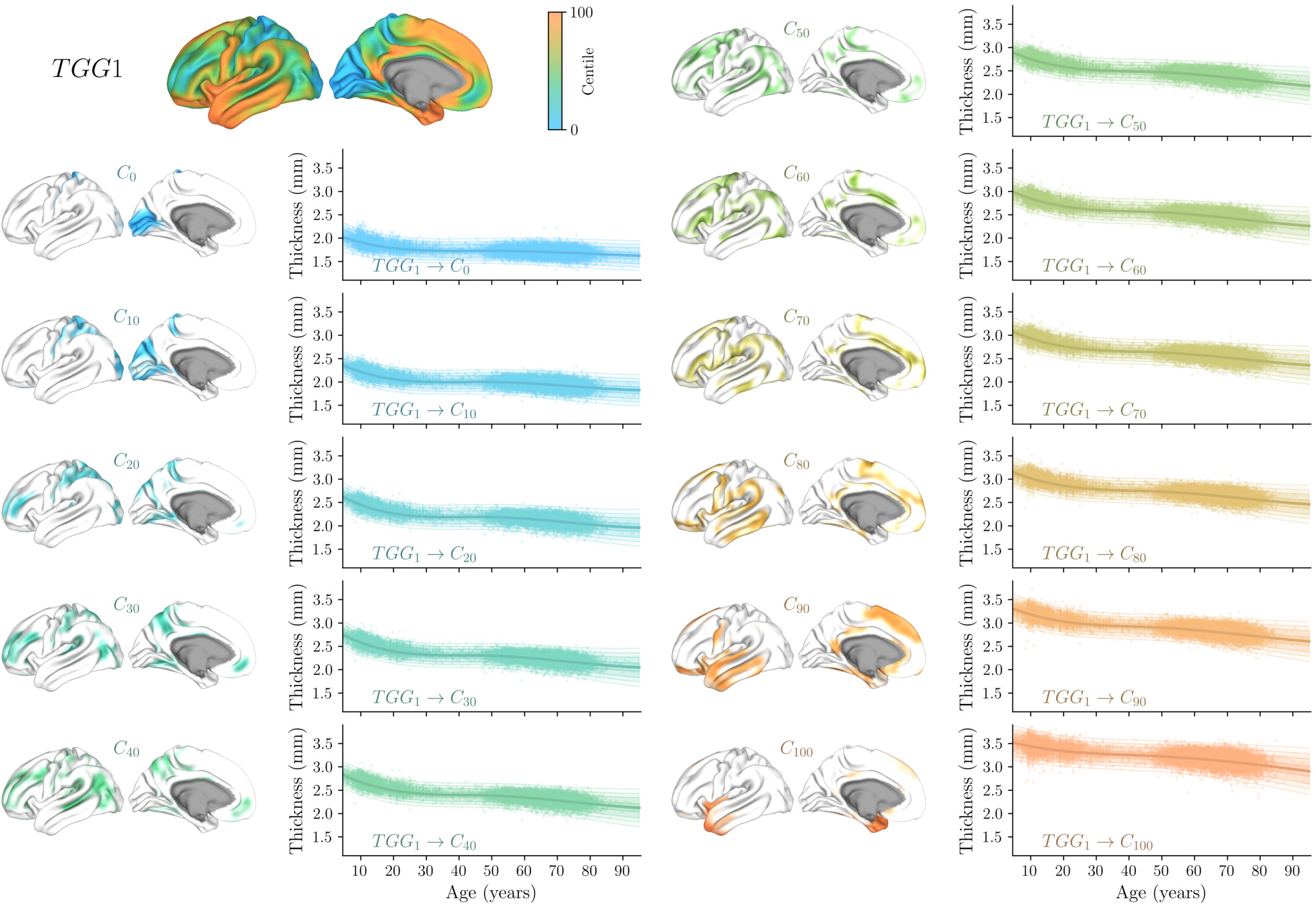
Normative Cortical Thickness Trajectories along TGG1. Normative charts derived from spatial queries defined by probabilistic strata along the first growth gradient (TGG1, shown at the top left). For each of the eleven strata, the corresponding cortical delineation and age-dependent normative trajectory are displayed. Charts depict cortical thickness trajectories estimated using SNM-1000 as a function of age, after accounting for site and sex effects. Solid lines indicate the median estimate, with shaded regions showing the [25, 75], [5, 95], and [1, 99] percentiles. Scatter points represent queried cortical thickness values from held-out test subjects not used for model training. Together, these trajectories illustrate a stable lifelong pattern along TGG1, in which regions that are relatively thicker remain thicker and regions that are thinner remain thinner, with all strata following similar age-related profiles but differing in baseline thickness.

**Extended Data Fig. E.6.**
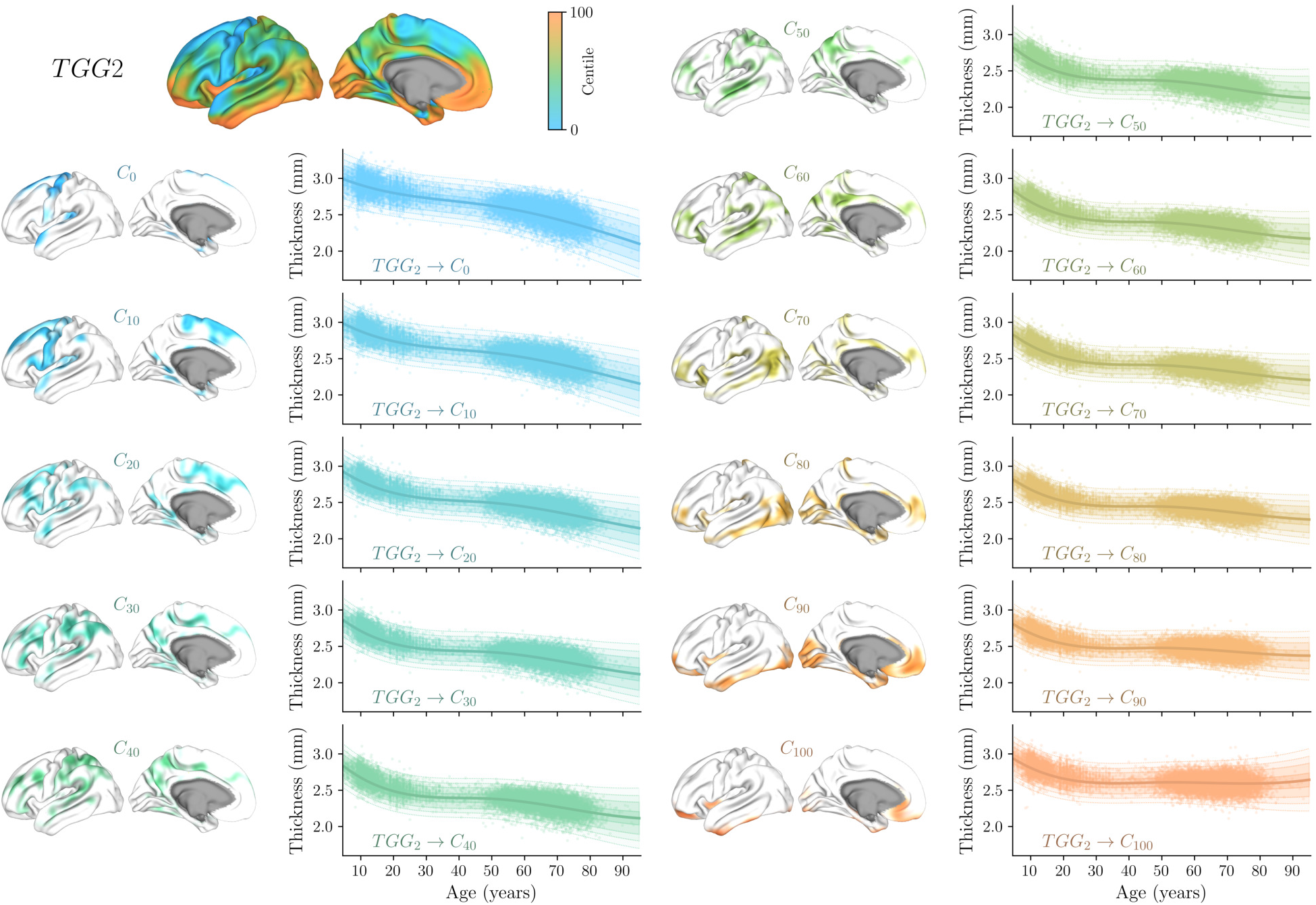
Normative Cortical Thickness Trajectories along TGG2. Normative charts derived from spatial queries defined by probabilistic strata along the second growth gradient (TGG2, shown at the top left). For each of the eleven strata, the corresponding cortical delineation and age-dependent normative trajectory are displayed. Charts depict cortical thickness trajectories estimated using SNM-1000 as a function of age, after accounting for site and sex effects. Solid lines indicate the median estimate, with shaded regions showing the [25, 75], [5, 95], and [1, 99] percentiles. Scatter points represent queried cortical thickness values from held-out test subjects not used for model training. Together, these trajectories highlight the sensitivity of TGG2 to late-life thinning rates. While all strata show largely consistent characteristics in childhood, adolescence, and early adulthood, they diverge primarily after age 30: one end of the gradient shows accelerated thinning (e.g., the superior temporal and precentral gyri), whereas the other end exhibits relative stability and reduced late-life thinning (e.g., visual and ventromedial prefrontal cortices).

**Extended Data Fig. E.7.**
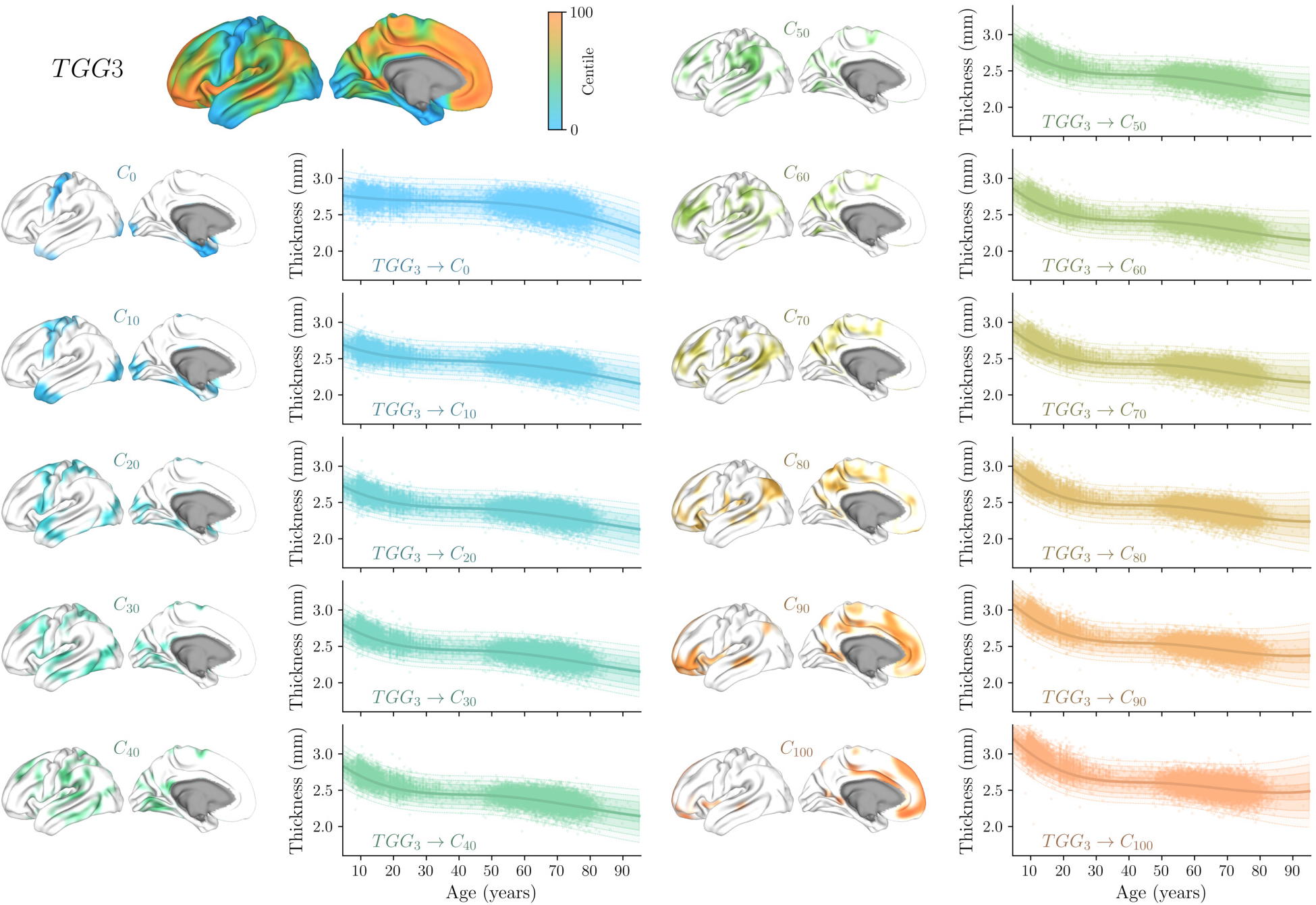
Normative Cortical Thickness Trajectories along TGG3. Normative charts derived from spatial queries defined by probabilistic strata along the third growth gradient (TGG3, shown at the top left). For each of the eleven strata, the corresponding cortical delineation and age-dependent normative trajectory are displayed. Charts depict cortical thickness trajectories estimated using SNM-1000 as a function of age, after accounting for site and sex effects. Solid lines indicate the median estimate, with shaded regions showing the [25, 75], [5, 95], and [1, 99] percentiles. Scatter points represent queried cortical thickness values from held-out test subjects not used for model training. Together, these trajectories illustrate that TGG3 captures variation in early-life cortical pruning rates. Notably, the strata are differentiated by accelerated early-life thinning on one end (e.g., cingulate and ventrolateral cortex), whereas the other end exhibits slower pruning (e.g., precentral gyrus and temporal pole).

**Extended Data Fig. E.8.**
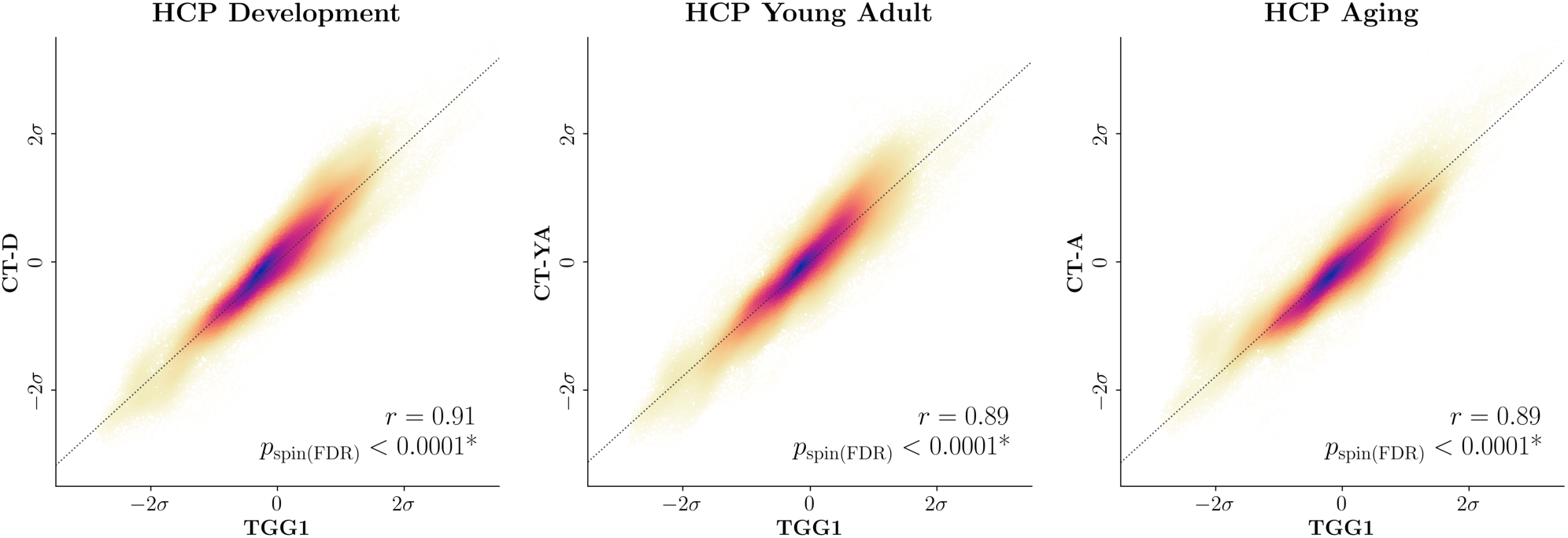
Replication of the TGG1–Thickness Correspondence Across Age-Restricted Cohorts. Correspondence between the first thickness growth gradient (TGG1) and empirical mean cortical thickness maps derived independently from three age-restricted cohorts spanning distinct portions of the lifespan: HCP Development (CT-D; left), HCP Young Adult (CT-YA; middle), and HCP Aging (CT-A; right). For each cohort, individual cortical thickness maps were averaged across participants to generate a cohort-specific mean thickness map (see Section S.1.1 Healthy Lifespan Imaging Data). Scatter plots show vertex-wise associations, with point density indicated by color and dotted lines denoting the identity relationship. TGG1 and thickness values were demeaned and scaled by their standard deviations to facilitate visual comparison. Spatial correspondence was assessed using spin permutations with FDR correction across the three tests. TGG1 closely corresponded with mean cortical thickness in all three cohorts (*r* = 0.91, 0.89, and 0.89, respectively; all *p*_spin(FDR)_ *<* 0.0001), indicating that the thickness hierarchy captured by TGG1 is maintained across the lifespan and is not specific to the age distribution of the training sample. These cohort-derived maps are distinct from the previously established mean thickness map shown in Fig. 4E, which was obtained from the neuromaps repository^38,50^. Abbreviations: TGG, Thickness Growth Gradient; CT, Cortical Thickness; HCP, Human Connectome Project.

**Extended Data Fig. E.9.**
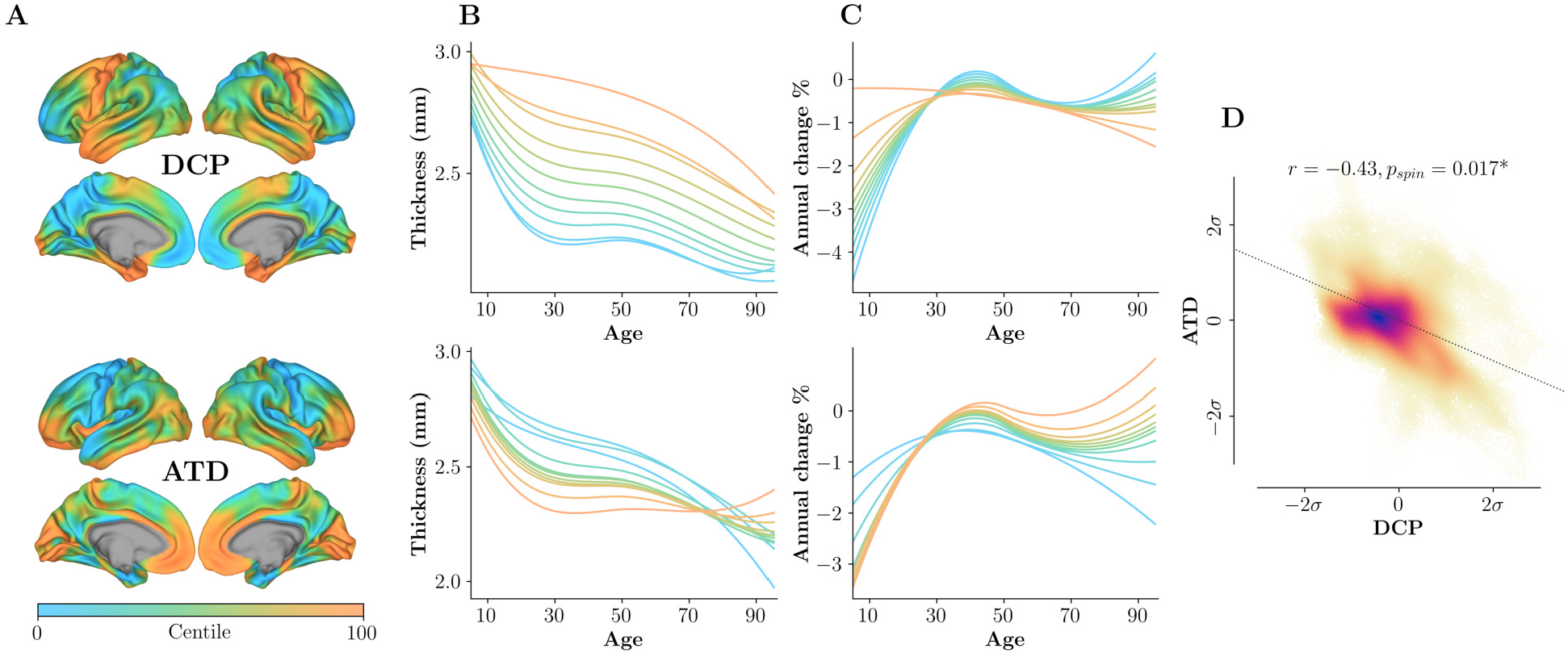
Relationship Between Developmental Cortical Pruning and Aging-Related Thickness Decline. **(A)** Cortical vertex-wise centile maps for developmental cortical pruning (DCP; top), defined as the average annual rate of cortical thickness change between ages 5–20 years, and aging thickness decline (ATD; bottom), defined analogously between ages 40–95 years. **(B)** Median normative cortical thickness trajectories for eleven probabilistic strata sampled along the centiles of each map (centile targets *C* = [0, 10, 20, …, 100]), estimated using SNM-1000. **(C)** Corresponding annual rates of cortical thickness percent change for each stratum. **(D)** Scatter plot showing the association between DCP and ATD across cortical vertices, revealing a significant negative relationship (*r* = −0.43, *p*_spin_ = 0.017). Together, these results indicate that cortical regions exhibiting faster pruning during development tend to show relative preservation from aging-related thinning, suggesting that spatial patterns of cortical maturation and degeneration partially mirror one another, in line with the retrogenesis hypothesis.

## Supplementary material

### S.1 Supplementary Information

#### S.1.1 Healthy Lifespan Imaging Data

Healthy human brain imaging data were aggregated from a large multi-cohort collection of publicly available neuroimaging datasets to capture normative variation across the human lifespan. In total, 30 primary datasets contributed to the healthy reference sample used for model training, encompassing a broad age range and diverse acquisition protocols, resulting in 78,405 individual scans after quality control and participant selection procedures (see Section S.1.2 Sample Inclusion Criteria). Table S.1 summarizes population demographics across the complete samples of each dataset. Two separate CSV files are provided for further details: one reporting demographic characteristics across train and test splits for each dataset (`dataset_demography.csv`), and another providing site- and scanner-specific demographic breakdowns (`cross_site_demography.csv`). The following sections provide dataset-specific details on image acquisition and preprocessing procedures for each contributing cohort.

**Table S.1.**
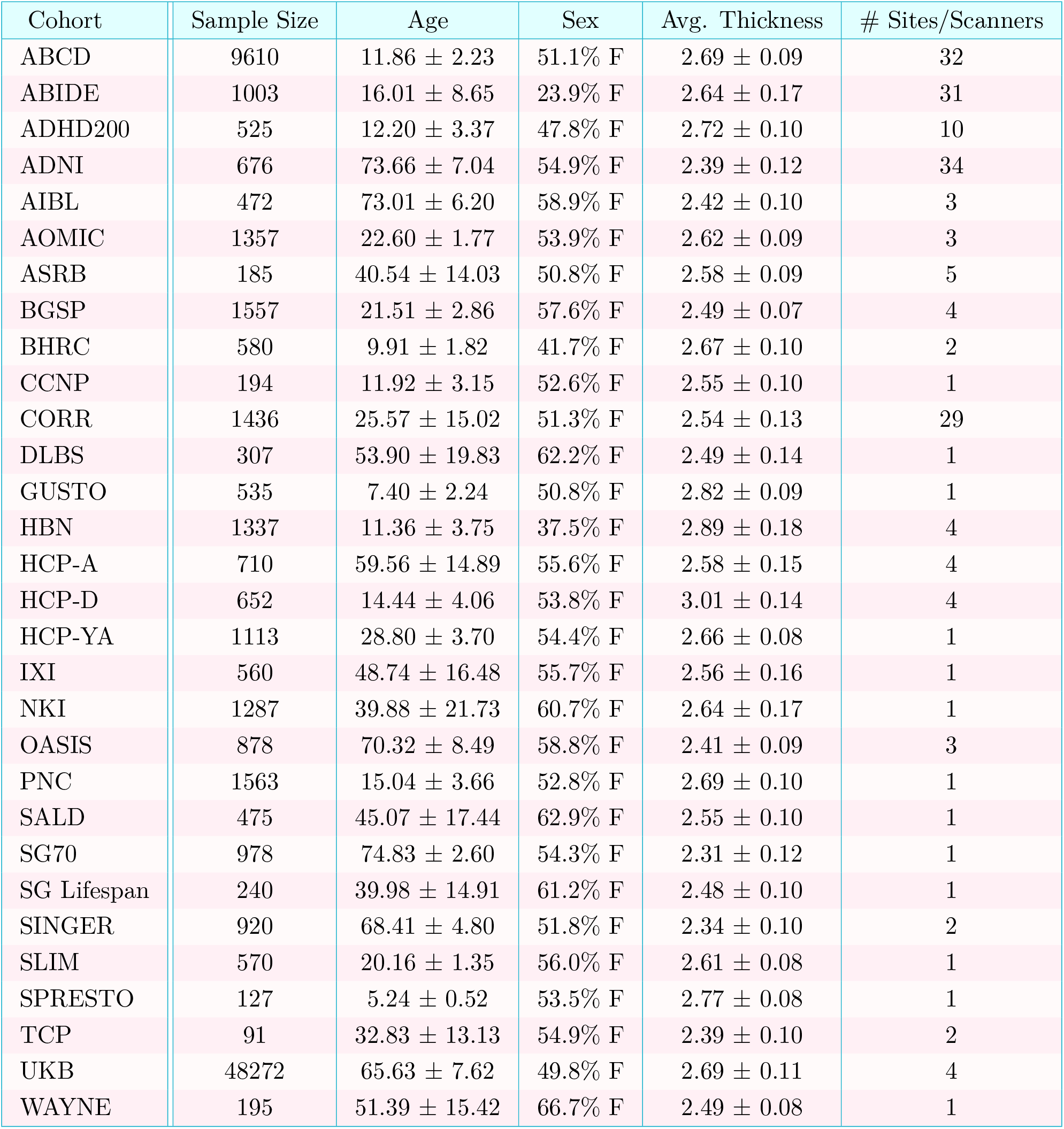
Sample Characteristics of the Healthy Reference Dataset. Values are presented as *µ* ± *σ*, indicating the mean (*µ*) and standard deviation (*σ*). The Sites / Scanners column indicates the number of unique site/scanner combinations per cohort.

##### S.1.1.1 ABCD Data

The Adolescent Brain Cognitive Development^®^ (ABCD) Study is a landmark, longitudinal investigation of brain development and child health, collected across 21 sites throughout the United States ^1^. Structural MRI data were acquired on three 3T scanner platforms (Siemens Prisma, General Electric 750, and Philips) and T1-weighted images were preprocessed using an automated FreeSurfer pipeline (v7.1.1). Preprocessed FreeSurfer outputs were accessed through the NIH Brain Development Cohorts Data Sharing Platform (NBDC). For inclusion in the healthy reference sample, participants were required to have no history of neurological, psychiatric, developmental, or major medical conditions. Specifically, participants were excluded if parental responses indicated a history of neurological or medical disorders (e.g., cerebral palsy, brain tumor, stroke, epilepsy), psychiatric or developmental conditions (e.g., ADHD, autism spectrum disorder, schizophrenia, intellectual disability, mood or anxiety disorders), or substance use disorders, based on the ABCD screening and risk questionnaire (https://nda.nih.gov/data_structure.html?short_name=abcd_screen01). Additional quality control procedures were applied as detailed in Section S.1.2 Sample Inclusion Criteria. After applying these exclusion criteria, the final reference sample comprised 9,610 individuals (51.1% female) aged 8 to 18 years, spanning 32 unique site/scanner combinations.

##### S.1.1.2 ABIDE Data

The Autism Brain Imaging Data Exchange (ABIDE) initiative has aggregated functional and structural neuroimaging data collected from laboratories worldwide to accelerate research into the neural bases of autism spectrum disorder (ASD)^2,3^. In this study, T1-weighted structural MRI data from ABIDE were used. All structural images were processed using an automated pipeline running `reconall` as implemented in FreeSurfer (v5.3.0). To construct a healthy reference sample, individuals with a reported ASD diagnosis were excluded. After applying additional quality control and participant selection procedures (see Section S.1.2 Sample Inclusion Criteria), the final ABIDE sample comprised 1,003 individuals (23.9% female) spanning ages 6 to 64 years. These data were acquired across 31 distinct site/scanner combinations, reflecting the multi-site and multi-scanner nature of the ABIDE data.

##### S.1.1.3 ADHD200 Data

The ADHD200 Sample is a grassroots, open data-sharing initiative designed to accelerate understanding of the neural basis of attention-deficit/hyperactivity disorder (ADHD) through large-scale, multi-site neuroimaging aggregation and discovery-based science. The initiative publicly released resting-state functional MRI and T1-weighted structural MRI data collected across multiple independent imaging sites, comprising both typically developing individuals and children and adolescents diagnosed with ADHD ^4^. For the present study, only T1-weighted structural MRI data from typically developing participants were considered for inclusion in the healthy reference sample. Structural images were processed using FreeSurfer (v6.0.0) to derive cortical thickness measures, following additional quality control and participant selection procedures (see Section S.1.2 Sample Inclusion Criteria). After exclusion of individuals with an ADHD diagnosis and scans failing quality control, the final ADHD200 sample comprised 525 individuals (47.8% female), spanning ages 7 to 25 years, and distributed across 10 distinct site/scanner combinations.

##### S.1.1.4 ADNI Data

Data used in the preparation of this article were obtained from the Alzheimer’s Disease Neuroimaging Initiative (ADNI) database ^5,6^. ADNI was launched in 2003 as a public-private partnership, led by Principal Investigator Michael W. Weiner, MD. The original goal of ADNI was to test whether serial magnetic resonance imaging (MRI), positron emission tomography (PET), other biological markers, and clinical and neuropsychological assessment can be combined to measure the progression of mild cognitive impairment (MCI) and early Alzheimer’s disease (AD). The current goals include validating biomarkers for clinical trials, improving the generalizability of ADNI data by increasing diversity in the participant cohort, and to provide data concerning the diagnosis and progression of Alzheimer’s disease to the scientific community. For up-to-date information, see https://adni.loni.usc.edu. To date, ADNI has comprised three major phases, ADNI1 (2004–2009), ADNI-GO/2 (2010–2016), and ADNI3 (2017–2023), each incorporating newly enrolled participants as well as individuals continuing from earlier phases. Across these phases, MRI data were acquired using a range of scanner models and imaging protocols, with ADNI1 primarily employing 1.5T scanners and later phases predominantly adopting 3T scanners, contributing to substantial site- and scanner-level heterogeneity.

For the present study, only T1-weighted structural MRI data from cognitively normal (CN) participants were included in the healthy reference sample. Individuals diagnosed with mild cognitive impairment (MCI) or Alzheimer’s disease were excluded. T1-weighted images were obtained from the USC Laboratory of Neuro Imaging (LONI) Image and Data Archive, with accompanying phenotypic and diagnostic information extracted from the ADNIMERGE dataset^7^. All structural images were processed using FreeSurfer (v6.0.0) to derive cortical thickness measures, following standard automated reconstruction procedures and additional quality control steps (see Section S.1.2 Sample Inclusion Criteria). After applying all inclusion, exclusion, and quality control criteria, the final ADNI sample comprised 676 cognitively normal individuals (54.9% female), spanning ages 50 to 93 years, and distributed across 34 distinct site/scanner combinations.

##### S.1.1.5 AIBL Data

The Australian Imaging, Biomarkers and Lifestyle Flagship Study of Ageing (AIBL) is a longitudinal research initiative designed to identify biomarkers, cognitive characteristics, and lifestyle factors associated with healthy aging and the development of Alzheimer’s disease ^8^. Data were collected by the AIBL study group, and the study methodology has been described previously ^9^. For the present study, only T1-weighted structural MRI data from cognitively normal participants (CN) were included. Participants with mild cognitive impairment, Alzheimer’s disease, or missing/uncertain diagnostic status (codes −4, 7, or NaN) were excluded. Structural images were processed using FreeSurfer (v6.0.0) to derive cortical thickness measures, following additional quality control and participant selection procedures (see Section S.1.2 Sample Inclusion Criteria). The final AIBL sample comprised 472 individuals (58.9% female), aged 60 to 92 years, across 3 distinct site/scanner combinations.

##### S.1.1.6 AOMIC Data

The Amsterdam Open MRI Collection (AOMIC) is a multi-cohort initiative providing multimodal MRI data, including structural T1-weighted, diffusion-weighted, and functional (resting-state and task-based) MRI, alongside detailed demographics and psychometric assessments ^10^. AOMIC comprises three datasets: ID1000, PIOP1, and PIOP2. All data were acquired on the same Philips 3T scanner (Philips, Best, The Netherlands), with scanner hardware and software upgrades occurring between studies. Scans were performed using a 32-channel head coil, which was upgraded alongside the dStream version of the scanner for PIOP2. Structural T1-weighted images were processed using FreeSurfer (v6.0.0) to derive cortical thickness measures. For inclusion in the healthy reference sample, only participants with complete data and passing quality control were retained (see Section S.1.2 Sample Inclusion Criteria). The final AOMIC sample consisted of 1,357 healthy individuals (53.9% female), aged 18 to 26 years.

##### S.1.1.7 ASRB Data

The Australian Schizophrenia Research Bank (ASRB) is a multi-site biobank providing clinical, neuroimaging, and genetic data from individuals with schizophrenia and healthy comparison participants ^11^. Structural T1-weighted MRI scans were acquired using Siemens Avanto scanners across five sites in Australia (Melbourne, Sydney, Brisbane, Perth, and Newcastle), employing a harmonized MPRAGE protocol (1 mm isotropic voxels, TR = 1980 ms, TE = 4.3 ms). To quantify inter-site variability, a single individual was scanned at all sites, and a Siemens MRI phantom was used for calibration prior to image acquisition. For the present study, only healthy participants without a diagnosis were included, following additional quality control procedures to exclude scans with gross artifacts or significant motion (see Section S.1.2 Sample Inclusion Criteria). T1-weighted images were processed using FreeSurfer (v6.0.0) without manual edits of cortical surfaces. The final ASRB sample comprised 185 individuals (50.8% female), aged 18 to 65 years, distributed across five site/scanner combinations.

##### S.1.1.8 BGSP Data

The Brain Genomics Superstruct Project (BGSP) Open Access Data Release^12,13^ comprises a carefully-vetted collection of neuroimaging, behavioral, cognitive, and personality data from over 1,500 healthy participants. Each neuroimaging dataset includes one T1-weighted structural MRI acquisition and one or more resting-state functional MRI acquisitions, with accompanying automated quality assessments and pre-computed brain morphometrics. For the present study, T1-weighted structural scans were processed using FreeSurfer (v6.0.0) to derive cortical thickness measures. After additional quality control and participant selection procedures (see Section S.1.2 Sample Inclusion Criteria), the final BGSP sample comprised 1,557 individuals (57.6% female), aged 19 to 35 years, spanning 4 distinct site/scanner combinations.

##### S.1.1.9 BHRC Data

The Brazilian High-Risk Cohort Study for Childhood Psychiatric Disorders (BHRC) is a large, community-based longitudinal study designed to investigate typical and atypical developmental trajectories of psychopathology and cognition in childhood and adolescence^14^. Participants were recruited from state-funded schools in the Brazilian cities of São Paulo and Porto Alegre, comprising both a representative community sample and children at increased familial risk for mental disorders. Neuroimaging data were acquired across two sites using consistent acquisition protocols, with participants re-scanned on the same scanner across follow-up waves. For the present study, BHRC data were accessed indirectly through the Reproducible Brain Charts (RBC) initiative, which harmonizes and redistributes curated neuroimaging datasets to facilitate large-scale normative modeling^15^. RBC-provided structural MRI derivatives, preprocessed using FreeSurfer (v6.0.1) and sMRIPrep^16^ (v0.7.1), were retrieved using datalad^17^. Following quality control and exclusion procedures (see Section S.1.2 Sample Inclusion Criteria), the final BHRC sample comprised 580 healthy individuals (41.7% female), aged 5 to 15 years, spanning 2 distinct site/scanner combinations.

##### S.1.1.10 CCNP Data

The Chinese Color Nest Project (CCNP) is a large-scale, longitudinal neuroimaging initiative with the long-term goal of characterizing normative developmental trajectories of brain structure and function across the lifespan, and of understanding neurobiological changes associated with the emergence of mental health and learning disorders ^18,19^. CCNP comprises three coordinated phases spanning development (devCCNP), maturation (matCCNP), and aging (ageCCNP). The present study makes use of data from the developmental CCNP (devCCNP) cohort, also referred to as “Growing Up in China”, which includes longitudinal neuroimaging data from typically developing children and adolescents aged 6 to 18 years. For the present study, CCNP data were accessed indirectly through RBC ^15^. RBC-provided structural MRI derivatives, preprocessed using FreeSurfer (v6.0.1) and sMRIPrep^16^ (v0.7.1), were retrieved using datalad^17^. Following quality control and exclusion procedures (see Section S.1.2 Sample Inclusion Criteria), the final CCNP sample comprised 194 healthy individuals (52.6% female), aged 6 to 18 years, collected at a single imaging site.

##### S.1.1.11 CORR Data

CORR Data were obtained from the Consortium for Reliability and Reproducibility (CoRR), an open neuroimaging resource established to facilitate the assessment of test–retest reliability and reproducibility in functional and structural connectomics^20^. The present study included 1,436 individuals, aged 6–88 years (51.3% female), acquired across 29 site/scanner combinations. As CoRR aggregates retrospectively collected datasets from multiple international laboratories, the sample spans a wide range of acquisition protocols and study designs. All structural T1-weighted MRI scans were processed using an automated pipeline running `recon-all` as implemented in FreeSurfer (v5.3.0).

##### S.1.1.12 DLBS Data

The Dallas Lifespan Brain Study (DLBS) is a longitudinal, open neuroimaging cohort designed to characterize cognitive and brain changes across the adult lifespan, with a particular emphasis on trajectories of healthy aging ^21,22^. The study includes extensively phenotyped participants spanning early to late adulthood, with comprehensive assessments of cognition, brain structure, and brain function. Neuroimaging data were acquired at a single imaging site using a 3T MRI scanner, ensuring consistency of acquisition across participants and study waves. For the present study, structural T1-weighted MRI data were used. Raw structural images were processed using FreeSurfer (v6.0.0) to derive cortical morphometric measures. Following exclusion of individuals failing quality control procedures (see Section S.1.2 Sample Inclusion Criteria), the final DLBS sample comprised 307 healthy individuals (62.2% female), aged 20 to 89 years.

##### S.1.1.13 GUSTO Data

The Growing Up in Singapore Towards Healthy Outcomes (GUSTO) study is a prospective, longitudinal birth cohort established to investigate early-life determinants of health and neurodevelopment in children growing up in Singapore ^23^. The cohort recruited pregnant women attending first-trimester antenatal ultrasound clinics at the National University Hospital and KK Women’s and Children’s Hospital. Parents were Singapore citizens or permanent residents of Chinese, Malay, or Indian ethnic background. Birth outcomes and pregnancy-related measures were obtained from hospital records, and socioeconomic indicators, including household income, were collected via structured questionnaires administered during pregnancy. The GUSTO study was approved by the National Healthcare Group Domain Specific Review Board and the SingHealth Centralized Institutional Review Board, and written informed consent was obtained from all participating mothers. Children underwent MRI scanning at multiple time points across development. For the present study, structural T1-weighted MRI scans were processed using FreeSurfer (v6.0.0) to derive cortical morphometric measures. Following quality control and exclusion procedures (see Section S.1.2 Sample Inclusion Criteria), the final GUSTO sample comprised 535 healthy children (50.8% female), aged 4 to 11 years.

##### S.1.1.14 HBN Data

The Healthy Brain Network (HBN) is a large-scale, ongoing initiative led by the Child Mind Institute, designed to create and openly share a richly phenotyped biobank to advance understanding of brain development and developmental psychopathology in childhood and adolescence ^24^. The HBN cohort comprises children and adolescents recruited from the New York City area, including both typically developing and help-seeking individuals, and includes extensive psychiatric, behavioral, cognitive, and lifestyle assessments alongside multimodal neuroimaging data. Neuroimaging protocols include structural T1-weighted MRI, diffusion MRI, and functional MRI acquired across multiple imaging sites. For the present study, HBN data were accessed indirectly through RBC ^15^. RBC-provided structural MRI derivatives, preprocessed using FreeSurfer (v6.0.1) and sMRIPrep^16^ (v0.7.1), were retrieved using datalad^17^. Following additional exclusion and quality control procedures (see Section S.1.2 Sample Inclusion Criteria), the final HBN sample comprised 1,337 healthy individuals (37.5% female), aged 5 to 22 years, collected across four imaging sites.

##### S.1.1.15 HCP Aging Data

The HCP Aging (HCP-A) imaging data was sourced from the HCP-Aging Lifespan 2.0 release^25^ (HCPA) which included structural MRI images from 725 healthy adults aged 36 to 100+ years old. For all participants aged above 90 years old, age was reported as 100 to avoid releasing protected health information. Participants with ages above 90 were excluded from the analyses due to the lack of precise age information, resulting in a final sample of 710 individuals (55.6% female). The acquisition and preprocessing protocols for this cohort were generally consistent with the earlier acquired HCP-YA protocols (see Section S.1.1.17 HCP Young Adult Data), with particular necessary changes due to hardware differences that are detailed elsewhere^26^. In summary, data were collected on Siemens 3T Prisma scanners (with 32-channel head coils) located at 4 different acquisition sites in the United States. Particularly, T1w images were collected by a multi-echo MPRAGE sequence with real-time motion correction (0.8mm isotropic voxels, TR = 2500ms, TE = 1.8/3.6/5.4/7.2ms, TI = 1000ms, flip angle = 8°, 6/8 slice partial Fourier)^26^. The preprocessing pipeline for structural images was similar to that of HCP-YA following the minimal preprocessing pipeline^27^ with minor variations including a different version of FreeSurfer (version 6.0) and an update to MSMAll using higher order smoothness constraints^28^.

##### S.1.1.16 HCP Development Data

The development imaging data was sourced from HCP-Development Lifespan 2.0 release^29^ (HCP-D) which contained structural MRI images of 652 healthy participants (53.8% female) aged 5 to 22 years old. This cohort aimed to adapt existing HCP protocols to the practical challenges of studying developmental populations. The acquisition and processing of the HCP-D dataset^29^ were generally consistent with the HCP-A dataset^25^. Namely, similar Prisma scanners (with 32-channel head coils) were used to collect imaging data across four acquisition sites, while a tailored 32-channel head coil was used for 5-7 year-old participants^26,29^. Identical to HCP-A, a multi-echo MPRAGE sequence was used to collect T1w images followed by the same preprocessing procedures^26^.

##### S.1.1.17 HCP Young Adult Data

Young adult imaging data was sourced from the HCP S1200 release^30^ (HCP-YA). This contained structural MRI images obtained from 1,113 healthy participants (54.4% female) aged 22 to 37 years old. A comprehensive report on imaging acquisition and preprocessing is available elsewhere^27^. In brief, the images were acquired with a Siemens 3T Skyra scanner with a 32-channel head coil (housed at Washington University). T1w images were acquired using a single-echo MPRAGE sequence (0.7mm isotropic resolution, TR = 2400ms, TE = 2.14ms, TI = 1000ms, flip angle = 8°, no partial Fourier)^27^. Acquired images underwent preprocessing using the HCP’s minimal preprocessing pipeline. Notably, structural images underwent gradient distortion correction, skull stripping, readout distortion correction, and bias field correction, followed by a nonlinear registration to a standard template volumetric space. Cortical surface reconstruction was performed using FreeSurfer (version 5.2) which produced 3-dimensional meshes for the white and pial cortical surface along with segmentation and folding-based registration to the *fsaverage* surface. Finally, the outputs were registered to the *Conte69* surface template with a multimodal surface matching algorithm (MSMAll)^31^ and downsampled to triangulated meshes with ~32k vertices per cortical hemisphere, i.e. the *fs-LR* 32k space.

##### S.1.1.18 IXI Data

The IXI dataset is an open-access neuroimaging resource comprising structural and multimodal MRI data from nearly 600 healthy adult participants, made publicly available as part of the Information eXtraction from Images (IXI) project (https://brain-development.org/ixi-dataset/). MRI data were collected at three hospitals in London using different scanners, including a Philips 3T system at Hammersmith Hospital, a Philips 1.5T system at Guy’s Hospital, and a GE 1.5T system at the Institute of Psychiatry. The dataset includes T1-, T2-, and proton density–weighted images, as well as diffusion and angiographic sequences. For the present study, only T1-weighted structural MRI scans were considered for inclusion. Structural images were processed using FreeSurfer (v6.0.1) to derive cortical morphometric measures. Following quality control and participant selection procedures (see Section S.1.2 Sample Inclusion Criteria), the final IXI sample comprised 560 healthy individuals (55.7% female), aged 20 to 86 years.

##### S.1.1.19 NKI Data

The Nathan Kline Institute (NKI) Rockland Sample is a large-scale, community-ascertained neuroimaging initiative designed to characterize normative and atypical brain organization across the human lifespan ^32^. The study aims to provide a deeply phenotyped sample spanning childhood through late adulthood, including detailed demographic, behavioral, and neuroimaging measures. Participants were recruited from the United States and assessed under protocols approved by the Institutional Review Board at the Nathan Kline Institute. For the present study, NKI data were accessed indirectly through RBC ^15^. RBC-provided structural MRI derivatives, preprocessed using FreeSurfer (v6.0.1) and sMRIPrep^16^ (v0.7.1), were retrieved using datalad^17^. Following additional exclusion and quality control procedures (see Section S.1.2 Sample Inclusion Criteria), the final NKI sample comprised 1,287 healthy individuals (60.7% female), aged 6 to 86 years, collected at a single imaging site.

##### S.1.1.20 OASIS Data

The Open Access Series of Imaging Studies (OASIS) is a multimodal neuroimaging resource designed to facilitate the study of healthy aging and Alzheimer’s disease across the adult lifespan ^33^. The OASIS initiative aggregates data from several longitudinal and cross-sectional studies conducted through the Charles F. and Joanne Knight Alzheimer Disease Research Center (Knight ADRC) at Washington University in St. Louis, including the Memory and Aging Project, the Adult Children Study, and the Healthy Aging and Senile Dementia study. Imaging data were acquired over multiple years using both 1.5T and 3T MRI scanners across several scanner platforms and acquisition protocols. The present study makes use of data from the OASIS-3 release, which integrates earlier OASIS cohorts into a large, longitudinal dataset spanning mid-to late adulthood. For inclusion in the healthy reference sample, only cognitively normal individuals were retained, excluding participants with Alzheimer’s disease or other forms of dementia. MRI scans and accompanying phenotypic data were downloaded from XNAT Central ^34^. Structural T1-weighted MRI scans were processed using FreeSurfer (v6.0.0) to derive cortical thickness measures, following additional quality control and participant selection procedures (see Section S.1.2 Sample Inclusion Criteria). The final OASIS sample comprised 878 healthy individuals (58.8% female), aged 42 to 96 years, distributed across 3 distinct site/scanner combinations.

##### S.1.1.21 PNC Data

The Philadelphia Neurodevelopmental Cohort (PNC) is a large, community-based neuroimaging initiative designed to characterize brain development and its relationship to cognition, behavior, and mental health across adolescence and young adulthood ^35,36^. Participants were recruited from the Children’s Hospital of Philadelphia care network, forming a diverse developmental sample of children and adolescents residing in the greater Philadelphia area, United States. The study was approved by the Institutional Review Boards of the University of Pennsylvania and the Children’s Hospital of Philadelphia. For the present study, PNC data were accessed indirectly through RBC ^15^. RBC-provided structural MRI derivatives, preprocessed using FreeSurfer (v6.0.1) and sMRIPrep^16^ (v0.7.1), were retrieved using datalad^17^. Following additional exclusion and quality control procedures (see Section S.1.2 Sample Inclusion Criteria), the final PNC sample comprised 1,563 healthy individuals (52.8% female), aged 8 to 23 years, collected at a single imaging site.

##### S.1.1.22 SALD Data

The Southwest University Adult Lifespan Dataset (SALD) is a cross-sectional neuroimaging cohort designed to characterize structural and functional brain changes across adulthood^37^. The dataset comprises a community-based adult lifespan sample and includes multimodal neuroimaging and basic phenotypic information. Data were collected at a single imaging site at the Center for Brain Imaging, Southwest University, under locally approved research protocols. For the present study, structural T1-weighted MRI data from SALD were used. All scans were processed using FreeSurfer (v6.0.0). Following additional exclusion and quality control procedures (see Section S.1.2 Sample Inclusion Criteria), the final SALD sample comprised 475 healthy individuals (62.9% female), aged 19 to 80 years, collected at a single site.

##### S.1.1.23 SG70 Data

The SG70 study was conducted as a subcohort study nested within the Singapore Chinese Health Study ^38^ to facilitate an in-depth investigation of multi-organ and multi-system aging among a sub-group of older adults in their seventies. In total, 1,140 participants aged 68 to 82 years were enrolled during follow-up assessments from 2022 to 2025 ^39^. Participants underwent comprehensive MRI brain imaging and cognitive assessments covering multiple domains at a single study location. For the current analysis, T1-weighted structural MRI scans were processed using FreeSurfer (v5.3.0). After implementing exclusion and quality control protocols (see Section S.1.2 Sample Inclusion Criteria), data from 978 individuals (54.3% female) from the SG70 study were included in this analysis. The study received approval from both the National University of Singapore Institutional Review Board (NUS-IRB, 2020/398) and the Domain Specific Review Board (DSRB, 2020/01465), and was conducted in accordance with the Human Biomedical Research Act (HBRA) and ICH-GCP guidelines.

##### S.1.1.24 SG Lifespan Data

The SG Lifespan dataset is a multimodal, community- and staff-based cohort established by the Multimodal Neuroimaging in Neuropsychiatric Disorders Laboratory (MNNDL) to investigate how brain structure, activity, and connectivity underlie perception and cognition across the lifespan. Data collection includes neuroimaging, physiological and psychological characteristics, cognitive and behavioral assessments, blood biomarkers, and digital health tracking (e.g., sleep and activity monitoring) across different age cohorts. The study recruited three sub-cohorts: young adults (21–30 years) and middle-aged adults (40–80 years) from the community, and NUS staff participants referred via the NUS1000 Staff Edition study in collaboration with the Sleep and Cognition Lab. For the present study, structural T1-weighted MRI scans were processed using FreeSurfer (v5.3.0). Following exclusion and quality control procedures (see Section S.1.2 Sample Inclusion Criteria), the final SG Lifespan sample comprised 240 healthy individuals (61.2% female), aged 21 to 73 years, all scanned at a single imaging site.

##### S.1.1.25 SINGER Data

The SINGER (Singapore Intervention Study to Prevent Cognitive Impairment and Disability) study is a multi-center, two-year clinical trial evaluating the effects of multidomain lifestyle interventions on cognitive decline and brain health in older adults at risk of dementia ^40^. It represents Singapore’s adaptation of the Finnish Geriatric Intervention Study to Prevent Cognitive Impairment and Disability (FINGER) trial, following the same multidomain dementia-prevention framework but tailored to an Asian population. For the present study, structural T1-weighted MRI scans were processed using FreeSurfer (v5.3.0). After exclusion and quality control procedures (see Section S.1.2 Sample Inclusion Criteria), the final sample comprised 920 healthy older adults (51.8% female), aged 60–80 years, scanned across two imaging sites: Nanyang Technological University and the National University of Singapore. The study was approved by the National Healthcare Group Domain-Specific Review Board and conducted in accordance with Good Clinical Practice guidelines, and is registered on ClinicalTrials.gov (NCT05007353).

##### S.1.1.26 SLIM Data

The Southwest University Longitudinal Imaging Multimodal (SLIM) dataset is a community-based cohort designed to map brain structure and function and their associations with behavior in young adults ^41^. The dataset includes multimodal MRI scans, structural, diffusion, and resting-state functional MRI, alongside detailed behavioral assessments covering demographic, cognitive, and emotional measures, collected over a long-term retest duration of approximately three and a half years. Structural T1-weighted scans were processed using FreeSurfer (v5.3.0). Following exclusion and quality control procedures (see Section S.1.2 Sample Inclusion Criteria), the final SLIM sample comprised 570 healthy individuals (56.0% female), aged 16 to 28 years, collected at a single imaging site.

##### S.1.1.27 SPRESTO Data

The Singapore PREconception Study of long-Term maternal and child Outcomes (SPRESTO) is a large, prospective, longitudinal, multi-wave observational birth cohort designed to investigate the influence of preconception and early-life factors on maternal and child health outcomes ^42,43^. Women planning pregnancy were recruited in Singapore between 2015 and 2017 through KK Women’s and Children’s Hospital and community outreach, and mother–child dyads have been followed longitudinally from preconception through early childhood with extensive phenotypic, clinical, and developmental assessments. Neuroimaging data were acquired as part of longitudinal follow-up assessments during early childhood. For the present study, we made use of structural MRI data collected at the preschool-age follow-up. Structural T1-weighted scans were processed using FreeSurfer. Following exclusion and quality control procedures (see Section S.1.2 Sample Inclusion Criteria), the final SPRESTO sample comprised 127 healthy children (53.5% female), aged 4 to 7 years, collected at a single imaging site.

##### S.1.1.28 TCP Data

The Transdiagnostic Connectome Project (TCP) is a richly phenotyped neuroimaging dataset designed to investigate brain organization and transdiagnostic dimensions of psychopathology across adulthood ^44^. The study includes comprehensive behavioral, cognitive, and clinical assessments alongside multimodal MRI data and was conducted across two imaging sites in the United States. MRI data were acquired using harmonized Siemens 3T Prisma scanners with standardized acquisition protocols across sites. For the present study, we made use of structural T1-weighted MRI data from the healthy comparison group. Structural scans were processed using FreeSurfer. Following exclusion and quality control procedures (see Section S.1.2 Sample Inclusion Criteria), the final TCP sample comprised 91 healthy individuals (54.9% female), aged 18 to 68 years, collected across two imaging sites (Yale University and McLean Hospital).

##### S.1.1.29 UKB Data

The UK Biobank (UKB) is a large-scale population-based cohort providing extensive phenotypic, genetic, and neuroimaging data from middle-aged and older adults in the United Kingdom ^45,46^. Neuroimaging data were acquired across four scanner–site combinations using harmonized protocols ^47^. For the present study, we used structural MRI data accessed from UKB application `60698`. Specifically, we made use of preprocessed FreeSurfer-derived structural outputs provided directly by UK Biobank showcase (compressed bulk file, Data-Field 20263), which include T1-weighted cortical surface models and associated structural segmentations generated using standardized UKB processing pipelines. Individuals were included as healthy participants based on responses to the UKB mental health questionnaire (Data-Field 20544), retaining only those who reported no history of mental health disorders diagnosed by a health professional. After exclusion and quality control procedures (see Section S.1.2 Sample Inclusion Criteria), the final UKB sample comprised 48,272 individuals (49.8% female), aged 44 to 86 years, collected across four scanner–site combinations.

##### S.1.1.30 WAYNE Data

The Wayne State dataset was obtained from the Brain Aging in Detroit Longitudinal Study and made available through the 1000 Functional Connectomes Project (https://fcon_1000.projects.nitrc.org/indi/retro/wayne_11.html). This longitudinal study was designed to investigate brain and cognitive changes across the adult lifespan, with a focus on identifying factors influencing age-related trajectories of brain structure and function^48^. Participants were screened at baseline using a health questionnaire to exclude individuals with cardiovascular, neurological, psychiatric, endocrine, or thyroid disorders, use of centrally acting medications, excessive alcohol consumption, head injury with loss of consciousness, and other factors that could confound brain aging effects. Cognitive screening included the Center for Epidemiologic Studies Depression Scale (CES-D) and Mini-Mental State Examination (MMSE) to exclude participants with significant depressive symptomatology or cognitive impairment. Neuroimaging data were acquired at a single site using a 1.5T Siemens Magnetom scanner, with T1-weighted MPRAGE anatomical images collected across multiple waves of data acquisition. For the present study, structural T1-weighted MRI scans were processed using FreeSurfer (v6.0.0). Following exclusion and quality control procedures (see Section S.1.2 Sample Inclusion Criteria), the final WAYNE sample comprised 195 healthy individuals (66.7% female), aged 18 to 78 years, collected at a single imaging site.

**Supplementary Fig. S.1.**
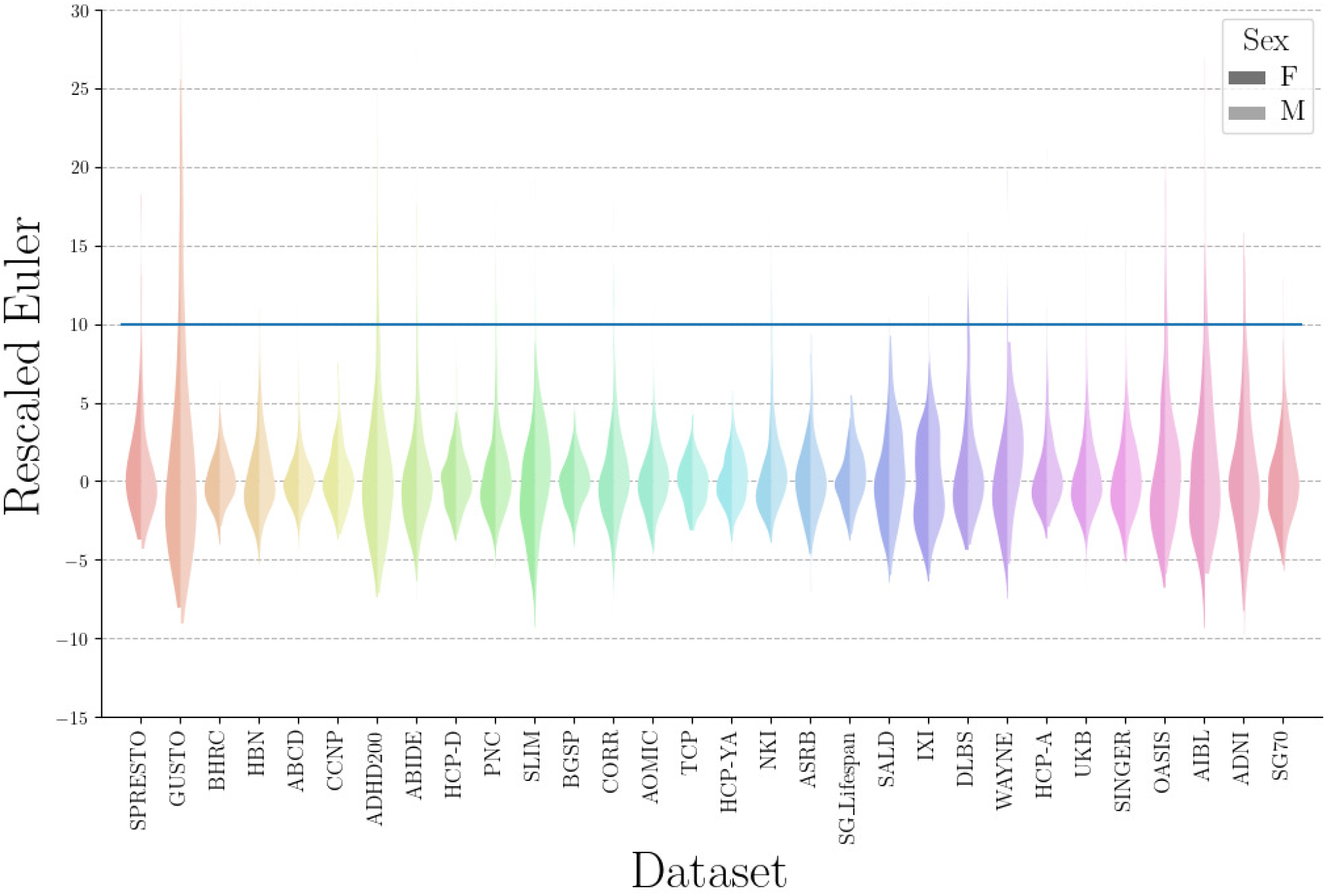
Quality Control of Cortical Surface Reconstruction. Violin plots show the distribution of rescaled Euler numbers across datasets. For each scan, the negative Euler number was square-root transformed and median-centered within dataset. Higher values indicate lower surface reconstruction quality. Each violin corresponds to one dataset. An exclusion threshold of 10 (dashed horizontal line) was applied to remove low-quality reconstructions, following established recommendations.

#### S.1.2 Sample Inclusion Criteria

Following dataset-specific exclusions (e.g., removal of individuals with clinical diagnoses where applicable), additional quality control and exclusion steps were applied. For datasets with multiple scans per individual (i.e., longitudinal follow-ups, such as ABCD, GUSTO, and UKB), a single scan per subject was randomly selected to maintain an inclusive age range while preventing overrepresentation of participants with repeated measures. Scanner–site combinations comprising fewer than 15 participants were excluded to ensure reliable estimation of site-specific effects. In line with previously established recommendations ^49,50^, low-quality surface reconstructions were further filtered based on the Euler number: for each scan, the negative Euler number was square-root–transformed and median-centered within each site, and scans with a resulting rescaled Euler number greater than 10 were excluded (see Supplementary Fig. S.1). This conservative, automated quality control procedure resulted in a final healthy sample of *N*_*p*_ = 78,405 individuals, which was subsequently divided into training and test splits.

#### S.1.3 Stratified Sample Splitting

As described in the Methods section, train test splitting was performed by a randomized approach that controls for the distribution of sample covariates (age, sex, and dataset/batch). In particular, this was implemented by `scikit-learn’s` train_test_split function by utilizing the `stratify` parameter. First, continuous values for individual age were grouped into bins by Quantile-based discretization. Next, discrete values encoding age, sex, and dataset information were utilized for stratified splitting. Supplementary Fig. S.2 demonstrates the stability of covariate distribution after train test splitting.

**Supplementary Fig. S.2.**
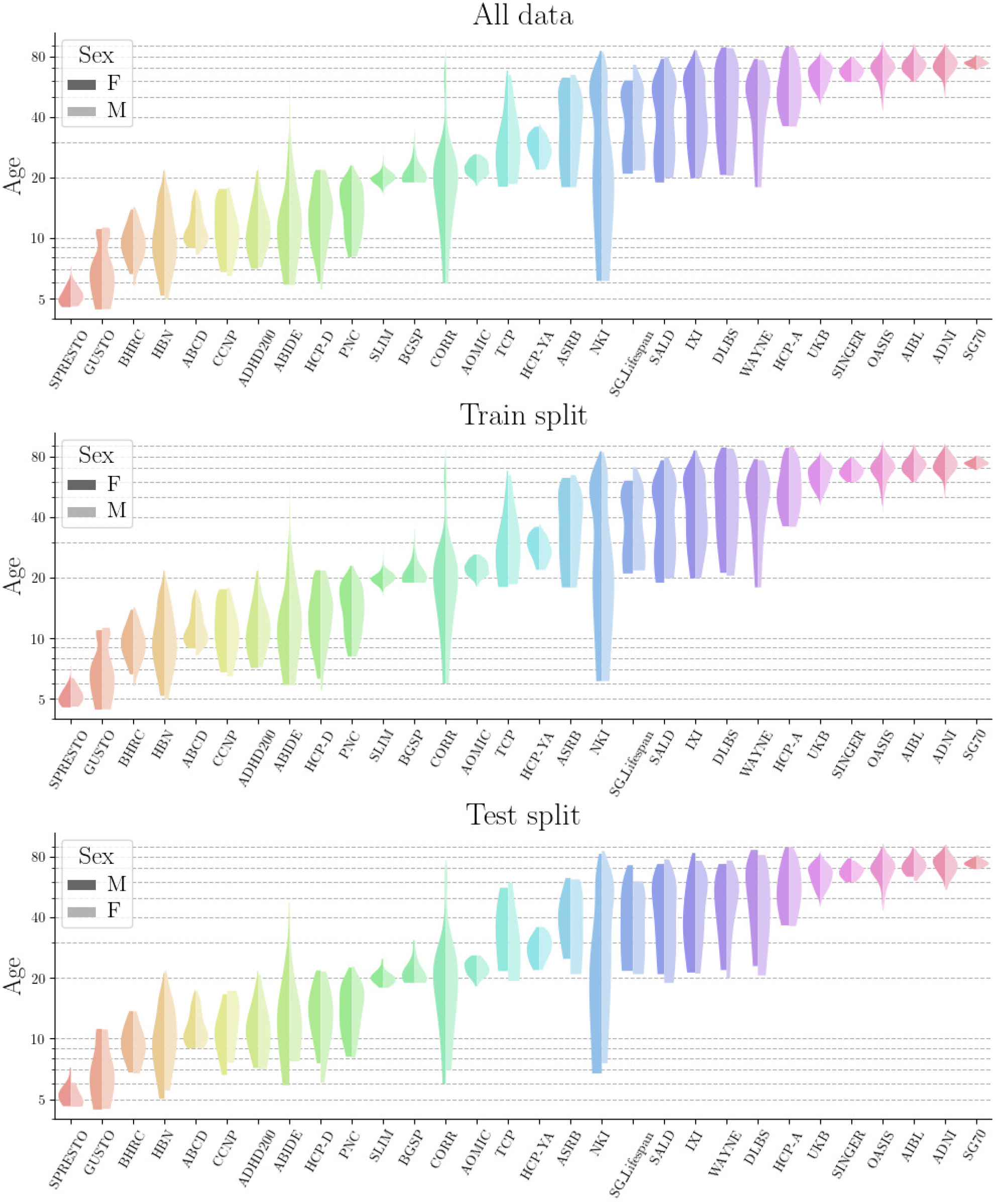
Distribution of Demographic Information Before and After Splitting. The distribution of demographic information is displayed for the entire sample before splitting and after splitting into training and test sets. The violin plots illustrate the age distribution (y-axis) across different datasets (x-axis). Each plot differentiates between female (dark shades) and male (bright shades) participants. The stratified group splitting ensures that the demographic distributions of covariates are matched between the training and test samples.

#### S.1.4 MACC Clinical Imaging Data

The clinical data (part of MACC harmonization study^∗^) was acquired by the Memory, Ageing & Cognition Centre at the National University of Singapore^51,52^. The dataset includes 542 participants aged 50 to 91 years (61.4% female, see Table S.2 for detail), grouped into three cohorts: i) a healthy cohort (HC) of 132 cognitively healthy individuals, ii) a mild cognitive impairment cohort (MCI) of 202 individuals with cognitive impairment but no AD, and iii) an AD cohort of 208 individuals with an AD diagnosis according to the Diagnostic and Statistical Manual of Mental Disorders, Fourth Edition (DSM-IV) criteria. AD diagnosis followed internationally accepted criteria using the National Institute of Neurological and Communicative Disorders and Stroke and Alzheimer’s Disease and Related Disorders Association (NINCDS-ADRDA) guidelines.

Each participant underwent neuroimaging and comprehensive clinical and neuropsychological evaluation, including the Mini-Mental State Examination (MMSE). MRI was performed on a 3T Siemens Magnetom Trio Tim scanner with a 32-channel head coil at the Clinical Imaging Research Centre of the National University of Singapore. The imaging protocol included a T1-weighted MPRAGE sequence (1 mm isotropic resolution, TR = 2300 ms, TE = 1.9 ms, TI = 900 ms, flip angle = 9°, 192 sagittal slices, matrix size = 256 × 256)^53^. T1-weighted scans were preprocessed using FreeSurfer 6.0 `recon-all` pipeline to generate cortical surfaces and thickness estimates in the fsnative surface space^54,55^. Matlab scripts and commands from Connectome Workbench and FreeSurfer were then used to project thickness data to the fs-LR surface space, aligning it with HCP thickness data to enable transfer learning from HCP datasets to MACC.

**Table S.2.**
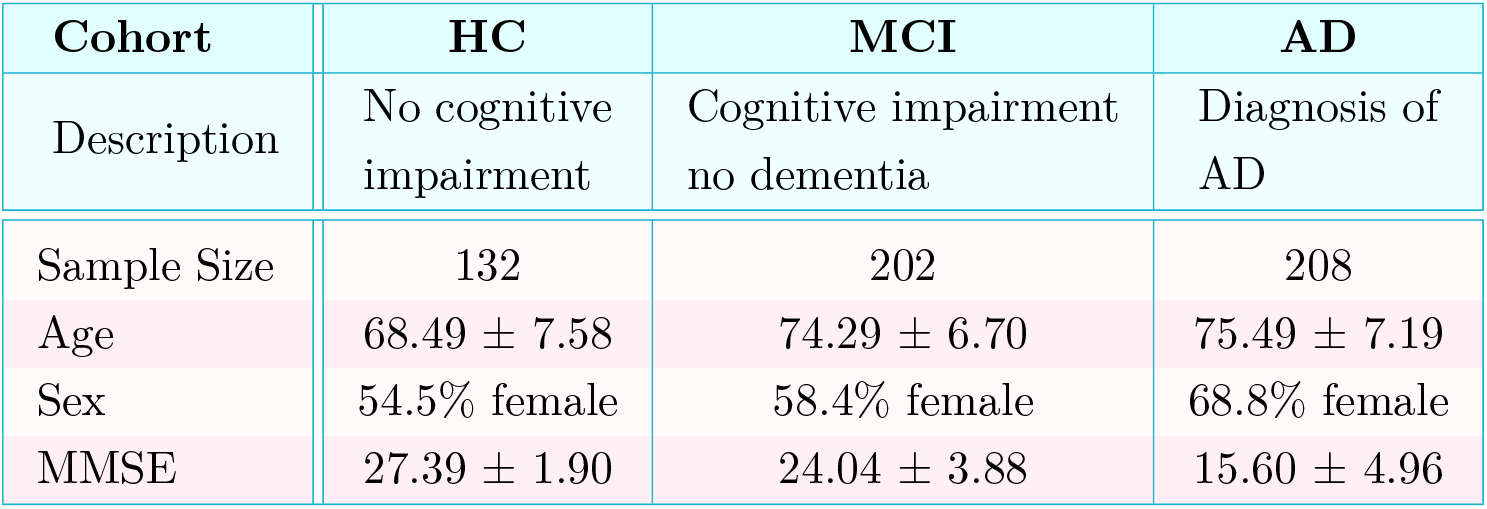
Sample Characteristic of MACC-H dataset. Note: Values presented as *µ* ± *σ* indicate the mean (*µ*) and standard deviation (*σ*).

| Cohort | HC | MCI | AD |
| --- | --- | --- | --- |
| Description | No cognitive impairment | Cognitive impairment no dementia | Diagnosis of AD |
| Sample Size | 132 | 202 | 208 |
| Age | $68.49 \pm 7.58$ | $74.29 \pm 6.70$ | $75.49 \pm 7.19$ |
| Sex | 54.5% female | 58.4% female | 68.8% female |
| MMSE | $27.39 \pm 1.90$ | $24.04 \pm 3.88$ | $15.60 \pm 4.96$ |

#### S.1.5 Cross-Basis Correlation Structure

As described in Section 4.8.3 Spectral Normative Model (SNM), SNM leverages the naturally sparse correlation structure among spectral coefficients to efficiently approximate cross-mode dependencies. Subthreshold correlations are set to zero, reducing computational complexity while preserving key dependencies. Supplementary Fig. S.3 visualizes this sparsity by showing the magnitude of cross-basis correlations for cortical thickness phenotypes. A threshold was applied such that only the top 0.5% of all pairwise correlations were retained, producing a sparse matrix that captures the most relevant dependencies between eigenmodes.

**Supplementary Fig. S.3.**
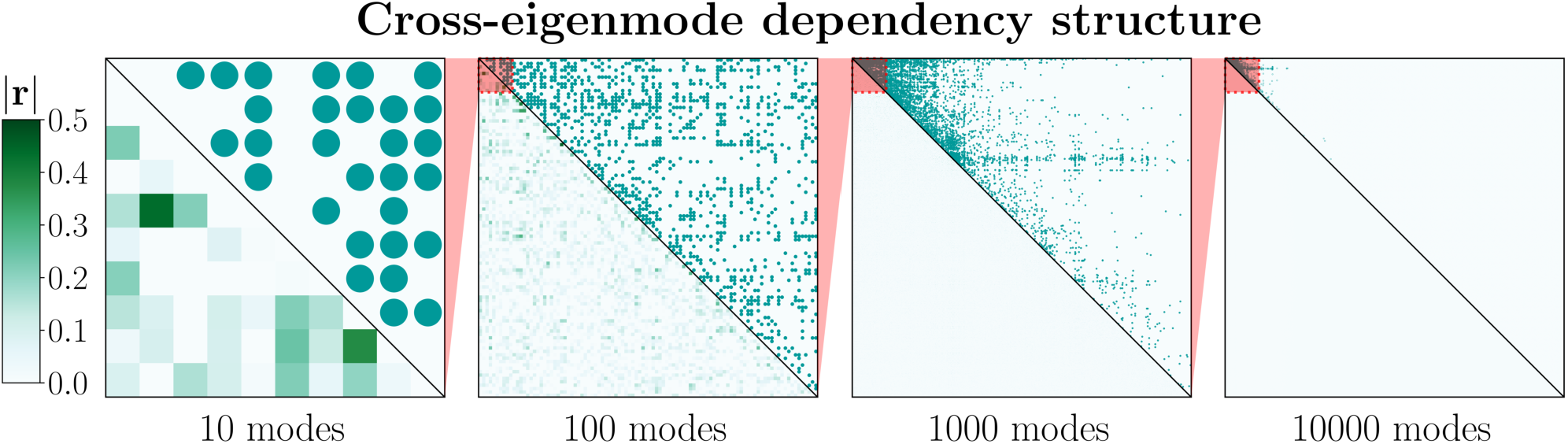
Sparse Cross-Basis Correlation Structure. Magnitudes of pairwise correlations between spectral coefficients encoding cortical thickness phenotypes. Nested heatmaps depict the sparsity of cross-mode dependencies: the lower triangle shows all correlation magnitudes, while the upper triangle highlights suprathreshold correlations retained by the model. The sparsity threshold corresponds to the top 0.5% of correlations, producing a sparse matrix that efficiently captures the most relevant cross-mode relationships.

#### S.1.6 Benchmarking High-Resolution Spatial Queries

Running direct models at full vertex resolution is computationally prohibitive, as a separate model must be fitted for each vertex. To enable a tractable vertex-wise benchmarking evaluation while preserving representative cortical coverage, we randomly sampled 4,000 vertices (2,000 per hemisphere). Left-hemisphere vertices were selected using a randomized distance-based *k*-means subsampling procedure (*k* = 2,000); the vertex-wise symmetry of the fs-LR atlas was then leveraged to include the homotopic right-hemisphere counterparts of these vertices, enabling assessment of asymmetric spatial queries (see Section S.1.8 Sensitivity Analyses for Query Characteristics). Supplementary Fig. S.4 shows the spatial distribution of the selected vertices. This subsample was used exclusively for benchmarking spectral normative model performance (Fig. 2); all other vertex-wise SNM analyses in the manuscript, including growth gradients and clinical applications, use the full vertex set, demonstrating the scalability of SNM to tens of thousands of concurrent spatial queries.

**Supplementary Fig. S.4.**
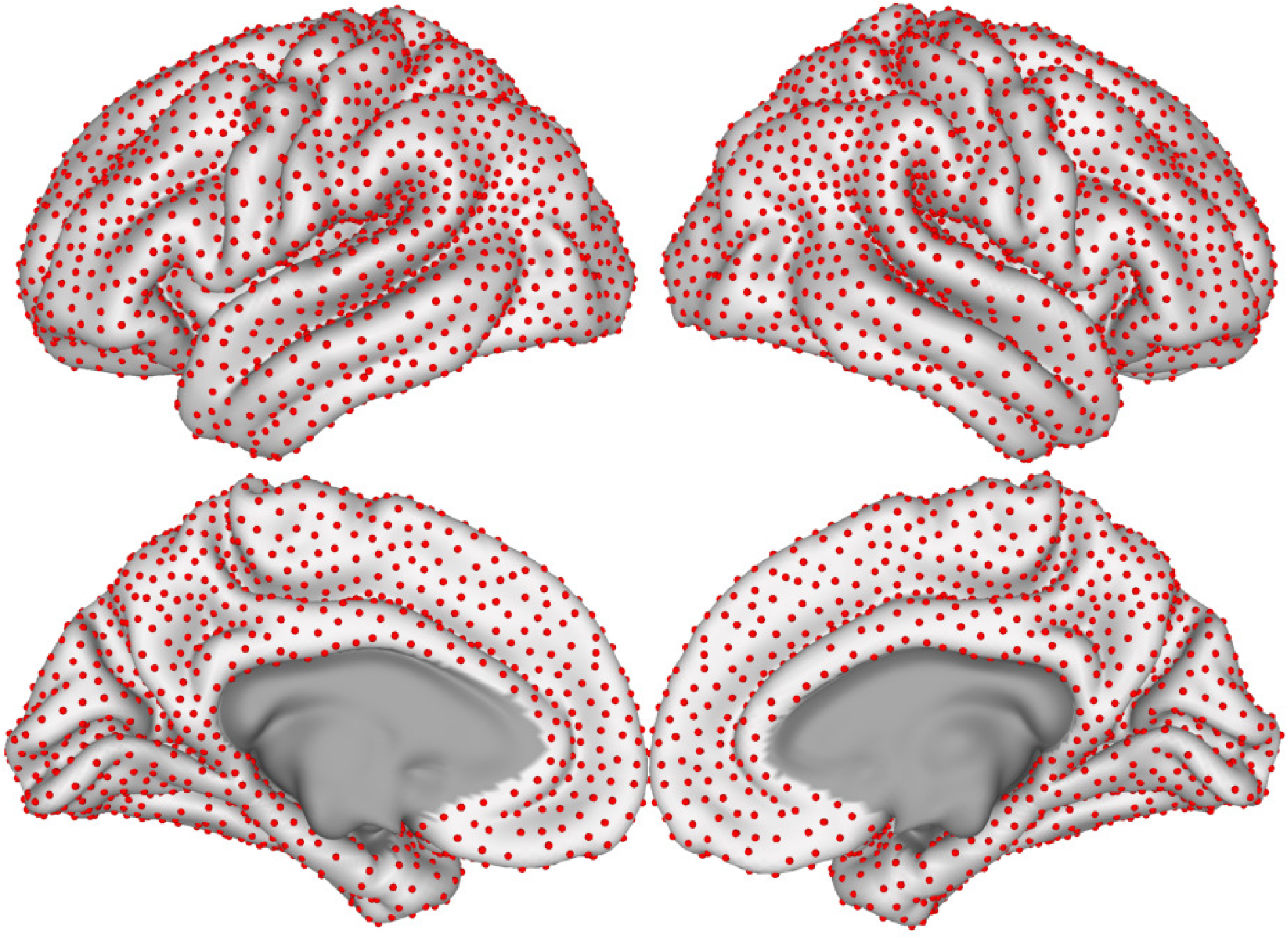
Cortical Projection of Sampled High-Resolution Vertices. The 4,000 vertices used for benchmarking are indicated by red spheres on the fs-LR cortical surface. Vertices were selected using a distance-based subsampling procedure to ensure uniform coverage across the cortex. This selection was used only for benchmarking normative performance; all other SNM analyses in the manuscript use the full vertex set.

#### S.1.7 Cortical Projections of Performance Metrics

Extended Data Fig. E.1 presents a summary of reconstruction accuracy across different families of brain signals, and Fig. 2 summarizes comparative performance between SNM and the direct model. In the following sections, we expand upon these findings, detailing the regional distribution of performance metrics to offer deeper insights into comparative SNM evaluations.

##### S.1.7.1 Reconstruction Accuracy

Extended Data Fig. E.1 quantifies the reconstruction accuracy of different spatial query families in terms of the proportion of signal energy captured as a function of the number of modes. Fig. S.5 extends this analysis by characterizing the regional variability of reconstruction accuracy for brain-wide (Yeo’s 7 functional networks^56^), regional (Yan 200 atlas^57^), and high-resolution (fs-LR 32k vertices) spatial queries (see Section 4.9 Model Evaluation). For each query, the proportion of signal energy captured by the first *k* ∈ {10, 10^2^, 10^3^, 10^4^} modes was projected onto the corresponding cortical locations. These results further demonstrate that the first 10 modes retain only a small fraction of spatial variance across all query types, whereas approximately 100 modes begin to capture a considerable proportion of variance for brain-wide queries. In contrast, regional and high-resolution queries require at least 1,000 modes for accurate reconstruction, with further increases to 10,000 modes yielding diminishing incremental gains in reconstruction accuracy across all signal families.

**Supplementary Fig. S.5.**
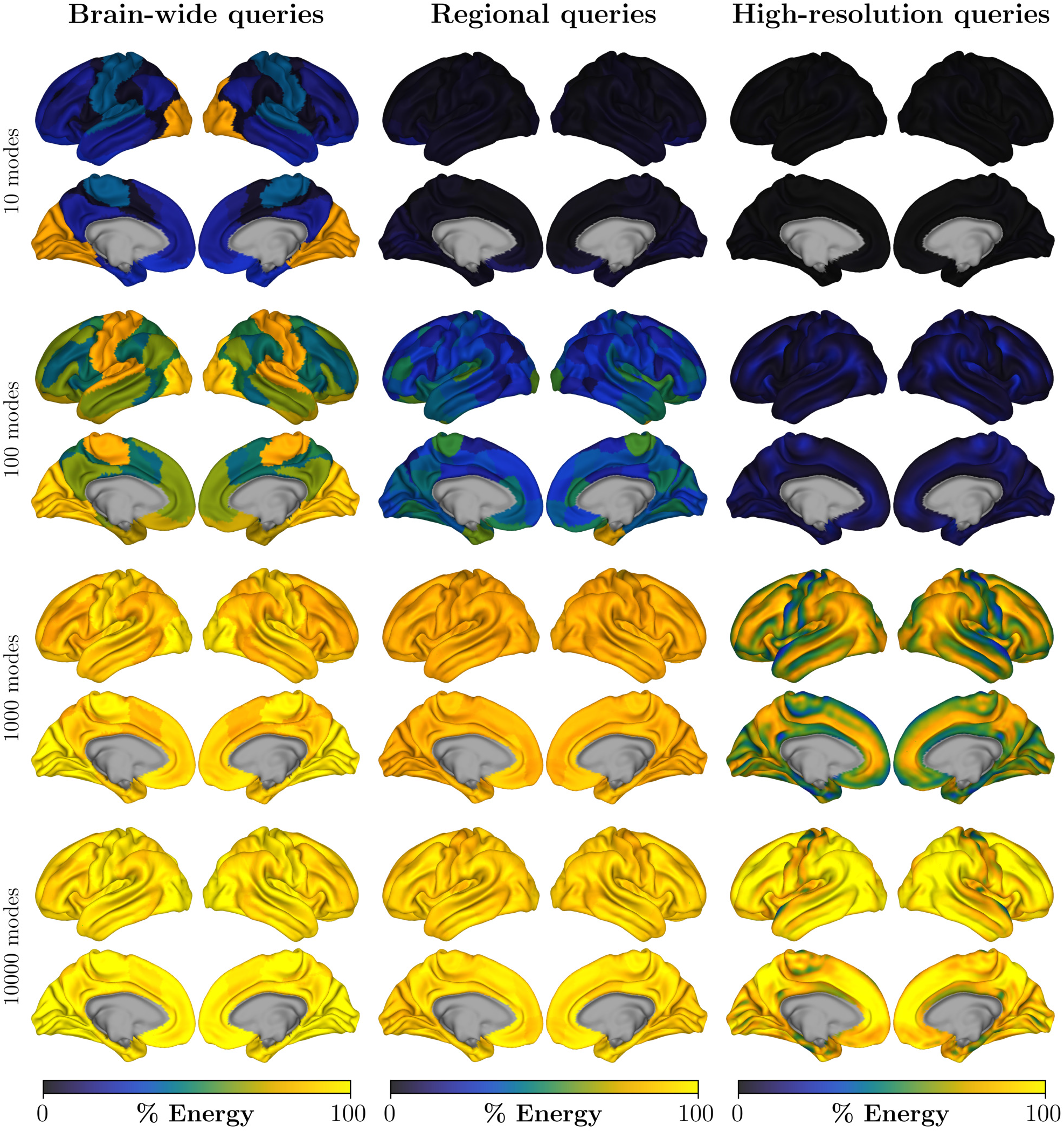
Regional Variability of Reconstruction Accuracy across Spatial Query Families. Reconstruction accuracy is shown for brain-wide (Yeo 7 networks, left column), regional (Yan 200 atlas, middle column), and high-resolution (fs-LR 32k vertices, right column) spatial queries. Rows correspond to spectral reconstructions using *k* = 10, 10^2^, 10^3^, and 10^4^ modes (top to bottom). For each query, reconstruction accuracy is quantified as the proportion of signal energy captured by the first *k* modes and projected onto the cortical location associated with the query.

##### S.1.7.2 Reconstruction Residuals

In Extended Data Fig. E.1, cortical projections of exemplary brain signals were presented alongside the original signal (last four rows). Supplementary Fig. S.6 shows the residuals of low-pass filtered graph approximations of the same signals. For four different low-pass approximations (*k* = 10, 10^2^, 10^3^, 10^4^), the approximation residuals 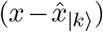 are projected onto the cortical surface. As anticipated, the magnitude of the approximation error (indicated by color intensity in the cortical projections of residuals) decreases as the number of incorporated eigenmodes increases. Furthermore, with the inclusion of more modes, only regions containing high spatial frequency information (such as the transition loci along parcellation borders) have higher approximation errors. This verifies our expectations that low-pass filtering by eigenmodes can accurately approximate smooth signals (i.e. lower graph frequencies).

**Supplementary Fig. S.6.**
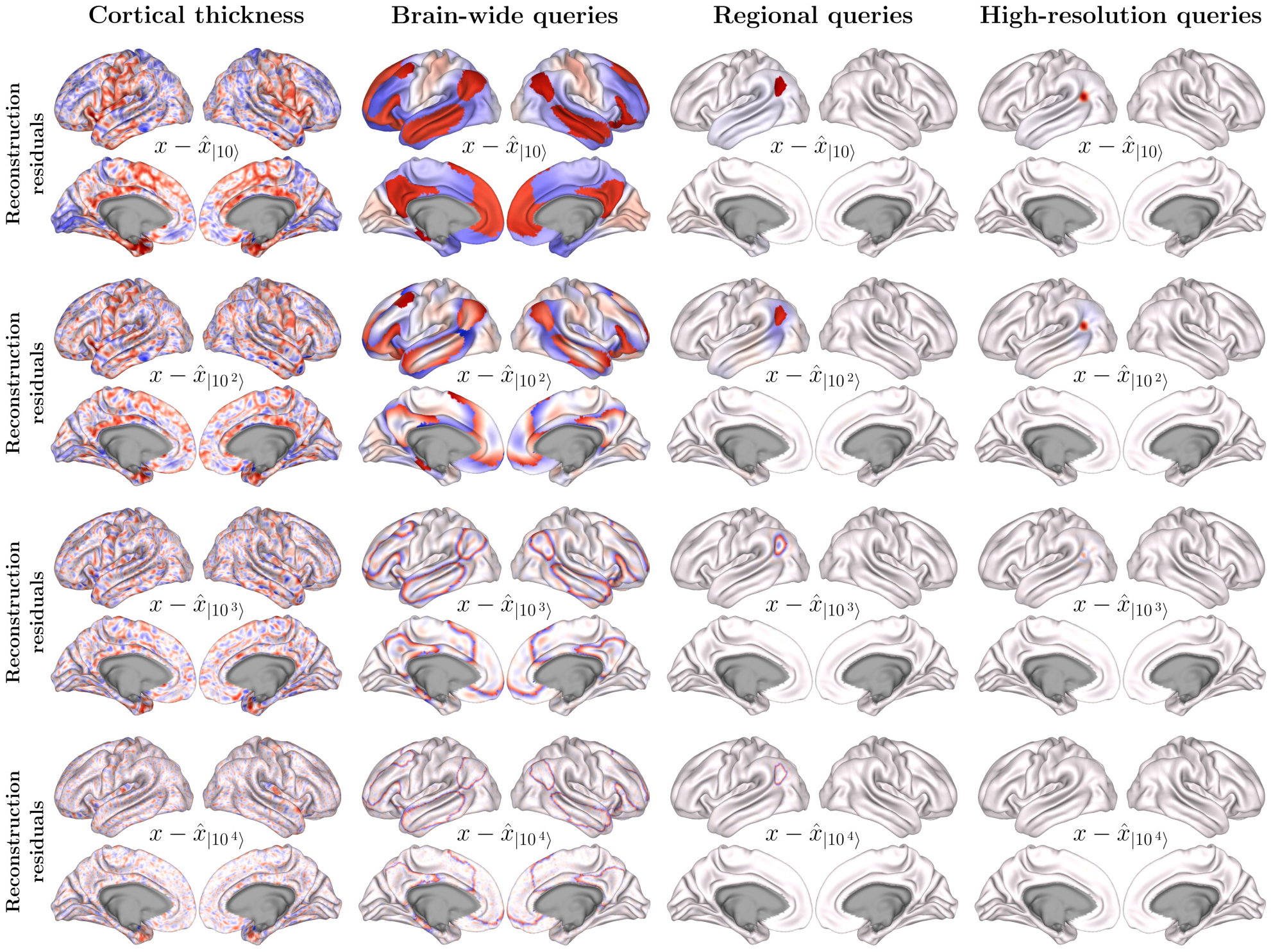
Signal Reconstruction Residuals. The columns (from left to right) display the cortical projections of approximation residuals for individual cortical thickness signals, as well as brain-wide, regional, and high-resolution spatial queries (similar to Extended Data Fig. E.1). The rows show the residuals for the same signal reconstructed using 10, 100, 1,000, and 10,000 eigenmodes, respectively.

##### S.1.7.3 Brain-Wide Normative Estimates

Fig. S.7 presents the performance of various SNMs in reconstructing brain-wide thickness maps, compared to a direct model. The direct model is trained on observed cortical phenotypes for each spatial query (*y* = *T*_train_. *x*), while SNMs rely solely on spectral approximations 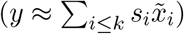. Four SNMs are evaluated, which respectively utilize the first *k* = {10, 10^2^, 10^3^, 10^4^} eigenmodes.

With as few as 10 modes, spectral normative models can produce acceptable normative estimates for some brain-wide signals, indicated by negative MSLL values. However, specific brain-wide signals, such as the average thickness of the ventral attention network and the limbic network (from Yeo’s 7 functional networks^56^), exhibit poor normative estimates at this level, showing noticeably inferior MAE and MSLL scores compared to the direct model. However, including at least 100 modes in the spectral normative model yields performance comparable to the direct model. These findings were also extended in sensitivity analysis for brain-wide asymmetric queries (see Supplementary Fig. S.14).

**Supplementary Fig. S.7.**
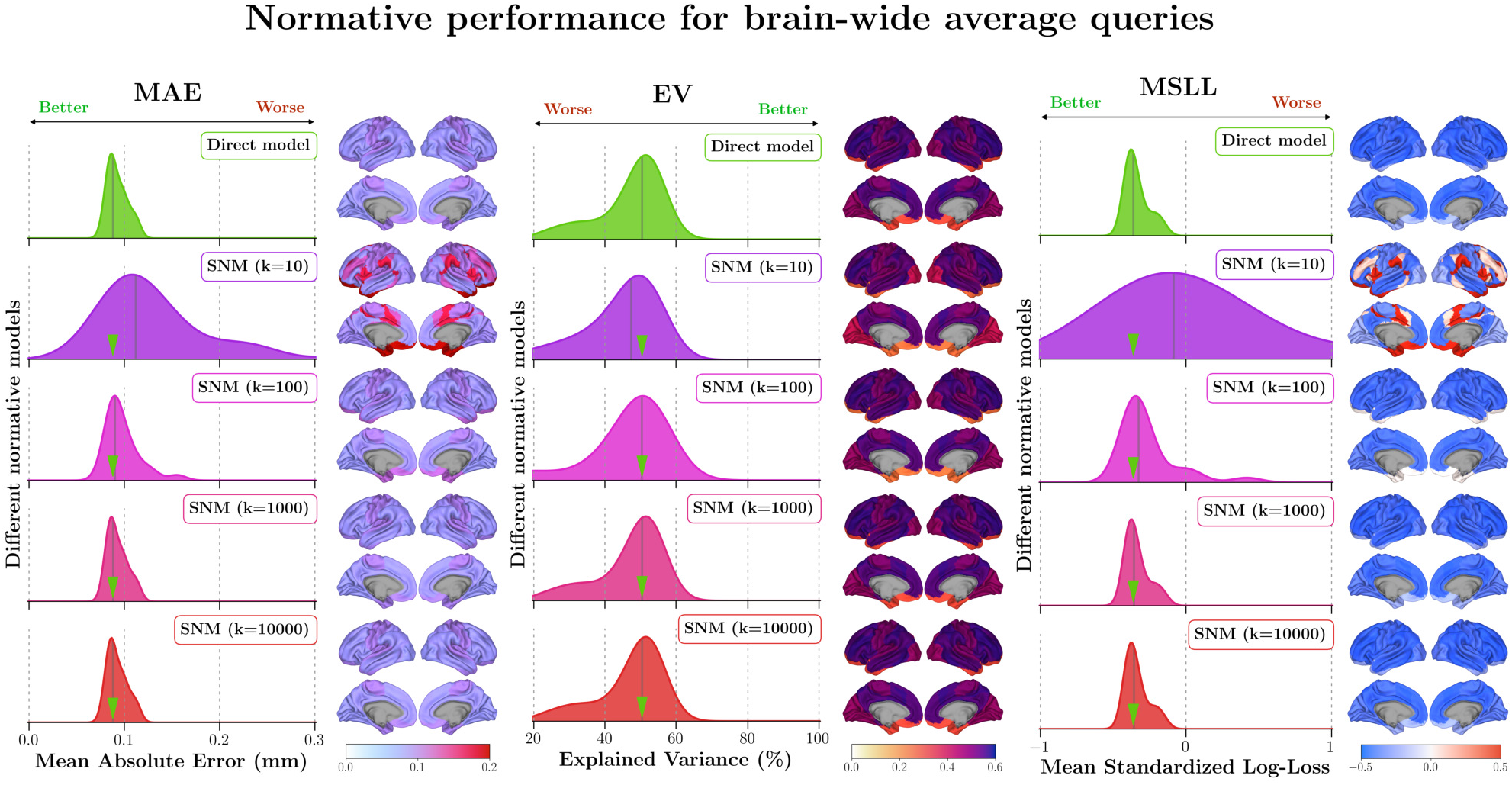
Comparison of Performance for Different Normative Models Assessing Brain-Wide Spatial Queries. Normative performance is evaluated using out-of-sample assessments of direct and spectral normative models. The top row displays the performance of the direct model, while the next four rows show the performance of spectral models with 10, 100, 1,000, and 10,000 eigenmodes, respectively. Mean absolute error (MAE), explained variance (EV), and mean standardized log-loss (MSLL) are used to quantify model performance. Violin plots illustrate the distribution of performance measures across 25 different brain-wide signals, including cortex-wide average thickness and thickness averaged over 7 and 17 functional networks. Median values are indicated by gray lines on the violin plots. A green arrow marks the median performance of the direct model over spectral performance distributions for visual comparison. Cortical surface projections show normative performance values for 7 functional networks, with shared colorbars across all five rows to facilitate visual comparison.

##### S.1.7.4 Regional Normative Estimates

Fig. S.8 extends the previous comparison to regional signals. At this level of spatial specificity, SNM with only 10 modes shows substantially inferior performance across several brain regions. Including the first 100 modes results in acceptable performance for most brain regions, except for regions on the post-central gyrus, insular cortex, and orbitofrontal cortex. Incorporating up to 1000 modes in the spectral model achieves performance matching that of the direct model across all brain regions. Further inclusion of modes (up to 10,000) provides minimal additional benefits in goodness of fit, as measured by MAE, EV, and MSLL. Sensitivity analyses evaluate the robustness of these findings to regional granularity and signal asymmetry (see Supplementary Figs. S.11, S.15).

**Supplementary Fig. S.8.**
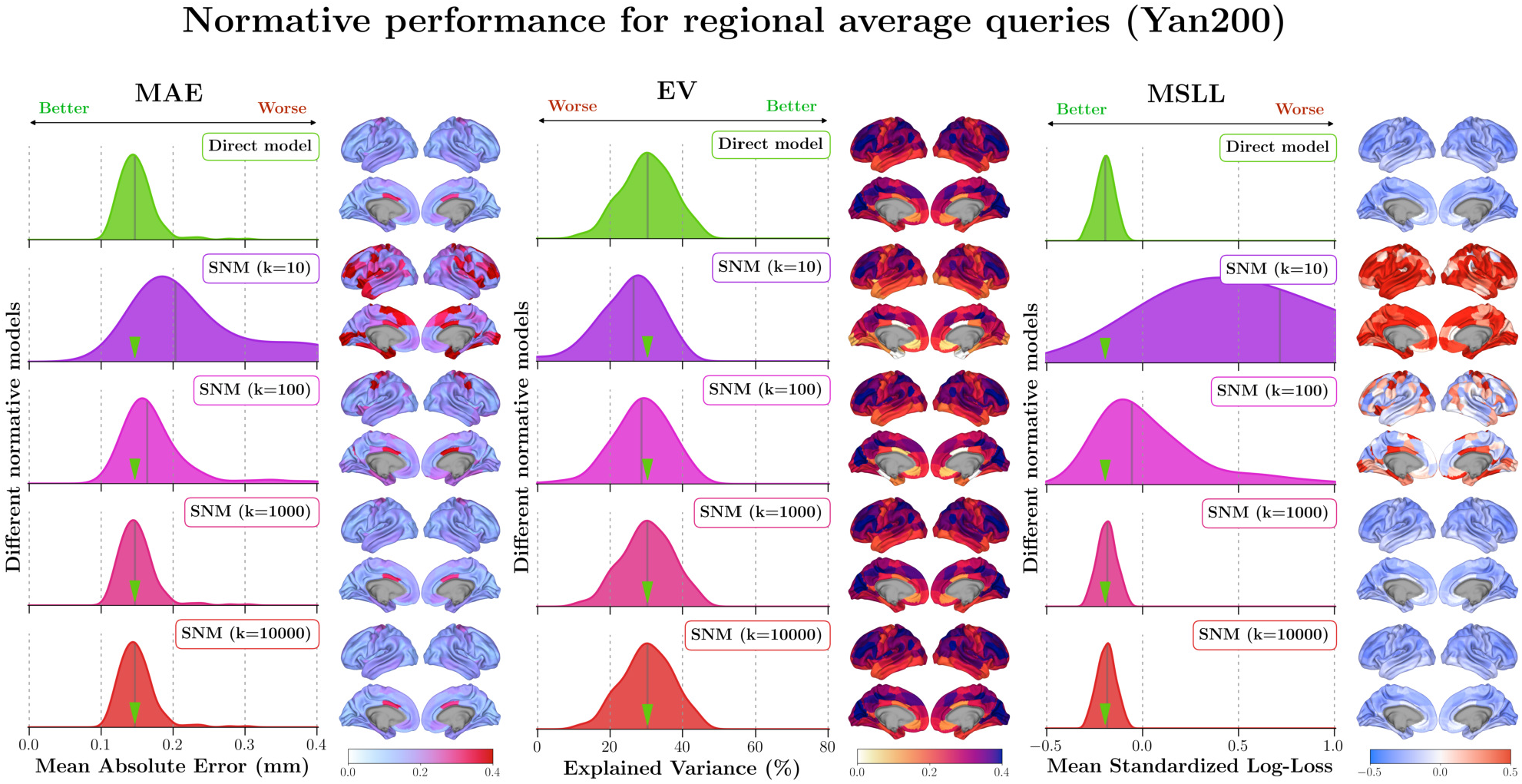
Comparison of Performance for Different Normative Models Assessing Regional Spatial Queries. Normative performance is evaluated using out-of-sample assessments of both direct and spectral normative models. The top row shows the performance of the direct model, while the following four rows present the performance of spectral models with 10, 100, 1,000, and 10,000 eigenmodes, respectively. Mean absolute error (MAE), explained variance (EV), and mean standardized log-loss (MSLL) are used to quantify model performance. Violin plots display the distribution of performance measures across 200 different spatial queries, representing regional average thickness based on the Yan200 atlas. Median values are marked by gray lines on the violin plots. A green arrow marks the median performance of the direct model over spectral performance distributions for visual comparison. Cortical surface projections show normative performance values for each region, with shared colorbars across all five rows to aid visual comparison.

##### S.1.7.5 High-Resolution Normative Estimates

Finally, Fig. S.9 assesses SNM’s performance in estimating high-resolution normative charts for spatial queries centered on various cortical surface vertices. Given the computational infeasibility of applying the direct model to every vertex, we evaluated a randomly selected subset of 4,000 vertices across the cortical surface. At this level of spatial specificity, spectral normative models with 10 and 100 modes fail to estimate normative ranges effectively compared to the direct model. However, the spectral model with 1000 modes achieves performance comparable to the direct model. Further inclusion of 10,000 modes yields marginal performance improvements. We also evaluated the sensitivity of these results to spatial granularity and asymmetry in normative queries (see Supplementary Figs. S.12, S.16).

**Supplementary Fig. S.9.**
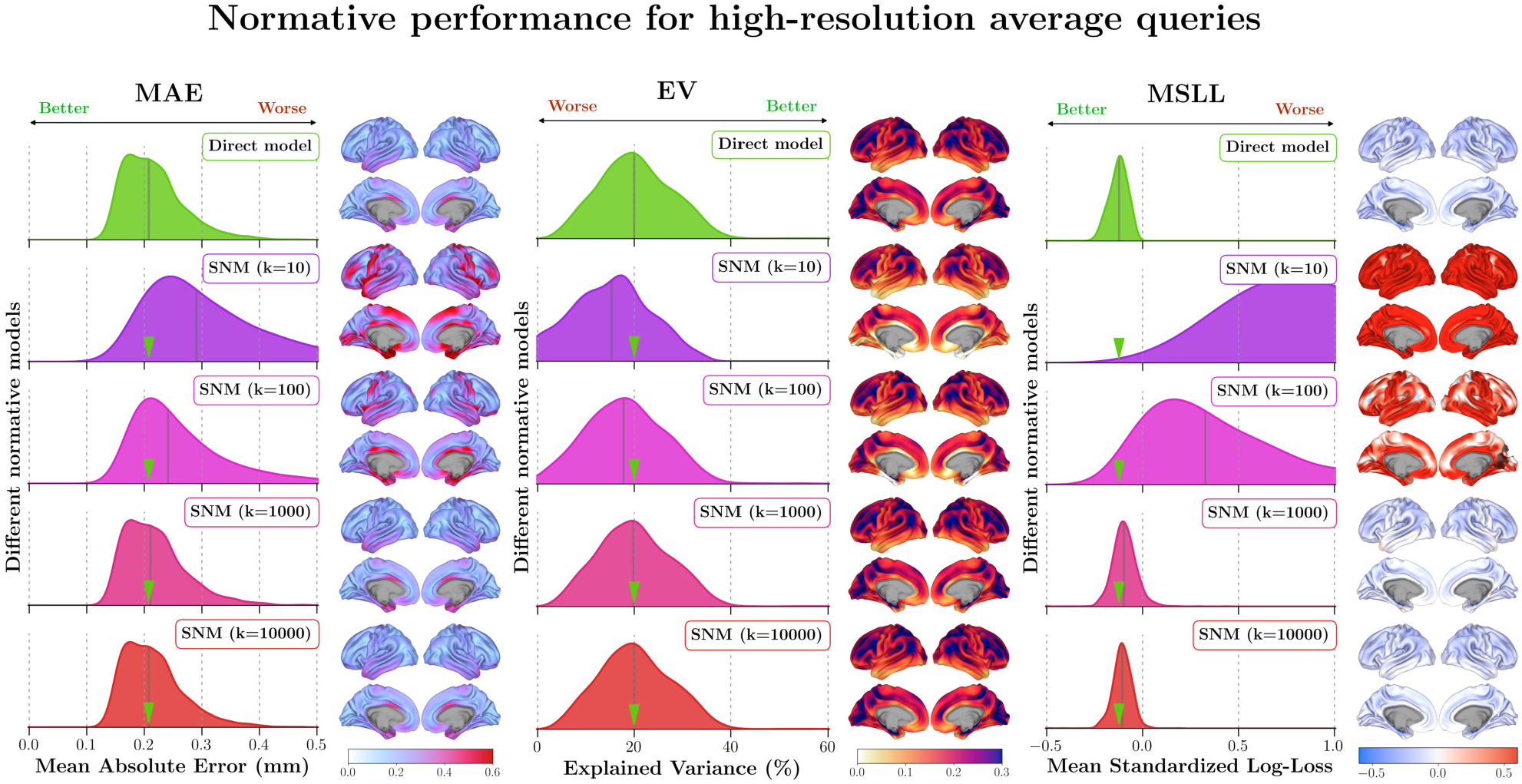
Comparison of Performance for Different Normative Models Assessing High-Resolution Spatial Queries. Normative performance is evaluated through out-of-sample assessments of both direct and spectral normative models. The top row displays the performance of the direct model, while the subsequent four rows show the performance of spectral models with 10, 100, 1,000, and 10,000 eigenmodes, respectively. Mean absolute error (MAE), explained variance (EV), and mean standardized log-loss (MSLL) are used to quantify model performance. Violin plots illustrate the distribution of performance measures across 4,000 different spatial queries, each representing an 8mm FWHM Gaussian kernel centered at a high-resolution cortical vertex (see Section S.1.6 Benchmarking High-Resolution Spatial Queries). Median values are indicated by gray lines on the violin plots. A green arrow highlights the median performance of the direct model in comparison to SNMs. Cortical surface projections show normative performance values for each vertex, depicted within its associated Yan400 region, with shared colorbars across all five rows to facilitate visual comparison.

#### S.1.8 Sensitivity Analyses for Query Characteristics

Several sensitivity analyses were conducted to evaluate the robustness of our findings to various spatial query properties. These analyses can be categorized into two main groups: those assessing the robustness of results against the granularity of the spatial queries, and those evaluating the impact of asymmetry in the spatial queries. These analyses will be respectively detailed in the ensuing sections.

##### S.1.8.1 Effect of Spatial Granularity

To evaluate the sensitivity of our findings to the spatial granularity of normative queries, we repeat the evaluations for both regional and high-resolution queries with altered parameters. For regional queries, we assess the impact of increased spatial granularity by using the Yan400 parcellation template, which contains twice as many brain regions as the Yan200 atlas, thereby doubling the spatial granularity of the regional signals. Similarly, the spatial granularity of the high-resolution signals is altered by adjusting the spatial smoothing kernel strength. Specifically, high-resolution evaluations are repeated using a 4mm FWHM Gaussian smoothing kernel, which effectively doubles the spatial granularity of the high-resolution signals.

Supplementary Fig. S.10 provides a comparative view of reconstruction accuracy performance as affected by signal granularity. As anticipated, increasing spatial granularity introduces higher spatial frequencies that are captured by higher graph frequencies. This is evident from the slight shift in proportional energy plots, which indicates an increased contribution of higher frequency modes to signal approximation. Additionally, the results show that the signal reconstruction accuracy for both regional and high-resolution spatial queries reaches a plateau after including the first 1,000 and 2,000 eigenmodes, respectively. Finally, the relatively higher values of SMSE for high-resolution signals with a 4mm FWHM smoothing kernel suggest that high-resolution spectral evaluations are better suited for signals smoothed to 8mm FWHM.

Supplementary Fig. S.11 illustrates the impact of regional signal granularity on the goodness of fit of normative models. Repeating the evaluations shown in Fig. S.8, we demonstrate that, irrespective of the regional signal granularity, the inclusion of 1,000 eigenmodes in the spectral model achieves performance comparable to that of a direct model. Similarly, Supplementary Fig. S.12 repeats the evaluations presented in Fig. S.9 and shows that a spectral model with 1,000 modes yields normative estimates comparable to those produced by the direct model. While these results indicate the robustness of the spectral normative framework to changes in spatial granularity, any query used with the spectral model should nevertheless be tested to assess adherence to the low-pass spectral regime (e.g., by evaluating reconstruction accuracy as quantified by metrics such as SMSE).

**Supplementary Fig. S.10.**
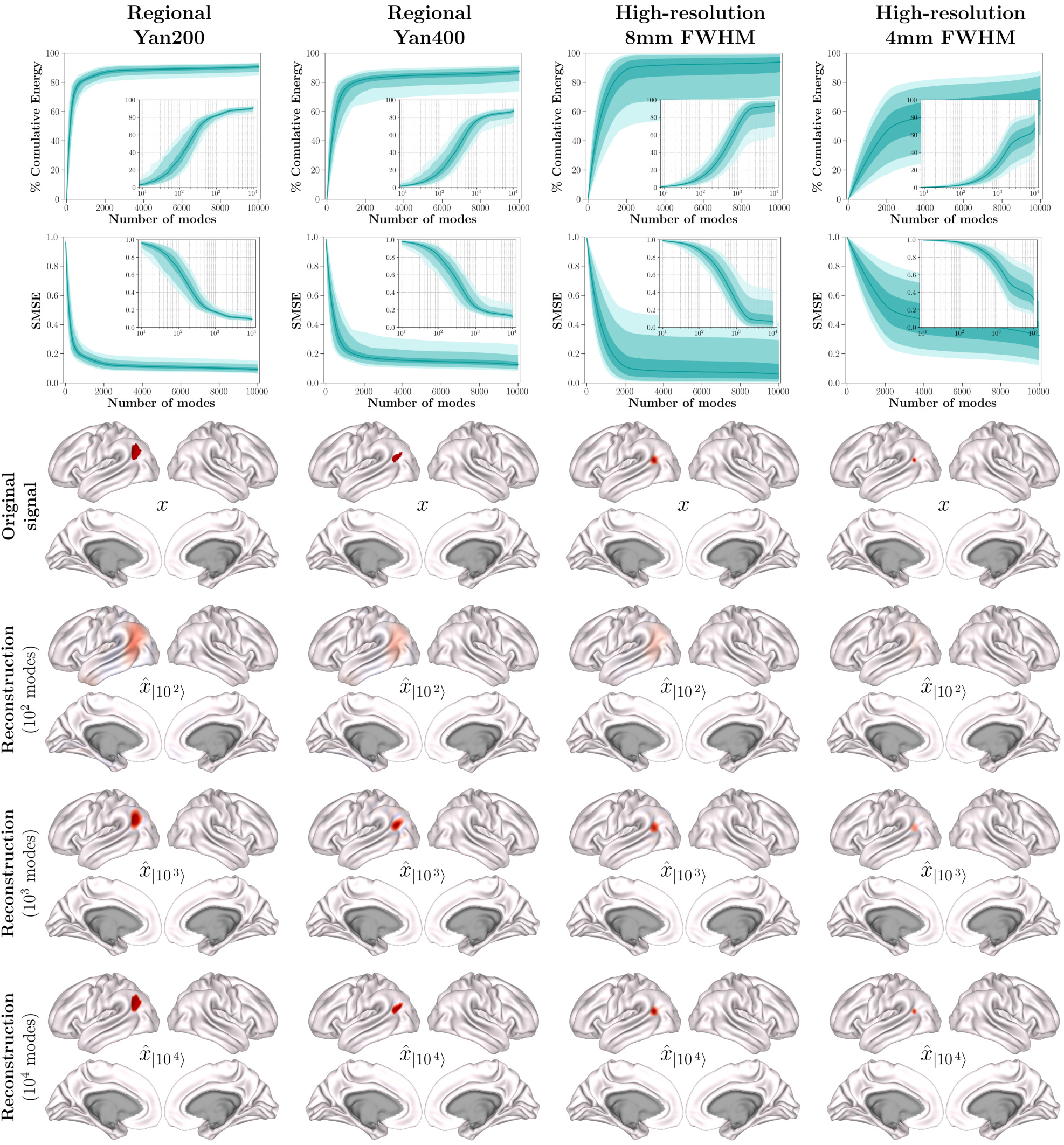
Signal Reconstruction Accuracies across Different Granularity Scales. The evaluations presented in Extended Data Fig. E.1 are repeated to examine the effect of doubling the spatial granularity of the brain signals. The columns (from left to right) display spectral reconstruction accuracies for regional signals using the Yan200 and Yan400 atlases, as well as for high-resolution signals defined by 8mm FWHM and 4mm FWHM smoothing kernels. In the shaded line plots, the lines represent the median across all observations, while the shades indicate the [25, 75], [5, 95], and [1, 99] percentiles. The first row shows the proportional energy independently contributed by each eigenmode (logarithmic x-axis). The second row presents the standardized mean square error (SMSE) as a function of the number of low-frequency eigenmodes used for reconstruction (logarithmic x-axis for the insets). The third row illustrates one exemplary brain signal from each category, while the last three rows show the same signal reconstructed using 100, 1,000, and 10,000 eigenmodes, respectively.

**Supplementary Fig. S.11.**
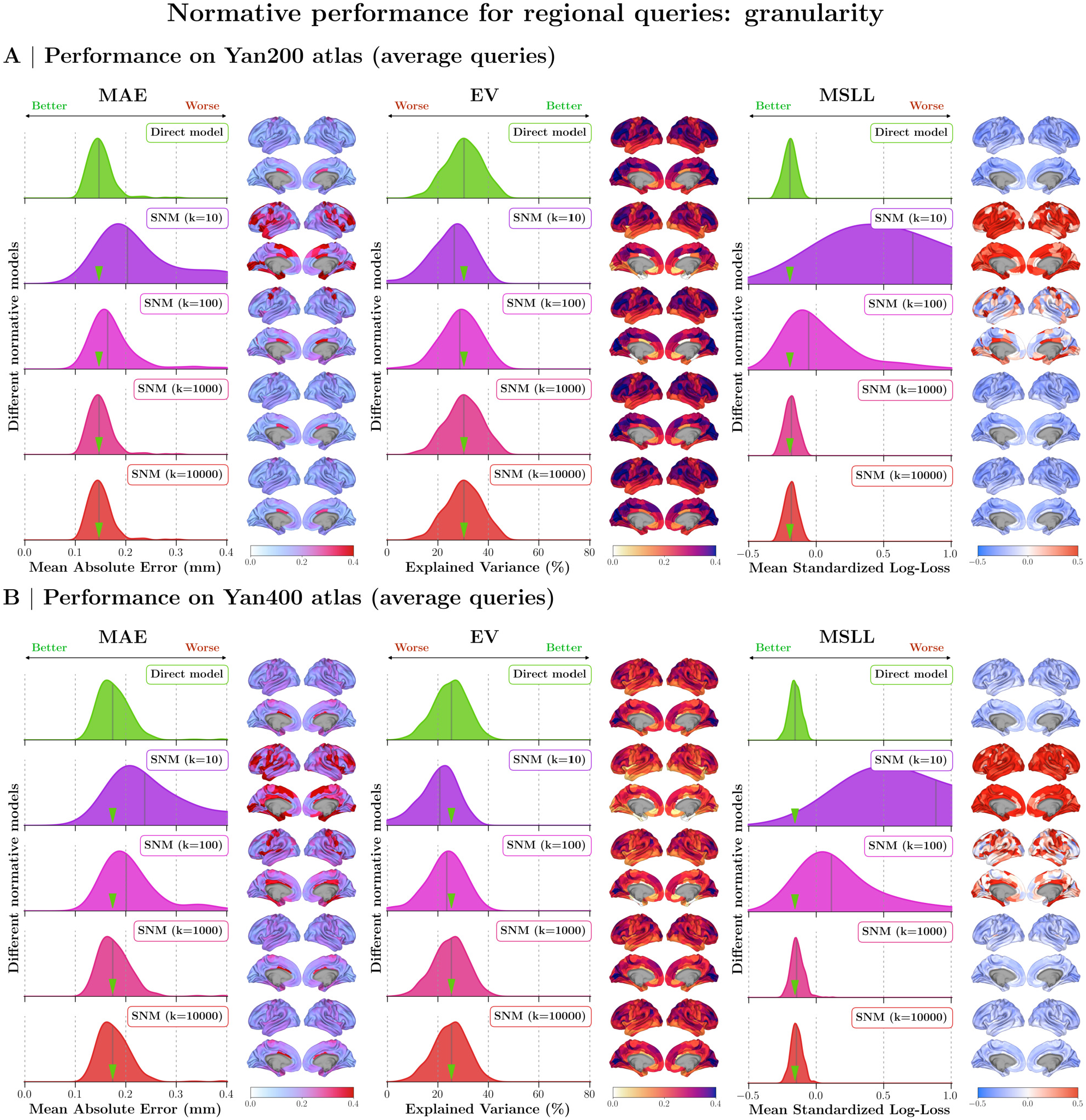
Sensitivity of Normative Models Assessing Regional Spatial Queries at Different Granularity Scales. Normative performance is evaluated using out-of-sample assessments of both direct and spectral normative models. **(A)** same as Fig. S.8, repeated to aid visual comparison. **(B)** Same evaluations as panel A, but on regional queries from Yan400 atlas (increased spatial granularity).

**Supplementary Fig. S.12.**
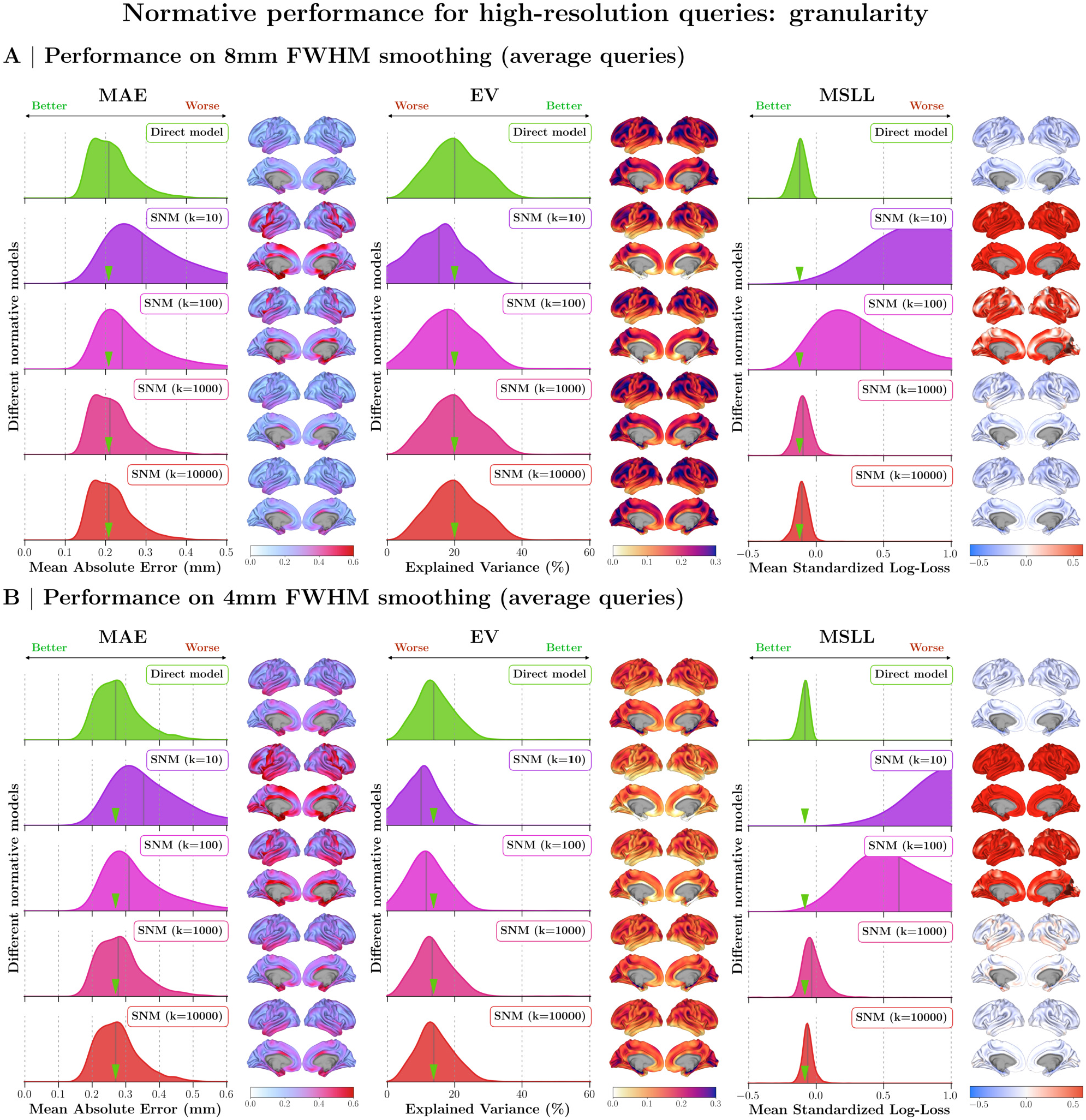
Sensitivity of Normative Models Assessing High-Resolution Spatial Queries at Different Granularity Scales. Normative performance is evaluated using out-of-sample assessments of both direct and spectral normative models. **(A)** same as Fig. S.9, repeated to aid visual comparison. High-resolution spatial queries are generated using an 8mm FWHM Gaussian smoothing kernel. **(B)** Same evaluations as panel A, but on high-resolution queries generated using a 4mm FWHM Gaussian smoothing kernel (increased spatial granularity).

##### S.1.8.2 Effect of Spatial Asymmetry

A notable improvement of the spectral normative framework is its spatial versatility. This enables spectral models to extend beyond conventional average-based normative queries and gives them the ability to model a wider range of possible queries. As long as the query of interest adheres to the lowpass spectral regime, it can include various spatial patterns of interest. Asymmetric spatial queries are an interesting example of such patterns. Specifically, spectral normative models can be used to infer abnormal deviations in thickness lateralization by constructing a query that assesses the difference between the average thickness of a region/locus on the left hemisphere and its contralateral counterpart on the right hemisphere. This added benefit requires no extra training time and works out of the box. Hence, we evaluate the sensitivity of our main findings to the spatial asymmetry of normative queries.

We repeat the evaluations for brain-wide, regional, and high-resolution queries after introducing lateralization asymmetry. For brain-wide queries, we use a query that computes the average thickness of the query on the left cortex and subtracts it from the average of the part of the query that lies on the right cortex. For regional queries, given that the Yan atlas provides a homotopic parcellation of the cerebral cortex (i.e., parcels on the left and right cortices are paired), we evaluate 100 spatial normative queries, each assessing the difference in the average thickness of one homotopic parcel pair from Yan200 parcellation. Finally, for high-resolution queries, given that the fs-LR template space is aligned across the left and right cortices, we select pairs of homotopic vertices and construct spatial queries that compare their average thicknesses as described by 8mm FWHM smoothing kernels centered on respective vertex pairs.

**Supplementary Fig. S.13.**
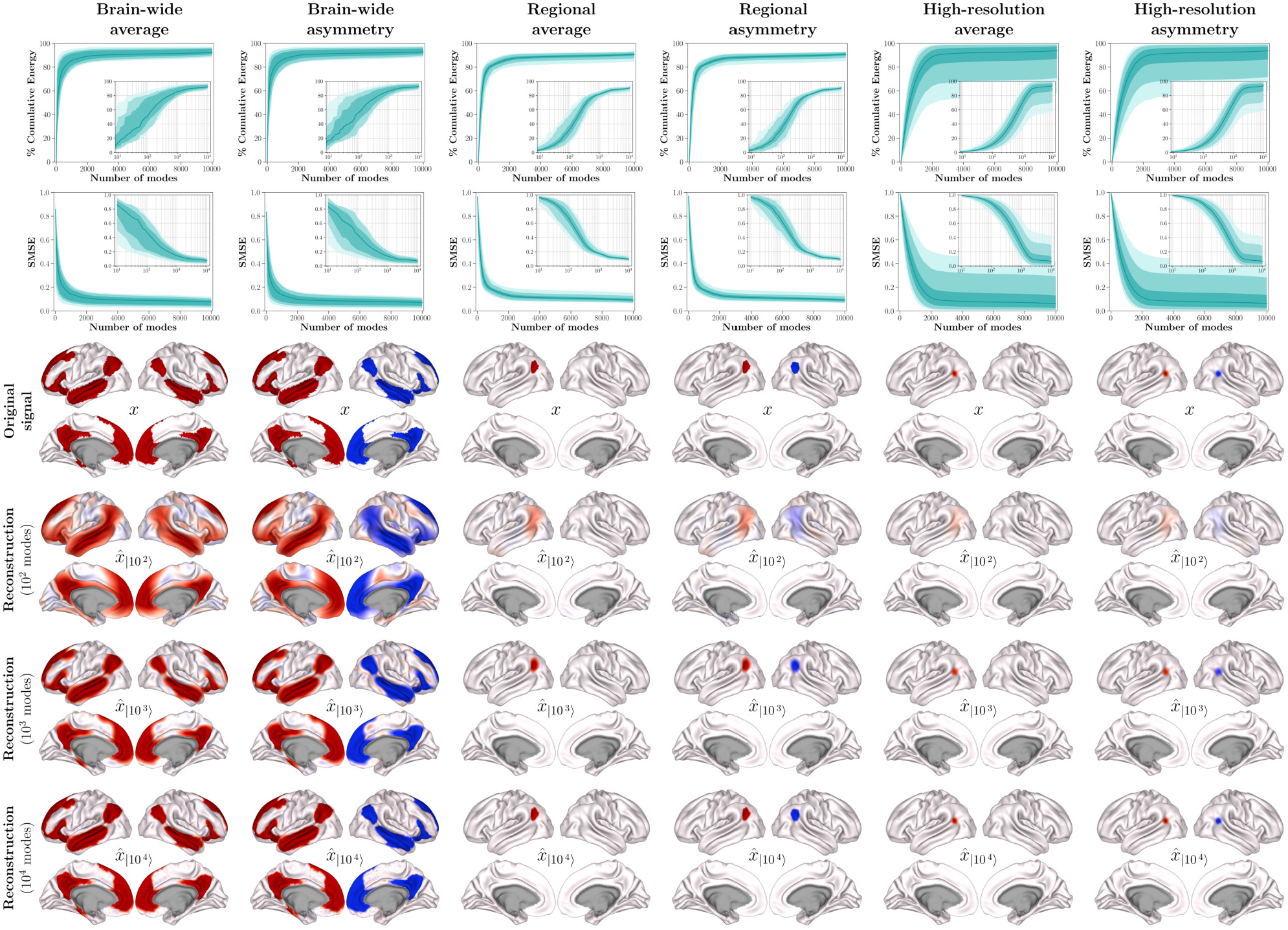
Signal Reconstruction Accuracies for Average vs. Asymmetric Signals. The evaluations presented in Extended Data Fig. E.1 are repeated to examine the effect of introducing asymmetry to the brain signals. The columns display spectral reconstruction accuracies for pairs of (average, vs. lateralized/asymmetric) brain-wide, regional, and high-resolution signals. In the shaded line plots, the lines represent the median across all observations, while the shades indicate the [25, 75], [5, 95], and [1, 99] percentiles. The first row shows the proportional energy independently contributed by each eigenmode (logarithmic x-axis). The second row presents the standardized mean square error (SMSE) as a function of the number of low-frequency eigenmodes used for reconstruction (logarithmic x-axis for the insets). The third row illustrates one exemplary brain signal from each category, while the last three rows show the same signal reconstructed using 100, 1,000, and 10,000 eigenmodes, respectively.

Supplementary Fig. S.13 provides a comparative view of reconstruction accuracy performance as affected by signal asymmetry. Results indicate that signal asymmetry has negligible impact on eigen-mode energy proportions and reconstruction accuracy (quantified by SMSE). Comparing asymmetric queries to average queries (presented in Extended Data Fig. E.1), we observe that asymmetric signals require a comparable number of modes for accurate reconstruction as average signals within each spatial query family (brain-wide, regional, and high-resolution). Regardless of signal symmetry, SMSE for the reconstruction of brain-wide, regional, and high-resolution signals respectively reaches below 0.2 after the inclusion of the first 400, 600, and 1,000 eigenmodes. Thus, the reported findings on the number of eigenmodes required to reconstruct brain signals remain consistent regardless of signal asymmetry.

Supplementary Figs. S.14, S.15, and S.16 illustrate the impact of signal asymmetry on the goodness of fit of the associated normative models. Repeating the evaluations shown in Figs. S.7, S.8, and S.9 shows that irrespective of the normative modeling framework (direct vs. spectral), goodness of fit, particularly when assessed by MSLL, is lower for lateralized norms. This effect is more pronounced at higher resolutions, suggesting that healthy norms of thickness lateralization are better studied at lower spatial specificity (functional networks or parcels). As the goodness of fit in predicting the central tendency (quantified via MAE) is less affected, we speculate that this reduction in the accuracy of lateralized queries is due to the misaligned gyrification patterns between contralaterally aligned high-resolution vertices, rendering lateralization evaluations less meaningful at higher resolutions.

**Supplementary Fig. S.14.**
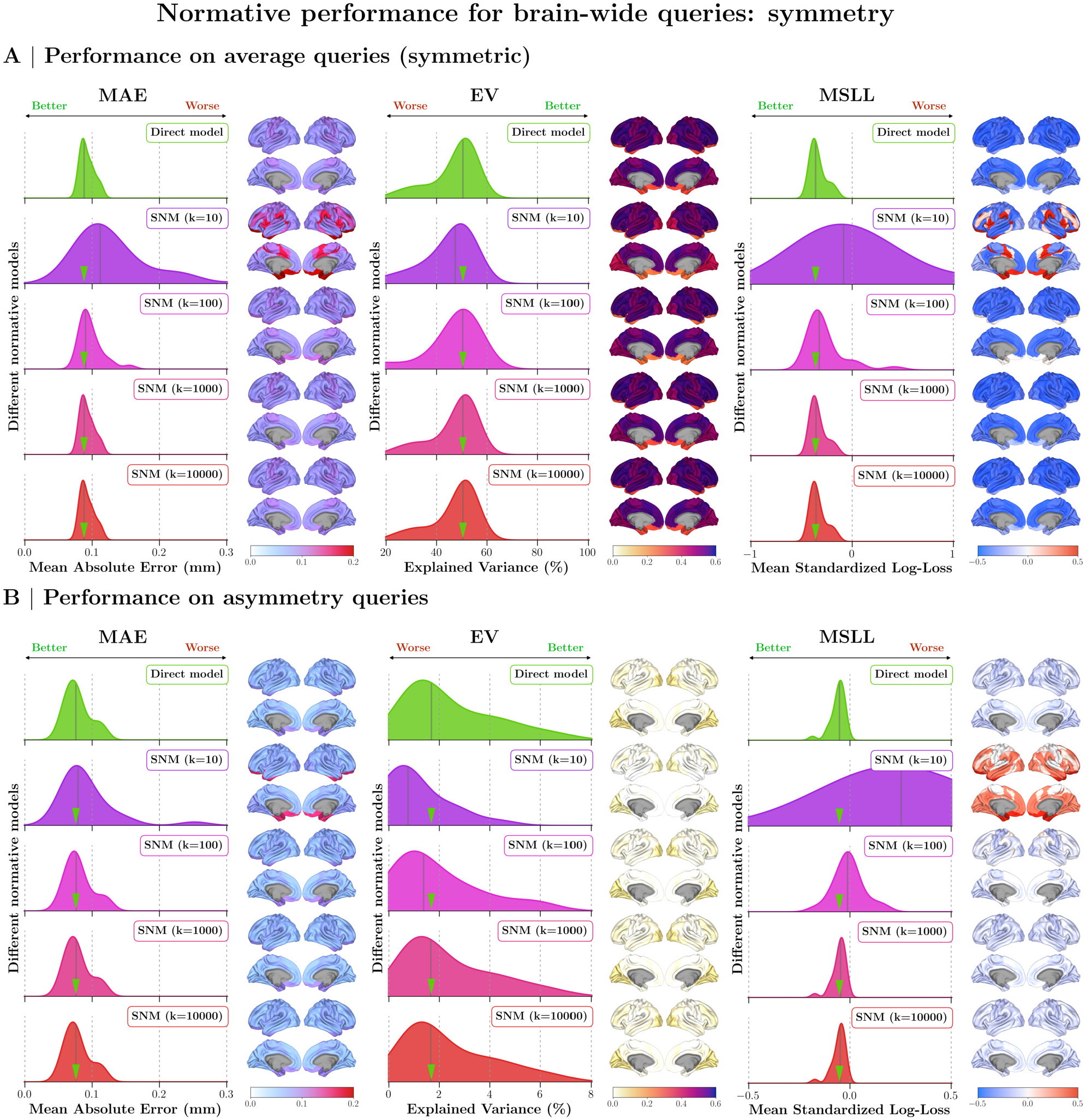
Sensitivity of Normative Models to Assessing Lateralized Brain-Wide Spatial Queries. Normative performance is evaluated using out-of-sample assessments of both direct and spectral normative models. **(A)** same as Fig. S.7, repeated to aid visual comparison. **(B)** Same evaluations as panel A, but on lateralized counterparts of the brain-wide queries.

Despite this effect, which was not specific to the spectral model, we observe that, similar to the main findings, the inclusion of 100 eigenmodes is sufficient to model brain-wide normative thickness ranges with the spectral model, achieving performance on par with the direct model. As with the main findings, the number of modes required to achieve comparable performance to that of a direct model at the resolution of regional or vertex-wise queries increases to 1,000 eigenmodes. These sensitivity evaluations demonstrate that our findings are robustly replicable for signals with asymmetry, and that the inclusion of 1,000 eigenmodes can provide comparable performance to that of a conventional model across a wide range of spatial queries.

**Supplementary Fig. S.15.**
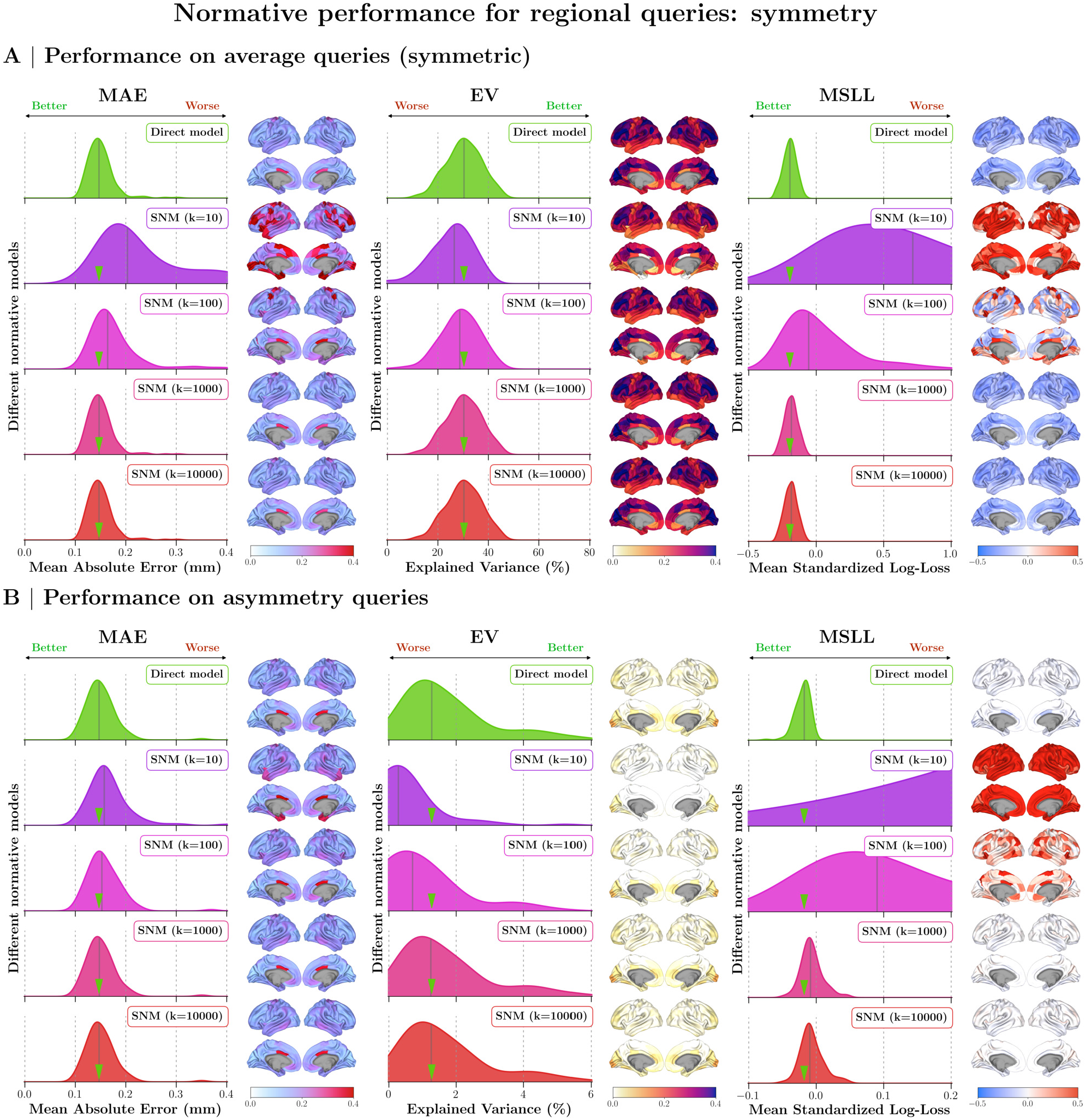
Sensitivity of Normative Models to Assessing Lateralized Regional Spatial Queries. Normative performance is evaluated using out-of-sample assessments of both direct and spectral normative models. **(A)** same as Fig. S.8, repeated to aid visual comparison. **(B)** Same evaluations as panel A, but on lateralized counterparts of the regional queries.

**Supplementary Fig. S.16.**
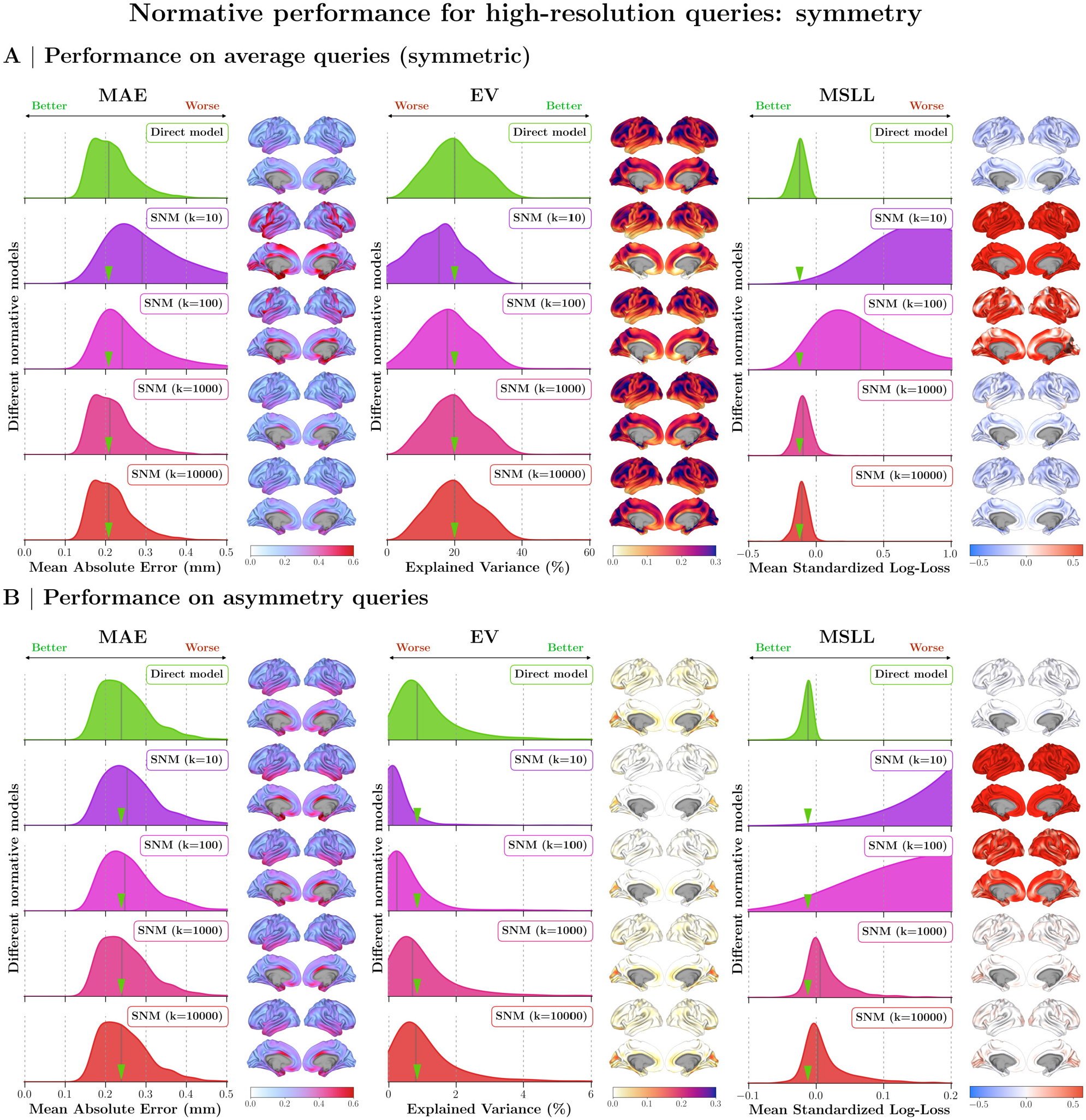
Sensitivity of Normative Models to Assessing Lateralized High-Resolution Spatial Queries. Normative performance is evaluated using out-of-sample assessments of both direct and spectral normative models. **(A)** same as Fig. S.9, repeated to aid visual comparison. **(B)** Same evaluations as panel A, but on lateralized counterparts of the high-resolution queries.

#### S.1.9 Extended Spatial Correspondence Analysis of Cortical Maps

The pairwise comparisons in Fig. 5B were restricted to a curated set of maps representing established cortical hierarchies. To extend this analysis, we compared each thickness growth gradient with a broader library of 81 cortical maps, comprising the maps included in the main analysis together with published maps predominantly sourced from the neuromaps repository^58^. These included neurotransmitter receptor and transporter densities, metabolic and perfusion maps, oscillatory power across canonical frequency bands, and meta-analytic functional maps. All maps were resampled to a common 360-parcel parcellation^59^ and tested as described in Section 4.11 Testing Spatial Correspondence of Cortical Maps, with FDR correction across all 243 tests. Of the 243 associations tested, 60 survived FDR correction at *p*_*FDR*_ *<* 0.05, comprising 27 for TGG1, 3 for TGG2, and 30 for TGG3 (Supplementary Fig. S.17). The relatively few significant associations for TGG2 suggest that the late-life thinning axis is less well represented among the molecular and functional maps in this library than are TGG1 and TGG3.

Grouping significant associations by map type revealed distinct profiles across the three gradients. TGG1 showed consistently positive associations with neuromodulatory receptor and transporter maps spanning serotonergic^60–62^ (5-HT_1A_, `cumi101` and `way100635`, *r* = 0.69 and 0.61; 5-HTT, `dasb` and `madam`, 0.34–0.50), dopaminergic^63–67^ (D_2_, `flb457` and `fallypride`, 0.54–0.62; DAT, `fpcit`, 0.41; D_1_, `sch23390`, 0.39), opioidergic^68–70^ (*µ*-opioid, `carfentanil`, 0.54–0.59; *κ*-opioid, `ly2795050`, 0.43), cannabinoid^71^ (CB_1_, `omar`, 0.52), cholinergic^72^ (VAChT, `feobv`, 0.46) and histaminergic^73^ (H_3_, `gsk189254`, 0.42) systems. TGG1 also showed positive correspondence with high gamma-band power^30,74^ (`meggamma2`, *r* = 0.46) and negative correspondence with intracortical myelin^59^ (`myelinmap`, *r* = −0.54) and oxidative and glucose metabolism^75^ (`cmr02` and `cmrglc`, *r* = −0.50 and −0.47). Significant associations for TGG2 comprised aging-related thickness decline, beta-band power^30,74^ (`megbeta`, *r* = −0.43), and the principal component of meta-analytic cognitive term maps^76–78^ (`cogpc1`, *r* = −0.36). TGG3 showed the strongest correspondence with molecular maps of synaptic and neurotransmitter organization, including metabotropic glutamate receptor density replicated across three independent tracer studies^79–81^ (mGluR_5_, `abp688`, *r* = 0.51–0.53), GABA_A_ receptor density^66,82^ (`flumazenil`, *r* = 0.32–0.40), synaptic vesicle glycoprotein 2A density^83^ (`ucbj`, *r* = 0.30), and several postsynaptic serotonin receptor subtypes^60,61,84,85^ (5-HT_2A_, `cimbi36`, *r* = 0.44; 5-HT_1B_, `az10419369` and p943, *r* = 0.33–0.37; 5-HT_4_, `sb207145`, *r* = 0.34; 5-HT_6_, `gsk215083`, *r* = 0.33). TGG3 was additionally associated with cerebral perfusion and glucose metabolism^75,86^ (`meancbf`, `cbf` and `cmrglc`, *r* = 0.34–0.50), including a developmental-cohort perfusion map (*r* = 0.50), with theta- and delta-band power^30,74^ (`megtheta` and `megdelta`, *r* = 0.35–0.43), and negatively with intracortical myelin^59^ (`myelinmap`, *r* = −0.31). Interpretation of these correspondences is provided in the Discussion.

**Supplementary Fig. S.17.**
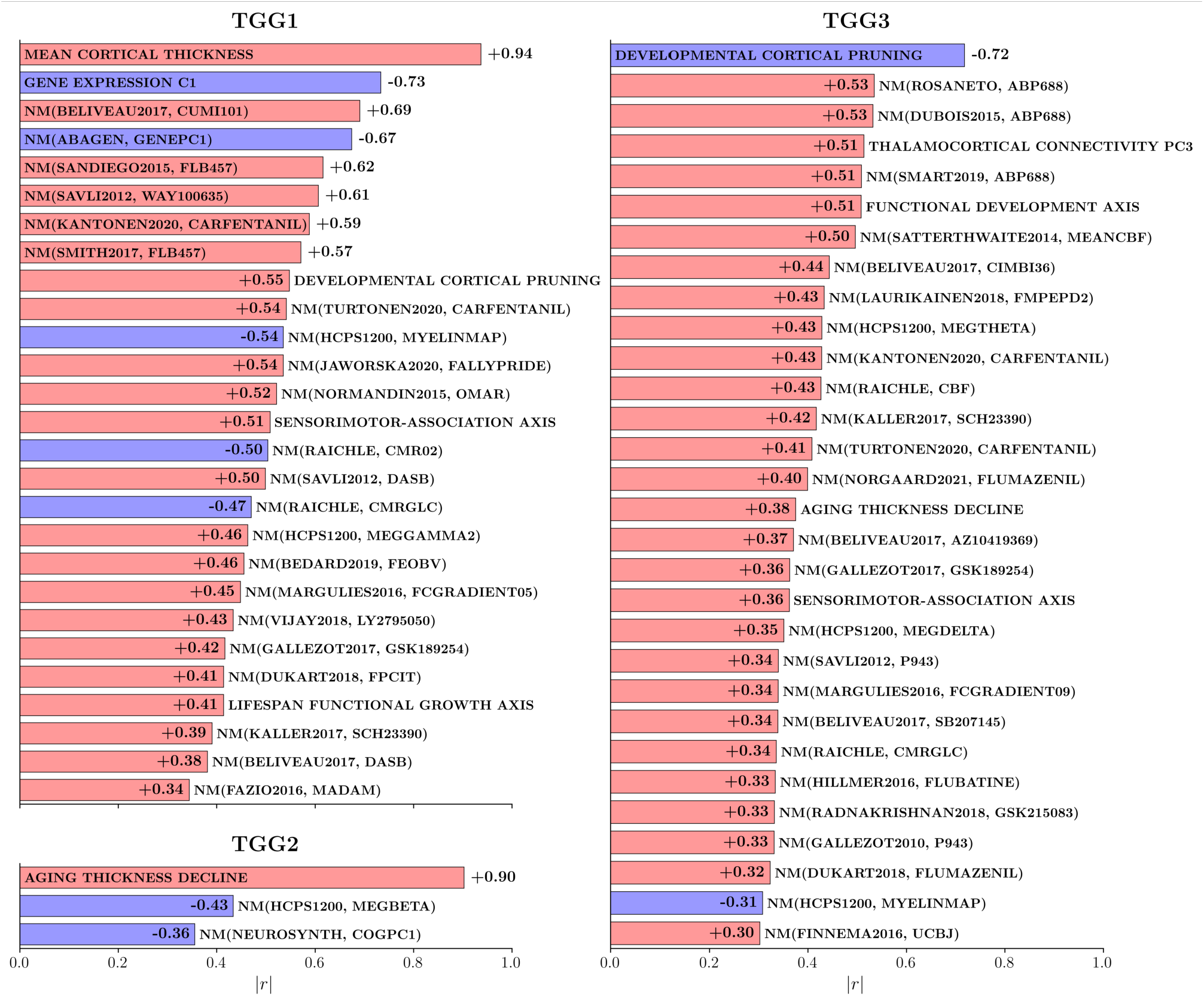
Extended Spatial Correspondence Between Thickness Growth Gradients and Other Cortical Maps. Each of TGG1–3 was compared with a library of 81 cortical maps at parcel resolution (81 × 3 = 243 tests; see Section 4.11 Testing Spatial Correspondence of Cortical Maps). Panels show all associations surviving FDR correction across the 243 tests, ranked by |*r*|. Bar length denotes |*r*|, and bar color indicates the sign of the association (red, positive; blue, negative). Labels prefixed with NM denote maps included only in this extended analysis and follow neuromaps naming conventions, with the source and description keyword indicated^58^. Other labels denote maps also included in the comparison in Fig. 5B and correspond to the maps abbreviated there.

#### S.1.10 Within-Group Cognitive Associations

To assess the significance of cognitive associations within each cohort, we repeated the analyses from Section 2.5 SNM Recovers Established AD Signatures and Quantifies Individual Atrophy Burden on the HC, MCI, and AD subsets, with the expectation of no association in HC, weaker associations in MCI, and stronger effects in AD. For healthy controls (HC), high-resolution cortical associations did not reach significance in any region after FDR correction (Supplementary Fig. S.18), and ETVC showed no association with cognitive performance (*r* = −0.02, *p* = 0.90), irrespective of the z-threshold. In the MCI cohort, ETVC showed a relatively weak but significant association with cognitive impairment (*r* = −0.20, *p* = 0.004; Supplementary Fig. S.19), while regional analyses identified atrophy in the entorhinal cortex to be associated with MCI cognitive impairment. In contrast, the AD cohort displayed widespread neocortical atrophy patterns significantly linked to cognitive impairment (Supplementary Fig. S.20), with reductions in the temporal pole, superior and middle temporal gyri, entorhinal cortex, parahippocampal cortex, precuneus, supramarginal gyrus, and superior, middle, and inferior frontal gyri. Additionally, ETVC showed a stronger association with cognitive impairment in the AD cohort (*r* = −0.34, *p <* 0.0001). Interestingly, this predictive capacity diminished when the z-score threshold was raised to zero or any positive value, with optimal predictions achieved between z-score thresholds of −2 and −3. Together, these findings suggest that the derived normative atrophy marker of cognitive impairment exhibits increasing sensitivity from MCI to AD and is most informative in the presence of extreme atrophy.

**Supplementary Fig. S.18.**
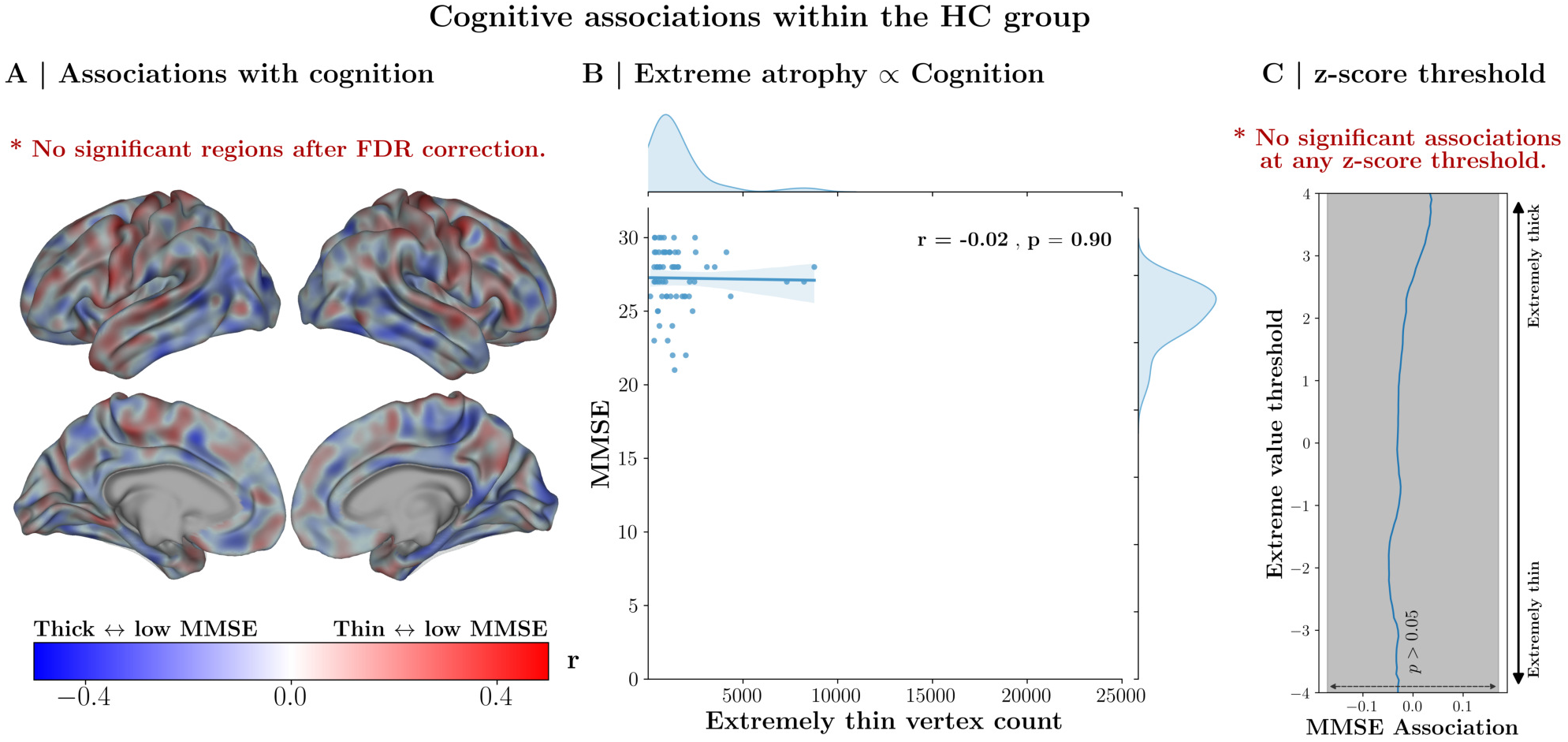
Cognitive Association Tests within the Healthy Cohort (HC). **(A)** Vertex-level normative assessments were tested for linear associations with cognitive performance (MMSE), but no vertices reached significance after nonparametric FDR correction. **(B)** Extreme value statistics for severe atrophy (ETVC, z-score threshold of −1.96) were assessed for their predictive capability on cognitive performance, revealing no sensitivity of ETVC to cognitive variations in HC. **(C)** A range of alternative z-score thresholds was tested, demonstrating that ETVC does not predict cognitive performance in HC, regardless of threshold choice.

**Supplementary Fig. S.19.**
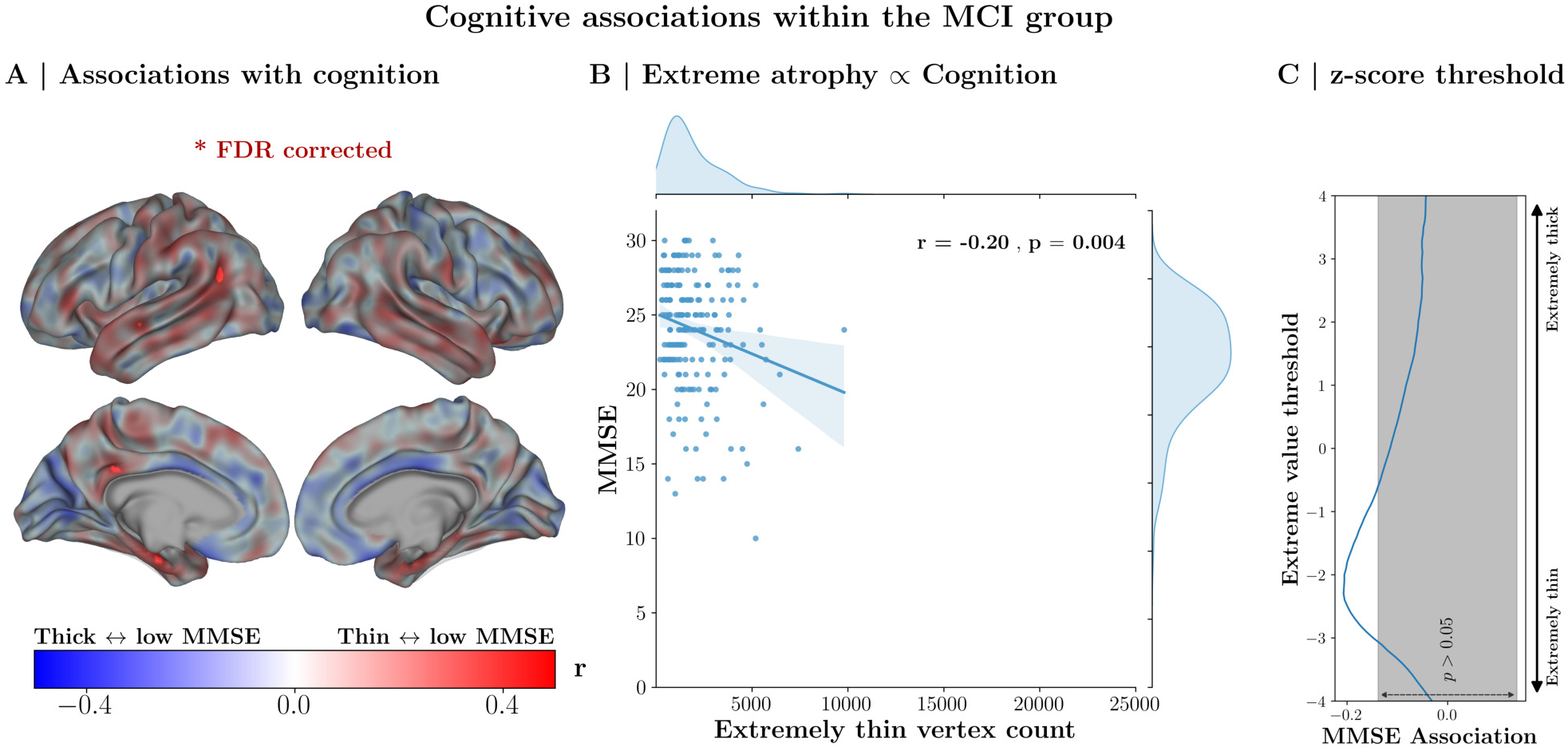
Cognitive Association Tests within the MCI Cohort. **(A)** Vertex-level normative assessments were tested for linear associations with cognitive performance (MMSE) within the MCI cohort. A limited set of vertices reached statistical significance, indicating that extreme atrophy in specific regions may serve as a marker of cognitive impairment. **(B)** Extreme value statistics for severe atrophy (ETVC, z-score threshold of −1.96) were evaluated for their predictive capability on cognitive performance, showing a modest association between ETVC and cognitive variations within MCI (*r* = −0.20). **(C)** Testing a range of alternative z-score thresholds shows that extreme focal atrophy (−3 *< z <* −2) can predict cognitive performance in MCI.

**Supplementary Fig. S.20.**
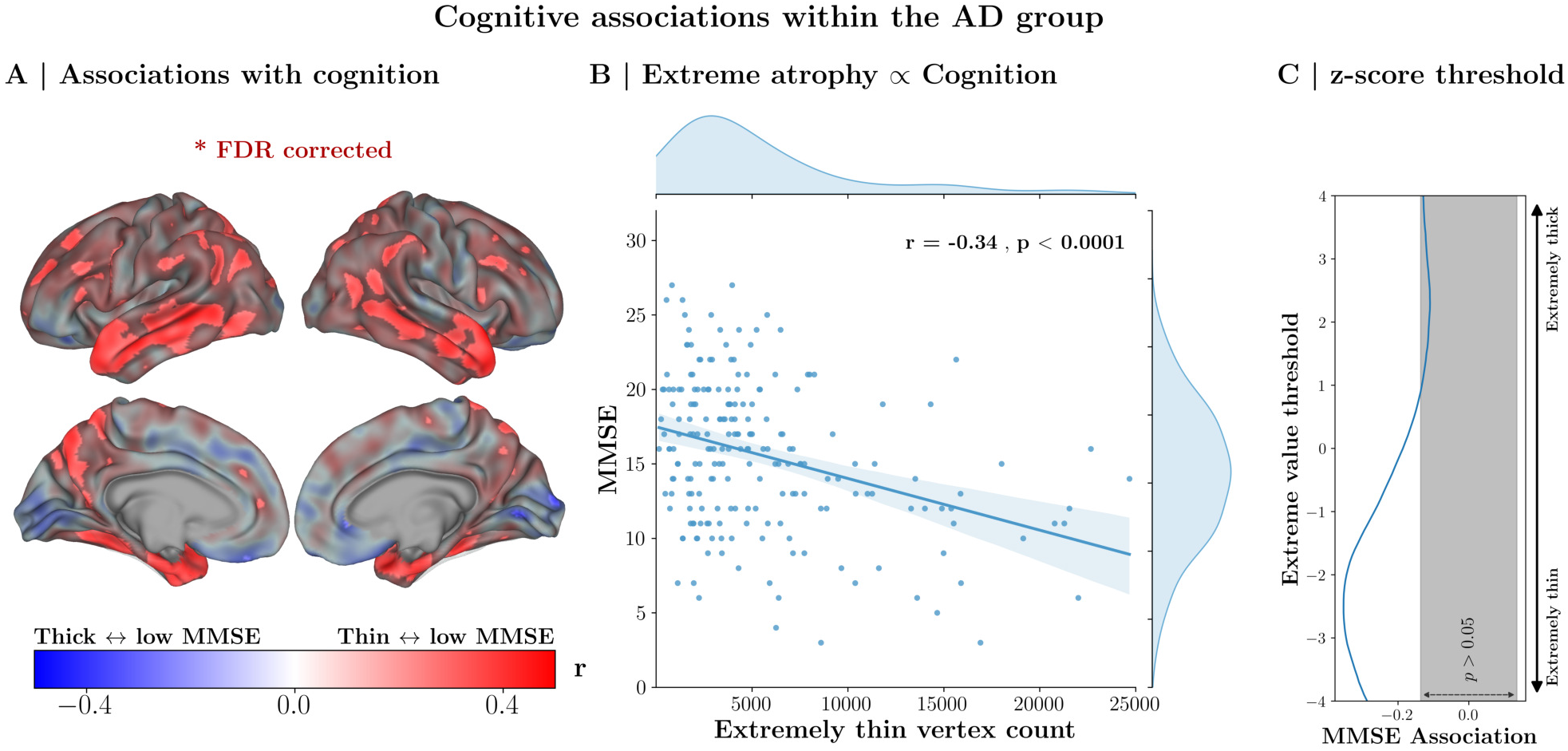
Cognitive Association Tests within the AD Cohort. **(A)** Vertex-level normative assessments were tested for linear associations with cognitive performance (MMSE) within the AD cohort. A widespread set of cortical regions reached statistical significance, highlighting extensive brain-wide associations as markers of AD-related cognitive impairment. **(B)** Extreme value statistics for severe atrophy (ETVC, z-score threshold of −1.96) were evaluated for predictive capability regarding cognitive performance, revealing significant predictive power of ETVC to capture cognitive variations within the AD cohort. **(C)** Testing across a range of alternative z-score thresholds indicated that only ETVCs with a negative z-threshold significantly predicted cognitive performance in the AD cohort. Notably, extreme thinning (z-thresholds from −2 to −3) proved to be the strongest predictor of cognitive impairment.

#### S.1.11 Brain-Wide Cognitive Associations

To highlight the benefits of inferring high-resolution spatial norms, we compare the predictive performance of the ETVC normative measure derived from high-resolution brain charting with that of a brain-wide normative z-score computed from a spatially coarse brain chart of mean cortical thickness (averaged across the entire cortex). Using the fine-tuned SNM, we calculate individual z-scores for deviations in average cortical thickness and replicate the evaluations from Figs. 6C, S.18B, S.19B, and S.20B. As shown in Fig. S.21, the results consistently indicate that while brain-wide normative measures can predict cognitive impairments, high-resolution ETVC measures outperform them. Specifically, ETVC achieves higher prediction accuracy in the entire sample (ETVC: |*r*| = 0.53, brainwide: |*r*| = 0.41) and in the AD subsample (ETVC: |*r*| = 0.34, brain-wide: |*r*| = 0.25).

**Supplementary Fig. S.21.**
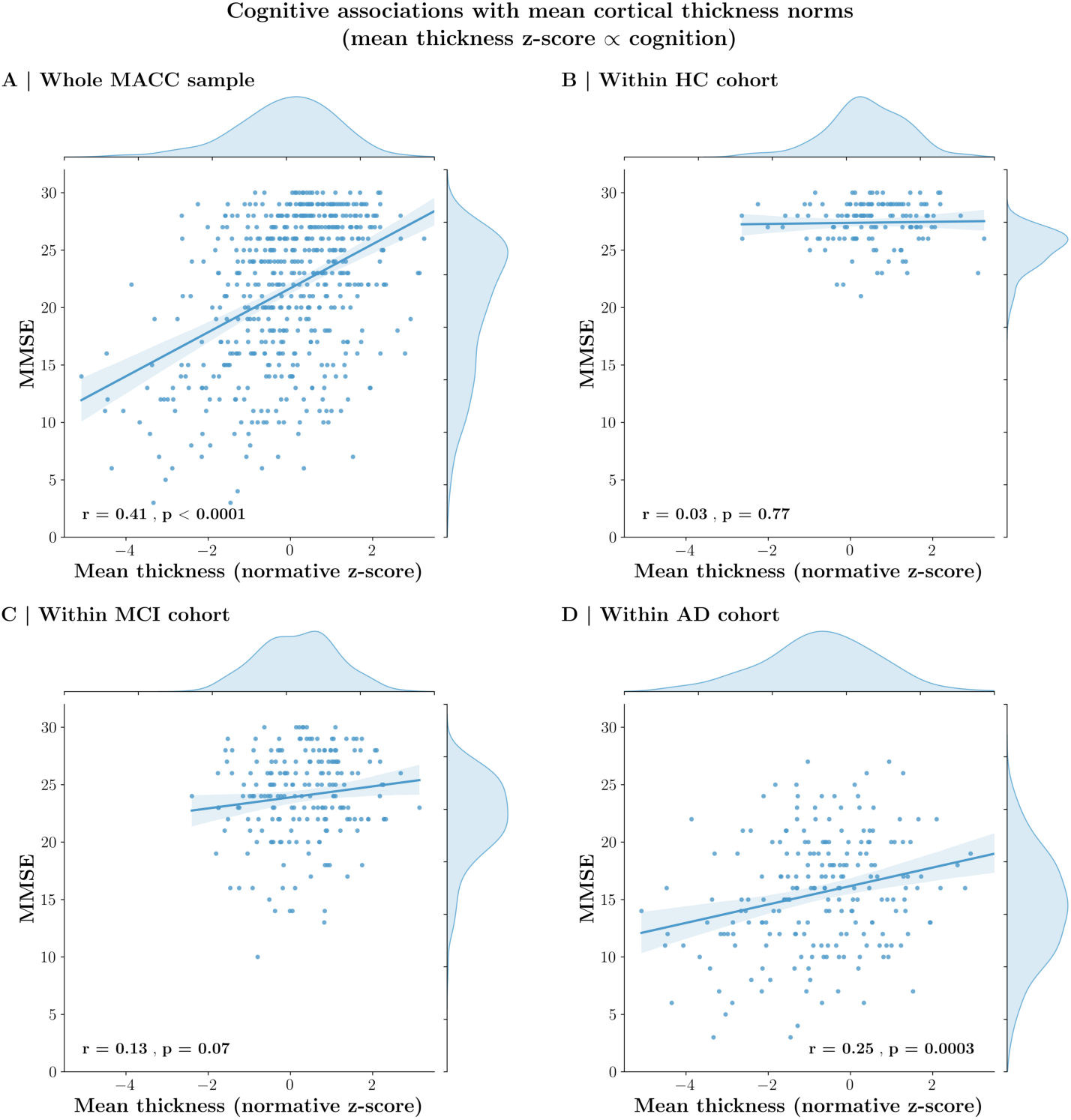
Cognitive Association Tests with a Coarse Brain-Wide Normative Metric. Normative deviations (z-scores) in mean cortical thickness were evaluated for their predictive capability regarding cognitive performance (quantified by MMSE). Predictive performance was assessed in **(A)** the entire MACC dataset, as well as within the **(B)** HC, **(C)** MCI, and **(D)** AD cohorts individually. While the global association is robust, the attenuated correlation in HC and MCI is attributed to restricted variance and a ceiling effect in MMSE scores. Sensitivity evaluations in Section S.1.12 Association Consistency Across Diagnostic Cohorts confirm that the direction of effect is preserved across groups.

#### S.1.12 Association Consistency Across Diagnostic Cohorts

The evaluation of brain-behavior associations across a heterogeneous clinical spectrum necessitates a verification that observed global trends are representative of underlying within-group relationships. Specifically, if diagnostic cohorts occupy distinct ranges of both the predictor (biomarker) and the outcome (MMSE), a pooled correlation could potentially be driven by mean differences between groups rather than a consistent biological effect. To ensure the robustness of the associations presented in Sections S.1.11 and S.1.10, we performed a series of sensitivity analyses to test for slope consistency and to control for group-level baseline shifts.

First, we implemented Linear Mixed-Effects Models (LMM) for both biomarkers (ETVC and mean thickness z-score), treating the diagnostic group as a random intercept to account for group-specific baseline shifts in MMSE (MMSE ~ Biomarker+(1|Diagnosis)). As detailed in Table S.3 and Table S.4, the fixed effect of the biomarker remained highly significant for both ETVC (*z* = −6.53, *p <* 0.001) and mean thickness z-scores (*z* = 4.28, *p <* 0.001). This confirms that the brain-behavior associations persist and remain significant after explicitly accounting for the variance explained by diagnostic groups (HC, MCI, and AD). Second, we performed interaction modeling (MMSE ~ Biomarker × Diagnosis) to formally test for slope heterogeneity between cohorts. For ETVC, no significant interactions were found (all *p >* 0.17, see Table S.5), indicating that the relationship between high-resolution normative deviations and cognitive impairment is statistically consistent across the entire clinical spectrum. For the mean thickness z-score, we observed an attenuation of the slope specifically in the HC group (*p* = 0.052 for the interaction term, Table S.6). However, as visualized in Fig. S.21B, this lack of significant correlation in healthy individuals is attributable to a marked ceiling effect and restricted variance in MMSE scores, rather than a reversal or absence of the underlying biological trend. Collectively, these results demonstrate that the direction of effect is preserved across the clinical spectrum and that the global associations reported in this study are robust to group-level confounds.

**Table S.3.** Mixed Linear Model Results for ETVC. The model (MMSE ~ ETVC + (1|Diagnosis)) treats diagnostic groups as random intercepts. The highly significant *z*-statistic for ETVC confirms the association persists after accounting for group-level baseline shifts.

| Variable | Coef. | Std.Err. | z | P > \|z\| | [0.025, 0.975] |
| --- | --- | --- | --- | --- | --- |
| Fixed Effects |  |  |  |  |  |
| Intercept | 23.5 | 3.1 | 7.645 | 0.000 | [17.5, 29.6] |
| ETVC | -2.6e-04 | 4.0e-05 | -6.532 | 0.000 | [-3.4e-04, -1.8e-04] |
| Random Effects | Variance (Std.Err.) |  |  |  |  |
| Group Variance | 28.2 (7.4) |  |  |  |  |

**Table S.4.** Mixed Linear Model Results for Mean Thickness *z*-score. The model (MMSE ~ Mean Thickness *z* +(1|Diagnosis)) confirms that average cortical thickness remains a significant predictor of cognitive performance even when accounting for diagnostic group as a random effect.

| Variable | Coef. | Std.Err. | z | P > \|z\| | [0.025, 0.975] |
| --- | --- | --- | --- | --- | --- |
| Fixed Effects |  |  |  |  |  |
| Intercept | 22.7 | 3.3 | 6.891 | 0.000 | [16.2, 29.1] |
| Mean Thickness (z) | 0.5 | 0.1 | 4.285 | 0.000 | [0.3, 0.8] |
| Random Effects | Variance (Std.Err.) |  |  |  |  |
| Group Variance | 32.4 (8.3) |  |  |  |  |

**Table S.5.**
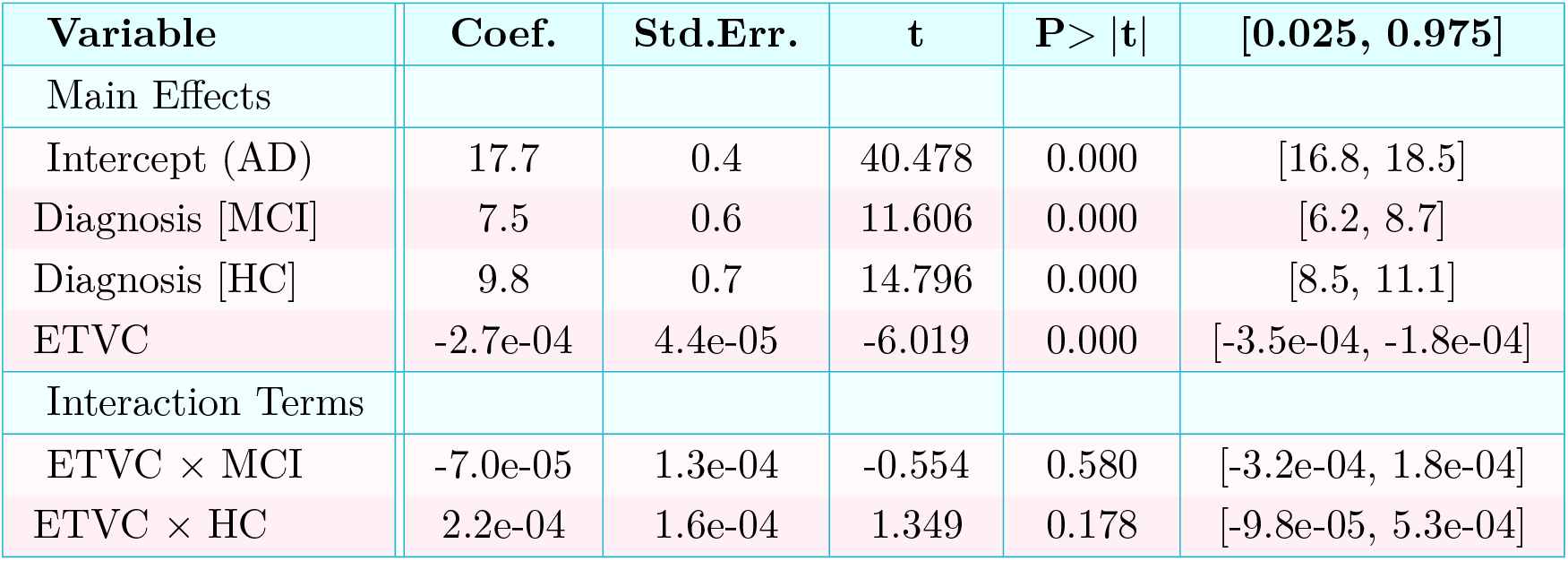
Interaction Analysis for ETVC. The OLS model (MMSE ~ ETVC × Diagnosis) tests for slope differences across groups. Interaction terms (*p >* 0.17) are non-significant, indicating that the relationship between ETVC and MMSE is consistent across diagnostic categories.

**Table S.6.** Interaction Analysis for Mean Thickness *z*-score. The OLS model (MMSE ~ Mean Thickness *z* × Diagnosis) explores slope variations across cohorts. While an attenuation of the slope is observed in the HC group (*p* = 0.052), the overall direction of the effect is consistent across groups, supporting the biological continuum.

| Variable | Coef. | Std.Err. | t | P > \|t\| | [0.025, 0.975] |
| --- | --- | --- | --- | --- | --- |
| Main Effects |  |  |  |  |  |
| Intercept (AD) | 16.6 | 0.4 | 46.266 | 0.000 | [15.9, 17.3] |
| Diagnosis [MCI] | 7.6 | 0.5 | 16.359 | 0.000 | [6.7, 8.5] |
| Diagnosis [HC] | 10.8 | 0.5 | 21.662 | 0.000 | [9.8, 11.7] |
| Mean Thickness ($z$) | 0.7 | 0.2 | 4.393 | 0.000 | [0.4, 1.0] |
| Interaction Terms |  |  |  |  |  |
| Mean Thickness ($z$) × MCI | -0.3 | 0.3 | -0.998 | 0.319 | [-0.9, 0.3] |
| Mean Thickness ($z$) × HC | -0.7 | 0.3 | -1.946 | 0.052 | [-1.4, 0.0] |

#### S.1.13 Heterogeneity Landscape Subgroups

Fig. 8A presents snapshots of cortical deviation patterns across various regions within the 2-dimensional atrophy landscape. To more comprehensively illustrate the overall cortical signature in this landscape, we have provided four supplementary videos (one each for the whole sample, HC, MCI, and AD subsets). These videos display changes in the cortical deviation patterns as the local region is gradually shifted over a circular area within the 2-dimensional space.

#### S.1.14 Robustness of the Heterogeneity Landscape

To assess the robustness of the heterogeneity landscape reported in the main text (Fig. 8), we performed a series of sensitivity analyses examining whether the reported qualitative characteristics depend on the choice of dimensionality reduction algorithm or associated hyperparameters. These analyses were designed to test whether the observed structure reflects stable properties of the underlying high-dimensional deviation space, rather than artifacts of a specific dimensionality reduction procedure.

First, using the same multidimensional scaling (MDS) framework adopted in the main analysis, we repeated the embedding under 3 alternative random seed initializations (Fig. S.22A). Across all runs, the principal visual features described in the Results were consistently reproduced. Specifically, healthy individuals formed a dense central cluster, MCI participants exhibited increased dispersion relative to controls, and AD-diagnosed individuals showed the greatest separation from the healthy cluster, with pronounced heterogeneity in the direction of deviations across individuals. These characteristics were preserved irrespective of seed choice, indicating that the observed structure is not sensitive to this hyperparameter.

Second, to evaluate whether similar landscape organization emerges under different dimensionality reduction paradigms, we repeated the analysis using principal coordinates analysis (PCoA, Fig. S.22B), principal component analysis (PCA, Fig. S.22C), and Isomap (Fig. S.22D). While these methods differ in their geometric assumptions and may yield embeddings with different orientations or relative scaling, all approaches consistently reproduced the core qualitative patterns observed with MDS. In particular, each method revealed a central concentration of healthy individuals, progressive dispersion in MCI, and a highly heterogeneous AD distribution spanning multiple directions in the latent space, without evidence of clear boundaries between potential subtypes.

Taken together with the statistical analyses reported in the Results, including group differences in mean pairwise distances and Hartigan’s dip tests for clusterability^87,88^, these sensitivity analyses indicate that the heterogeneity landscape reflects stable structure present in the high-dimensional normative deviation space. The observed patterns are therefore not specific to any single dimensionality reduction method, but instead represent an emergent property that is both visually apparent across embeddings and quantitatively supported by the underlying distance metrics.

**Supplementary Fig. S.22.**
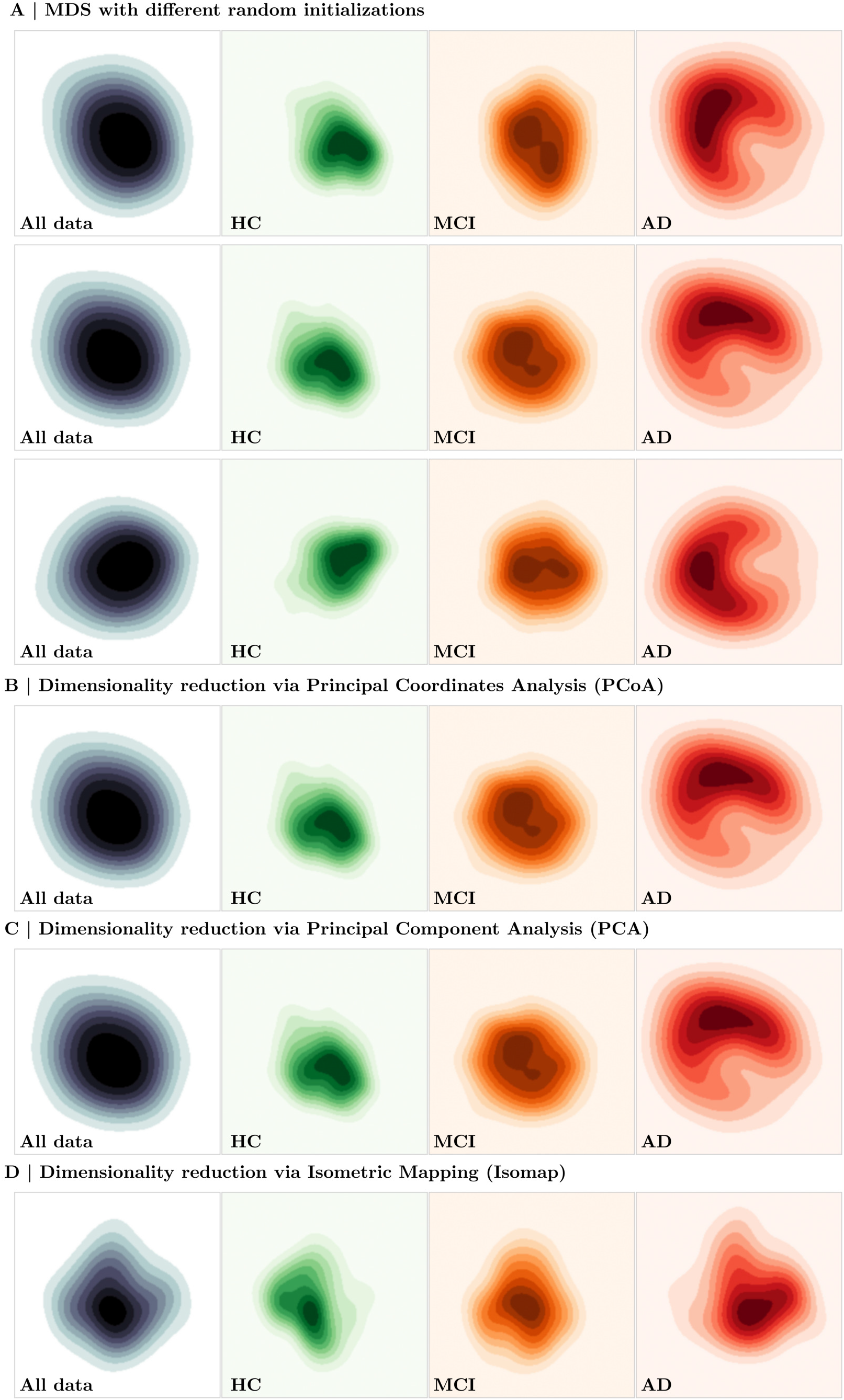
Robustness of the Heterogeneity Landscape to Dimensionality Reduction Choices. Kernel density estimates (KDEs) of the 2D heterogeneity landscape under different dimensionality reduction methods and parameter settings. Columns correspond to the KDE of the **whole sample** (left), followed by **HC, MCI**, and **AD** groups (right). **(A)** Results obtained using multidimensional scaling (MDS) with three different random initializations (top three rows), demonstrating consistent landscape organization across seeds. **(B)** Principal coordinates analysis (PCoA), **(C)** principal component analysis (PCA), and **(D)** Isomap embeddings are shown in subsequent rows. Across all methods, the same qualitative structure is preserved: healthy individuals form a dense central concentration, MCI participants show increased dispersion, and AD-diagnosed individuals exhibit the greatest spread and directional heterogeneity. These results confirm that the heterogeneity landscape is not an artifact of a specific embedding method but reflects stable organization in the high-dimensional normative deviation space.

#### S.1.15 Clusterability of AD Deviation Patterns

The heterogeneity landscape examined above raises a question that visual inspection alone cannot resolve: whether AD-related cortical deviation patterns partition into discrete subtypes with well-defined boundaries, or instead vary continuously. Previous characterizations of AD atrophy subtypes have commonly applied hierarchical clustering to low-dimensional representations of regional imaging features^89,90^, an approach broadly comparable to that adopted here. The number of subtypes has typically been selected using criteria such as the elbow method^90,91^ or the Hopkins statistic^89,92^. However, these approaches do not establish that a partition is warranted: the elbow method compares candidate numbers of clusters without testing whether more than one cluster is supported, whereas the Hopkins statistic can classify unimodal data as clusterable in a substantial proportion of cases^88^. Evidence for discrete subtypes instead requires complementary assessments of multimodality in interindividual distances, overall tendency to cluster, stability under resampling, generalizability across independent samples, and statistical distinguishability from unstructured variation^93,94^. We assessed each of these five aspects in the subsections below, using Ward linkage with Euclidean distances on the *N*_*AD*_ = 208 AD-diagnosed participants, consistent with clustering approaches used in previous subtyping studies^89,90^.

##### S.1.15.1 Unimodality of the Pairwise-Distance Distribution

As briefly reported in the main text (Section 2.7 Individual Atrophy Patterns Form a Continuous Heterogeneity Landscape), we first applied Hartigan’s dip statistic^87^ to the distribution of pairwise Euclidean distances between individual deviation maps. The dip statistic quantifies the maximum departure of an empirical distribution from the closest unimodal distribution and tests the null hypothesis of unimodality against a multimodal alternative. Its relevance to clusterability follows from the expected geometry of distance distributions: well-separated clusters can give rise to distinct within- and between-cluster distance scales, potentially producing multimodality, whereas a unimodal distribution is consistent with separation varying continuously across individuals without clear distance-based discontinuities^88^.

This analysis was performed directly on pairwise distances between vertex-wise deviation maps and therefore did not depend on dimensionality reduction. Distance distributions showed no evidence of multimodality either across the full clinical sample (Dip = 0.0005, *p* = 0.995) or within the AD group (Dip = 0.0014, *p* = 0.996). For completeness, repeating the analysis after dimensionality reduction yielded comparable results (two-dimensional MDS, Dip = 0.0015, *p* = 0.993; two-, ten-, and twenty-component PCA, Dip = 0.0012, 0.0017, and 0.0016, respectively; all *p >* 0.98), indicating that dimensionality reduction neither introduced nor removed distance-based multimodality. As the dip statistic assesses the overall distance distribution rather than a specific partition, it requires neither a prespecified number of clusters nor a choice of clustering algorithm.

##### S.1.15.2 Cluster Tendency

We next assessed whether the deviation maps exhibit an overall tendency to cluster using the gap statistic^95^. For a given number of clusters *k*, the gap statistic compares the within-cluster dispersion *W*_*k*_ of the observed data with its expectation under a reference distribution containing no cluster structure, Gap(*k*) = E^∗^[log *W*_*k*_]−log *W*_*k*_. Clustered data exhibit smaller within-cluster dispersion than expected under the reference distribution, resulting in larger gap values. Critically, the gap statistic is defined at *k* = 1 and can therefore directly assess whether partitioning the sample improves upon treating it as a single group. As the heterogeneity landscape provides our primary two-dimensional representation of interindividual variation, we evaluated cluster tendency in this embedding and repeated the analysis on the first two principal components to determine whether the conclusion depended on the choice of dimensionality reduction.

We evaluated *k* = 1 to 10 using 1000 reference datasets drawn uniformly within a bounding box aligned to the principal components of the observed data (Tibshirani’s method (b)^95^). Each reference dataset was clustered using Ward linkage with Euclidean distances, identically to the observed data, and standard errors were computed across reference draws. The recommended selection rule identifies the smallest *k* satisfying Gap(*k*) ≥ Gap(*k* + 1) − *s*_*k*+1_, which yielded *k*^∗^ = 1 in the two-dimensional MDS landscape (Supplementary Fig. S.23A). Moreover, Gap(1) exceeded Gap(*k*) − *s*_*k*_ for every *k* = 2 to 10 (Supplementary Fig. S.23B). Gap values were similar across *k* = 1 to 6 (0.44–0.54, with overlapping standard-error intervals) and declined thereafter. Repeating the analysis on the two-component PCA representation yielded the same result (*k*^∗^ = 1, with the criterion satisfied against every larger *k*; Supplementary Fig. S.23C,D). Thus, across both representations, the gap statistic provided no evidence that partitioning the AD deviation maps improved upon a single-cluster solution.

**Supplementary Fig. S.23.**
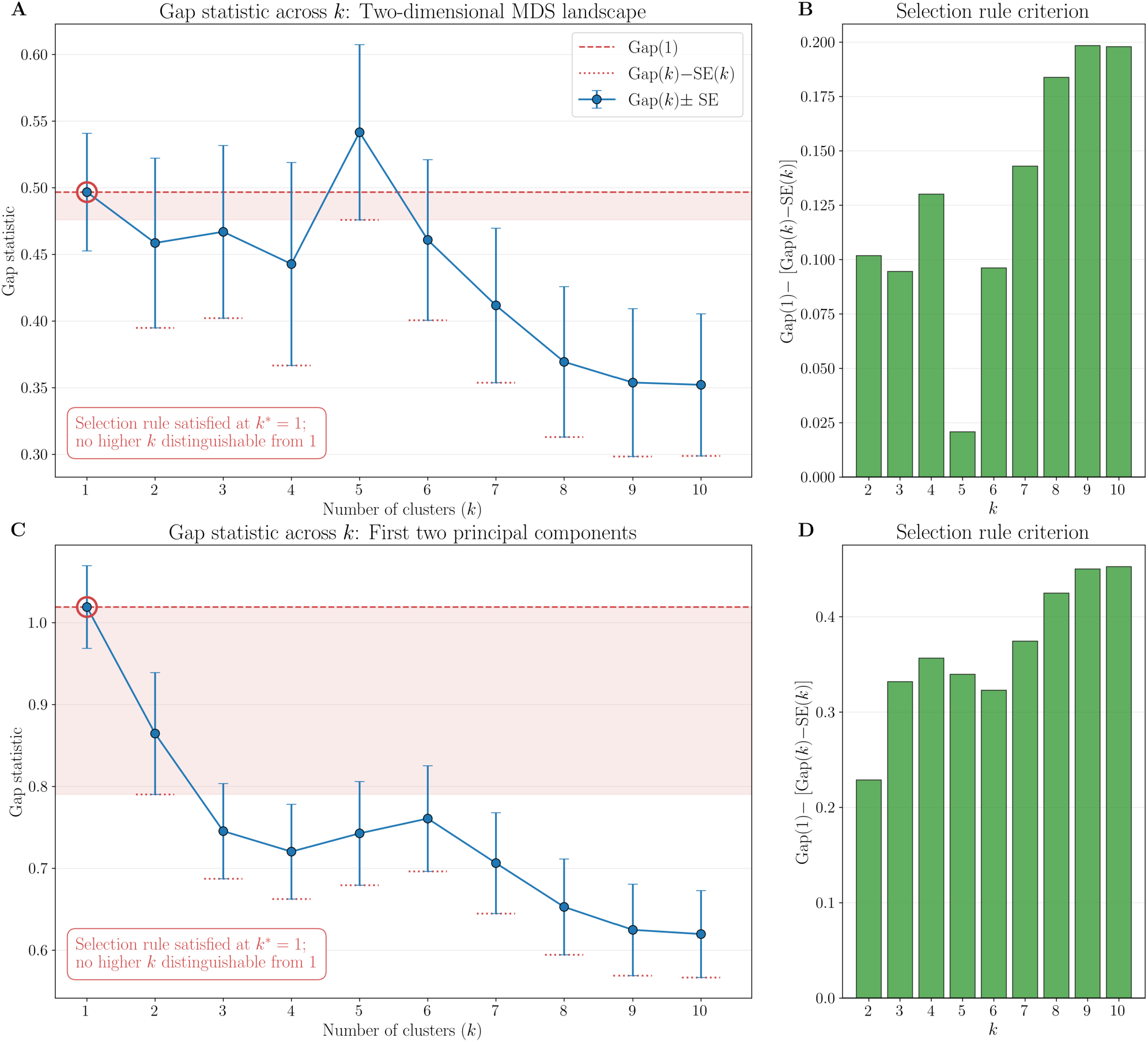
Cluster Tendency of AD Deviation Patterns. **(A)** Gap statistic across candidate numbers of clusters, computed on the two-dimensional heterogeneity landscape against 1000 uniform reference datasets. Error bars denote standard errors across reference draws; short dotted markers indicate Gap(*k*) − *s*_*k*_ for each *k* ≥ 2, and the dashed line and shaded band mark Gap(1) and its margin above the largest of these values. The circled point denotes the selected *k*^∗^ = 1. **(B)** The selection criterion evaluated against every larger *k*, expressed as Gap(1) − [Gap(*k*) − *s*_*k*_]; positive values indicate that a single cluster is preferred over *k* clusters. **(C, D)** The same analyses repeated on the first two principal components of the vertex-wise deviation maps. In both representations the criterion is satisfied for all *k*, and a single cluster is preferred.

##### S.1.15.3 Stability Under Resampling

A partition reflecting genuine structure should be reproducible when the sample is perturbed. We therefore assessed cluster-wise stability following established recommendations^96^. Briefly, the data were repeatedly resampled and independently clustered, after which each cluster in the original solution was matched to its most similar counterpart in the resampled solution. Agreement was quantified using the Jaccard similarity between cluster membership sets and averaged across resamples. This clusterwise assessment is important because a partition may contain a reproducible cluster alongside others that dissolve under resampling, a distinction that can be obscured by a single global stability index^96^.

**Supplementary Fig. S.24.**
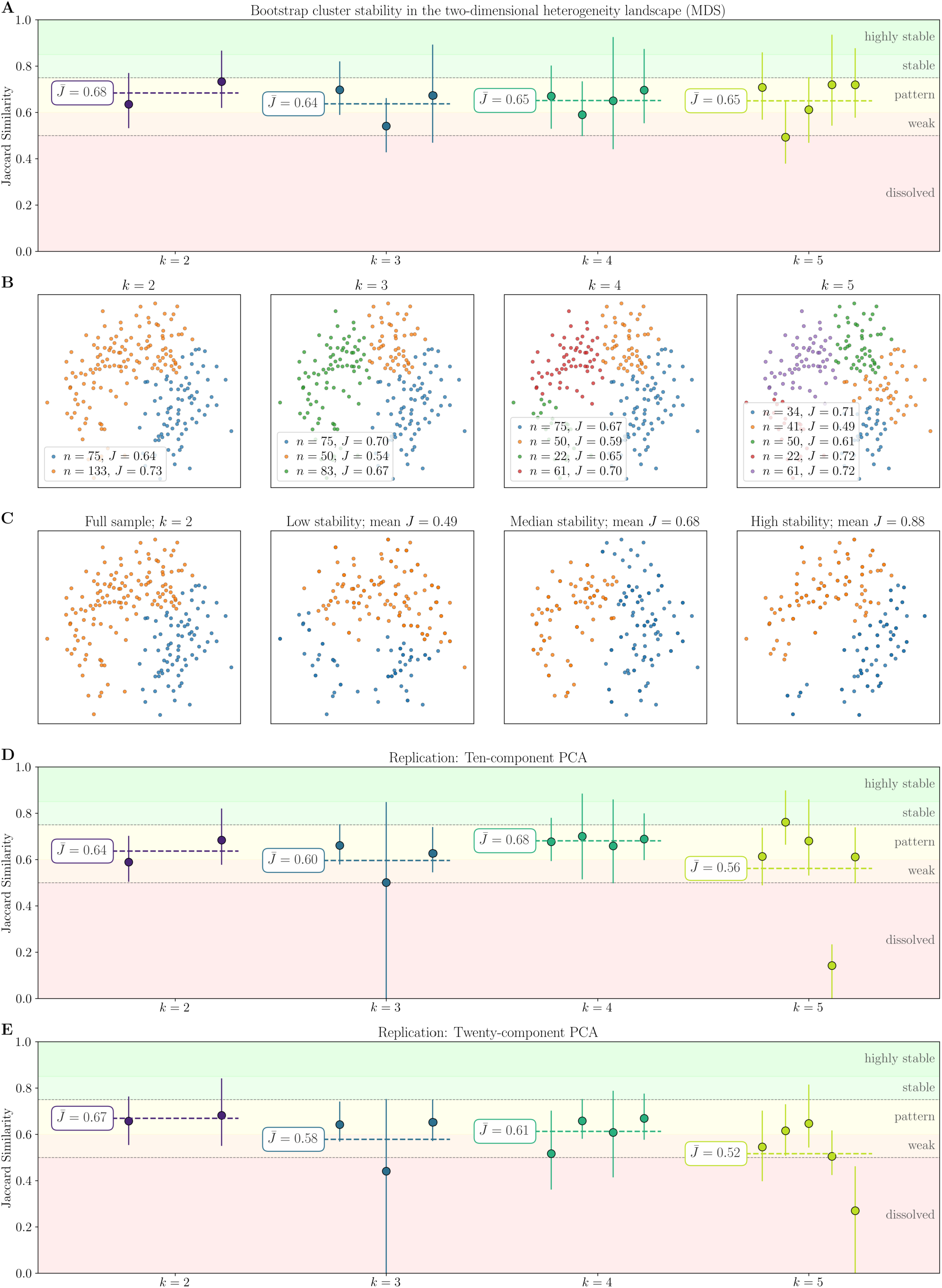
Stability of Candidate Partitions Under Resampling. **(A)** Cluster-wise stability across *k* = 2 to 5 in the two-dimensional heterogeneity landscape, assessed over 1000 bootstrap resamples. Points denote the mean Jaccard similarity between each cluster in the full-sample solution and its matched counterpart in the resampled solution, with vertical lines indicating the corresponding interquartile range across resamples. Dashed horizontal lines and annotations denote the mean stability across all clusters within each partition 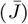. Background colors indicate the interpretation ranges described in the text: dissolved (*<* 0.5), weak (0.5–0.6), pattern (0.6–0.75), stable (0.75–0.85), and highly stable (*>* 0.85). **(B)** The heterogeneity landscape colored according to cluster assignment at each *k*, with cluster size and per-cluster stability indicated. **(C)** The full-sample partition at *k* = 2 alongside three bootstrap realizations spanning the observed range of agreement, illustrating the variability of cluster membership under resampling. **(D, E)** The same stability assessment in 10- and 20-component principal component representations of the vertex-wise deviation maps.

We evaluated *k* = 2 to 5 using 1000 bootstrap resamples for each *k*, matching clusters between the original and resampled solutions by Hungarian assignment on the Jaccard similarity matrix. Following commonly used interpretation thresholds^96,97^, mean Jaccard values below 0.5 indicate dissolved clusters, values between 0.5 and 0.6 indicate weak patterns that should not be trusted, values between 0.6 and 0.75 indicate existence of patterns whose membership is unreliable, values above 0.75 indicate valid stable clusters, and values above 0.85 indicate highly stable clusters. We additionally summarized each candidate partition by the mean Jaccard similarity across its constituent clusters 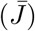, providing an overall measure of its reproducibility. We considered a partition viable as a subtyping scheme only if its constituent clusters were all stable, since a solution containing clusters that are not reproducible cannot support assignment of individuals to subtypes.

No candidate partition exhibited stable cluster structure (Supplementary Fig. S.24A). In the two-dimensional landscape, individual cluster stabilities ranged from 0.49 to 0.73 across *k* = 2 to 5, with no cluster exceeding the stable-cluster threshold; moreover, every solution beyond *k* = 2 contained at least one cluster in the weak or dissolved range. The most reproducible solution was 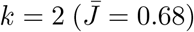, which nevertheless remained below the stable-cluster threshold. The corresponding partitions are shown in Supplementary Fig. S.24B. Supplementary Fig. S.24C further illustrates this instability: across bootstrap resamples of the *k* = 2 solution, mean agreement with the full-sample partition ranged from 0.49 to 0.88, with lower-agreement resamples reassigning a substantial proportion of individuals between clusters. This result was replicated in the 10- and 20-component principal component representations (Supplementary Fig. S.24D,E), where the most reproducible solutions likewise remained below the stable-cluster threshold (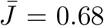 at *k* = 4 and 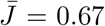 at *k* = 2, respectively). Collectively, these results provide no evidence that any of the candidate partitions examined were reproducibly recovered under resampling.

##### S.1.15.4 Generalizability Across Independent Subsamples

Beyond reproducibility under resampling, a partition reflecting genuine structure should generalize across independent samples. We assessed this property using prediction strength^98^. The sample was repeatedly divided into two halves, which were independently clustered at a given *k*. One half served as a training set, whose cluster centroids were used to assign observations in the other half, and the resulting assignments were compared with the clustering independently obtained in that test half. Agreement was assessed through within-cluster co-membership: for each cluster identified in the test half, we calculated the proportion of pairs of its members that were also assigned to the same cluster using the training-derived centroids. In practice, a genuinely reproducible cluster remains largely intact under this independent assignment, whereas a poorly generalizable cluster is divided across multiple predicted groups. Prediction strength is defined as the minimum of these proportions across test clusters, reflecting the principle that a partition generalizes only as well as its least reproducible constituent cluster^98^. It thus provides a partition-level assessment of generalizability, complementing the cluster-wise stability analysis described above.

We evaluated *k* = 2 to 10 across 500 random splits per *k*, assessing each split in both directions and thus obtaining 1000 estimates per *k*. Clustering used Ward linkage with Euclidean distances, as elsewhere, with nearest-centroid assignment as the classification rule following the original formulation^98^. Because prediction strength for *k* = 1 is trivially unity, the single-cluster solution is selected by default when no larger *k* satisfies the recommended criterion; values above 0.8, or more stringently 0.9, indicate well-supported cluster structure^98^. As in the preceding analyses, we repeated the assessment across multiple representations: the two-dimensional heterogeneity landscape and two-, ten-, and twenty-component PCA representations.

In the two-dimensional landscape, prediction strength declined monotonically from 0.60 at *k* = 2 to 0.29 at *k* = 10 (Supplementary Fig. S.25A). Even for *k* = 2, the strongest candidate partition, prediction strength remained well below the threshold (IQR: 0.50–0.67). PCA representations yielded closely comparable results, with mean prediction strength of approximately 0.61–0.62 at *k* = 2 and lower values for all larger *k* (Supplementary Fig. S.25B–D). Thus, across all four representations, no candidate partition (*k* ≥ 2) met the recommended criterion for generalizable cluster structure, supporting retention of the single-cluster solution.

**Supplementary Fig. S.25.**
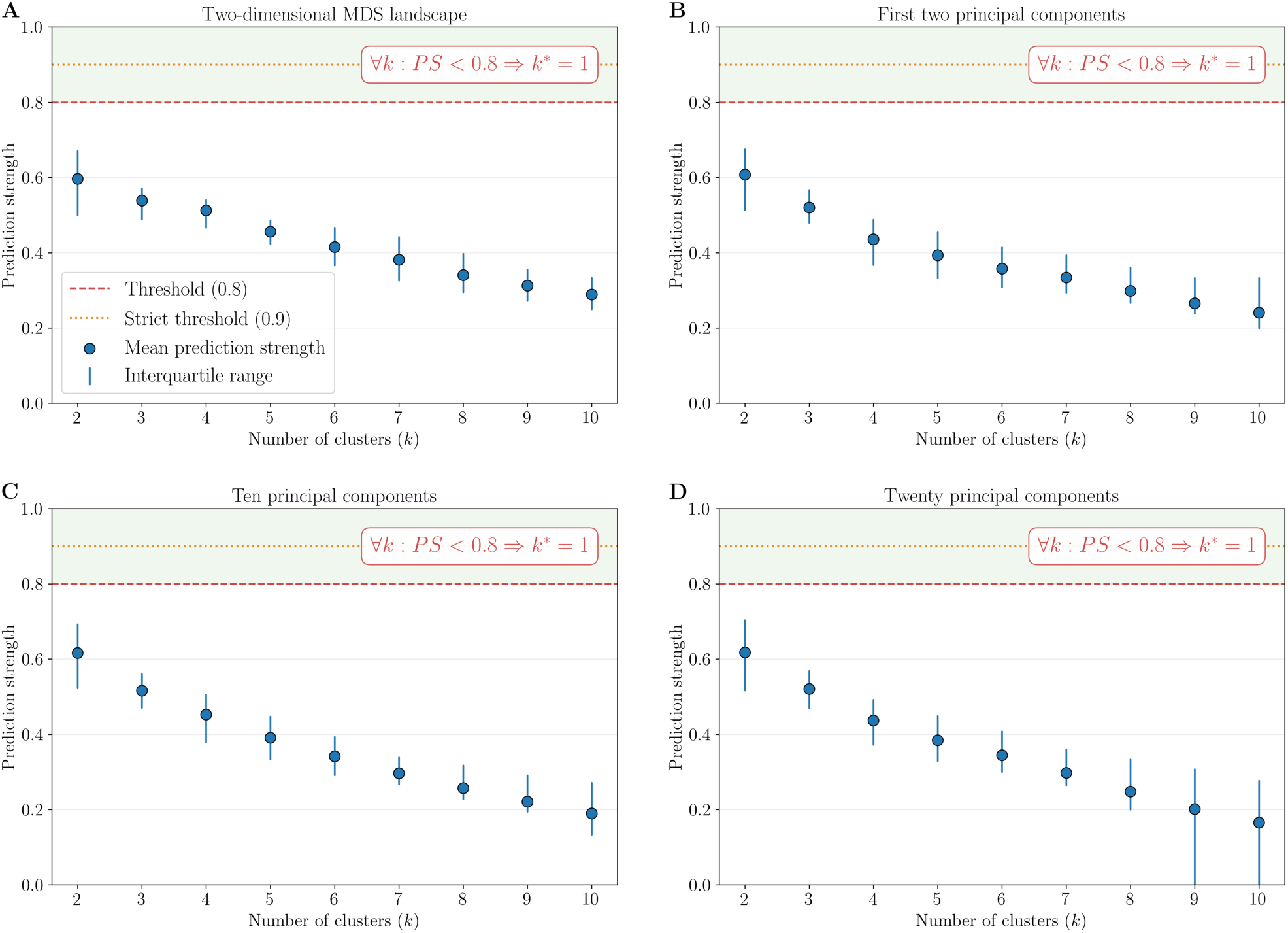
Generalizability of Candidate Partitions Across Independent Subsamples. Prediction strength for *k* = 2–10, estimated from 1000 split-half evaluations per *k* in **(A)** the two-dimensional heterogeneity landscape and **(B–D)** two-, ten-, and twenty-component PCA representations of the vertex-wise deviation maps. Points denote mean prediction strength, and vertical lines indicate the interquartile range across splits. Dashed and dotted horizontal lines mark the recommended threshold of 0.8 and the more stringent threshold of 0.9, respectively. Across all four representations, no candidate partition reached the recommended threshold.

##### S.1.15.5 Statistical Significance Against a Gaussian Null

The preceding criteria assess whether candidate partitions are reproducible or generalizable, but do not directly test whether observed cluster separation is statistically distinguishable from data lacking discrete cluster structure. We therefore applied SigClust2^99^, which evaluates each split of a hierarchical clustering against the null hypothesis that the data arise from a single multivariate Gaussian distribution. Separation at a given split is quantified by the two-means cluster index, defined as the proportion of total variance remaining within the two resulting groups after splitting; lower values therefore indicate stronger separation. The observed index is compared with its distribution under simulated Gaussian data, and hierarchical splits are tested sequentially from the root of the dendrogram downwards, with descendant nodes tested only if their parent attains significance, thereby controlling the family-wise error rate across the tree^99^. Thus, the test asks whether an empirical split produces greater separation than expected under a null model without discrete cluster structure.

We applied this procedure using the `sigclust2` R package^99^ to the two-dimensional embedding and the ten- and twenty-component principal component representations, using Ward linkage with Euclidean distances, 10,000 simulated null datasets per node, and *α* = 0.05. The package provides two procedures for clustering the simulated null datasets, and we report both to verify that our conclusions were not contingent on this choice: the hierarchical procedure recommended for rotation-invariant metrics such as Euclidean distance, and a 2-means alternative providing a more conservative test^99^.

The top-level split was not significant in any representation, and testing therefore terminated at the root without proceeding to lower-level splits (Supplementary Fig. S.26). Under the hierarchical null procedure, no representation reached the significance threshold (*p* = 0.062, *p* = 0.866, and *p* = 0.758 for the two-dimensional, ten-, and twenty-component representations, respectively). The conservative 2-means procedure likewise yielded no significant separation (*p* = 0.376, *p >* 0.999, and *p* = 0.995, respectively). Thus, no hierarchical split attained significance under either procedure or any representation, providing no evidence of discrete cluster structure beyond that expected under the Gaussian null.

**Supplementary Fig. S.26.**
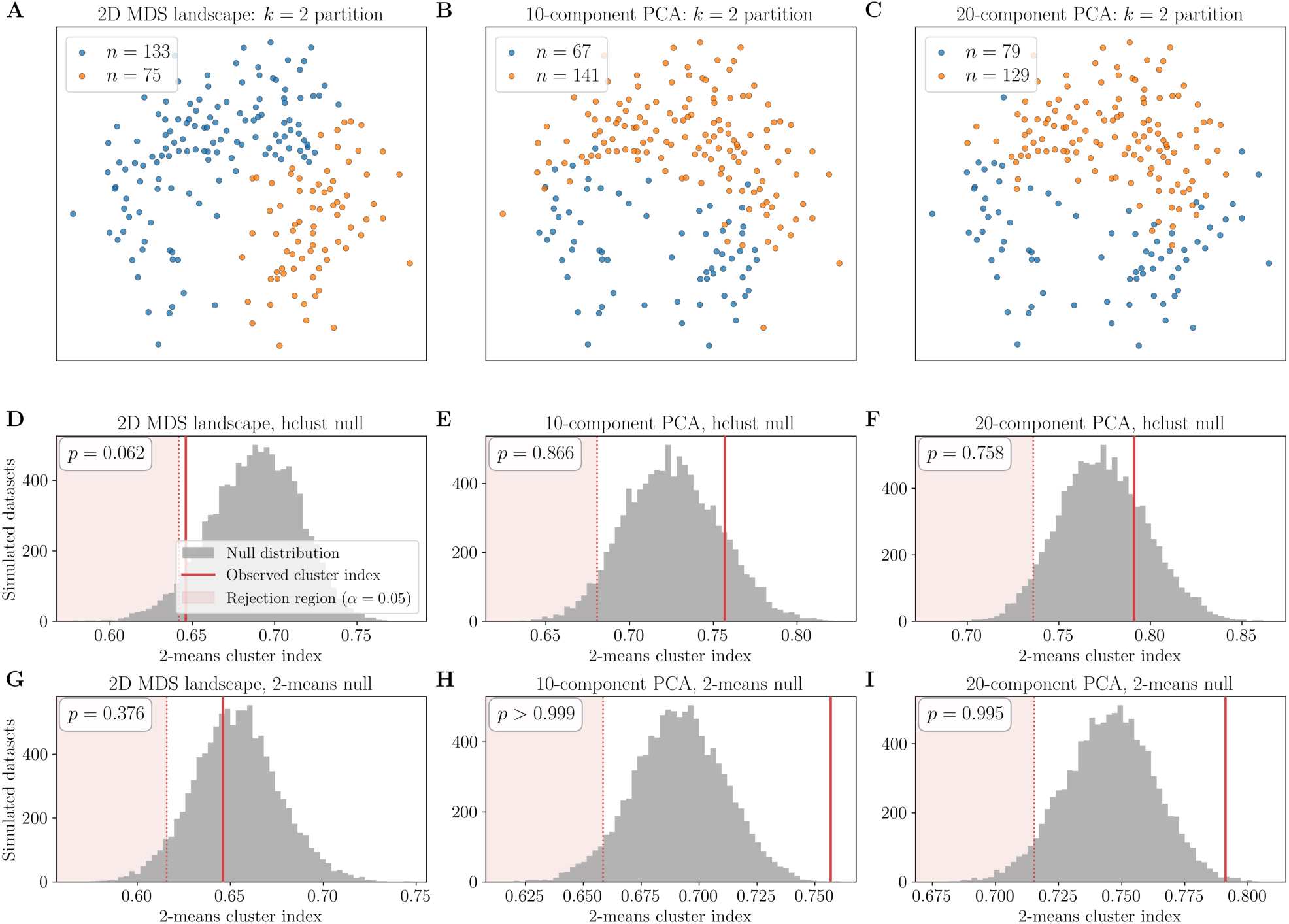
Statistical Significance of Cluster Splits. **(A–C)** Top-level (*k* = 2) partitions tested in the two-dimensional heterogeneity landscape **(A)** and the ten- **(B)** and twenty-component **(C)** principal component representations, with the latter two projected onto the same 2D landscape for visualization. **(D–F)** Null distributions of the two-means cluster index for the root split in each representation, generated from 10,000 datasets simulated under a single-Gaussian null and clustered using the hierarchical procedure. **(G–I)** Corresponding null distributions obtained using the 2-means procedure. In **(D–I)**, the observed cluster index is marked in red, with lower values indicating stronger separation; shaded regions and dotted lines denote the lower 5% of each null distribution and the corresponding rejection threshold at *α* = 0.05, respectively. Across all representations and null procedures, no split showed significantly greater separation than expected under the Gaussian null.

## Footnotes

∗ Access to the MACC clinical imaging dataset is subject to a data-transfer agreement with the Memory, Ageing & Cognition Centre, National University of Singapore (http://www.macc.sg/).

